# Methylation risk scores are associated with a collection of phenotypes within electronic health record systems

**DOI:** 10.1101/2022.02.07.22270047

**Authors:** Mike Thompson, Brian L. Hill, Nadav Rakocz, Jeffrey N. Chiang, IPH, Sriram Sankararaman, Ira Hofer, Maxime Cannesson, Noah Zaitlen, Eran Halperin

## Abstract

Inference of clinical phenotypes is a fundamental task in precision medicine, and has therefore been heavily investigated in recent years in the context of electronic health records (EHR) using a large arsenal of machine learning techniques, as well as in the context of genetics using polygenic risk scores (PRS). In this work, we considered the epigenetic analog of PRS, methylation risk scores (MRS), a linear combination of methylation states. Since methylation states are influenced by both environmental and genetic factors, we hypothesized that MRS would complement PRS and EHR-based machine-learning methods, improving overall prediction accuracy. To evaluate this hypothesis, we performed the largest assessment of methylation risk scores in clinical datasets to be conducted to date. We measured methylation across a large cohort (n=831) of diverse samples in the UCLA Health biobank, for which both genetic and complete EHR data are available. We constructed MRS for 607 phenotypes spanning diagnoses, clinical lab tests, and medication prescriptions. When added to a baseline set of predictive features, MRS significantly improved the imputation of 139 outcomes, whereas the PRS improved only 22 (median improvement for methylation 10.74%, 141.52%, and 15.46% in medications, labs and diagnosis codes, respectively, whereas genotypes only improved the labs at a median increase of 18.42%). We added significant MRS to state-of-the-art EHR imputation methods that leverage the entire set of medical records, and found that including MRS as a medical feature in the algorithm significantly improves EHR imputation in 37% of lab tests examined (median *R*^2^ increase 47.6%). Finally, we replicated several MRS in multiple external studies of methylation (minimum p-value of 2.72 *×* 10*^−^*^7^) and replicated 22 of 30 tested MRS internally in two separate cohorts of different ethnicity. In summary, our work provides a comprehensive evaluation of MRS in comparison to PRS and EHR imputation on the largest dataset consisting of methylation, genotype, and EHR data. Our publicly available results and weights show promise for methylation risk scores as clinical and scientific tools.

## 1 Introduction

Widespread adoption of electronic health record systems coupled with an increasing interest in hospital biobanking systems has spurred research efforts spanning machine-learning and genomics communities[1–7]. These efforts have produced increasingly accurate imputation (current state) and prediction (future state) of patient phenotypes from medical records [8, 9] and polygenic risk scores [1–3, 10–14], and are already being investigated in translational contexts [15–18]. For example, recent work has shown that machine learning can leverage high-dimensional data to aid in the prediction of a multitude of clinical phenotypes including cardiac function and arrhythmia [19–21], post-operative complications [8, 9], sepsis [22], breast cancer [11, 23], and prostate cancer [24]. Nonetheless, a genetics-based predictor such as the polygenic risk score may be limited in predictive utility as it does not account for changes in disease risk—for example, due to age, or changes in environment—throughout one’s lifespan [13].

In this work we examine the potential for epigenetic information to improve phenotype inference in combined biobank-EHR systems. As DNA methylation, henceforth referred to as simply “methylation”, is affected by both genetics and environment—such as lifestyle choices, diet, exercise, and smoking status—it captures multi-factorial information about predispositions to clinical conditions [25–31]. Moreover, methylation is readily available for use in existing biobanks that collect DNA samples, and recent advancements in methylation profiling technologies have enabled an abundance of large-scale studies of methylation and its role as a biomarker for a variety of phenotypes and health-related outcomes [25, 31–37]. It is therefore a natural candidate for an extension of PRS, and we hypothesized that methylation can be used to complement genetics as a clinical prediction tool. To that end, we have generated and evaluated methylation risk scores (MRS), which are linear combinations of CpG methylation states[25].

To comprehensively investigate the utility of MRS and characterize its properties, we conducted a study of 607 EHR-derived phenotypes spanning medications (e.g. vasopressers, glucocorticosteroids, fluoroquinolones), labs (e.g. creatinine, glucose, prothrombin time), and diagnoses (e.g. T2D, bacterial pneumonia, anemia) that were available for a sufficient number of patients in the cohort. The cohort contained 831 patients—to the best of our knowledge, the largest epigenetic biobank dataset to date (including genetics, methylation, and EHR)—from the UCLA Health ATLAS cohort across a wide range of ages (18-90), racial and ethnic groups, and overall health (including patients ascertained on kidney and heart disease, with matched controls), with corresponding genetic and EHR data. This provides the opportunity to study the potential contribution of methylation to larger biobanks and in multiple clinical contexts. We find that the MRS-based imputations were more informative compared to PRS in 84 (92%) medications, 32 (94%) labs, and 123 (82%) diagnoses, more than doubling the imputation accuracy in over half of the outcomes considered. We also show that the MRS improves the imputation accuracy over PRS for cases in which the PRS is trained on very large external biobanks (roughly 3 orders of magnitude larger), as opposed to 831 samples that are available in this study. We observe that MRS improves over PRS learned from large biobanks in 40% of the tested phenotypes. Further, as our cohort was ethnically diverse, we performed replicability analyses within each racial and ethnic subset of our data. We broadly showed the replicability of the five best-imputed (by MRS) medications, labs, and diagnoses—46% and 100% of which replicated in (n=118) non-white Hispanic-Latino- and (n=543) white non-Hispanic-Latino-identifying individuals respectively. Finally, we demonstrate the ability of MRS to transfer between methylation arrays and cohorts by conducting an association study of kidney-related MRS in an external diabetic nephropathy EWAS [38], where the minimum replication p-value was 2.72 *×* 10*^−^*^7^.

These results provide evidence for the utility of methylation in phenotype imputation in general, and in biobank settings in particular. However, the promise of clinical translation of genomic risk scores, including PRS or MRS, is highly dependent on the clinical context of the patient. There is a large body of work investigating phenotype imputation and prediction in clinical settings using EHR data alone, typically with machine learning techniques, without any genomic data. To the best of our knowledge, the question of whether genomic data can be used to complement such algorithms has not been studied. Since the application of MRS or PRS to clinical data without taking into account the EHR data provides a limited clinical utility, this is a natural question.

Here, we demonstrate that MRS can be used in conjunction with EHR data to improve the imputation of clinical data of patients. Critically, most machine learning approaches rely on imputation because of the inability of such algorithms to process missing data, making accurate imputation a crucial step. We found that the combination of MRS with a gold standard imputation approach—SoftImpute [39]—for clinical data imputation, provides improved accuracy (*R*^2^) in 37.3% of the examined phenotypes with a median increase of 47.6%. This result provides the potential to improve machine learning algorithms that use the EHR data, by complementing the data with methylation information for the patients.

In summary, our results quantify the contribution of methylation information in clinical settings, both in isolation and in conjunction with the EHR data, and they demonstrate the potential utility of epigenetic biobanks in clinical settings.

## 2 Results

### Risk model description

Analogous to the PRS[40, 41], we defined the MRS by a linear combination of *m* CpG site beta values *c* and weights *w*:

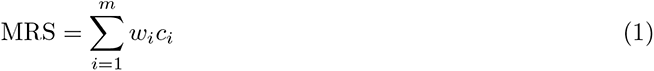

To ensure the methylation risk score added predictive value over commonly captured features (e.g. age and sex), we created a baseline predictive model that included patients’ age, sex, reference-based methylation cell-type composition estimates [42], self-reported race-ethnicity, self-reported smoking status, and the first ten genetic principal components [27] (see Supplementary Table S1 for cohort demographic data). We fit the baseline model using a linear or logistic regression model depending on whether the outcome was continuous or binary. We compared the baseline model to models that included the baseline features as well as either methylation or genotype data. For both the MRS and PRS, we used regression with LASSO, elastic net, and ridge regularization over the genomic features while treating the baseline features as fixed covariates. We fit all models using 10-fold double cross-validation, wherein each training set an additional cross-validation was performed for hyperparameter selection, then this training-set cross-validated model was used to predict the held-out test set. We tested for significance using an association test (via linear regression) between the cross-validated predicted outcome (i.e. the concatenated predictor across all folds) and the true outcome. For full details see Methods.

### Methylation risk scores significantly outperform the baseline and PRS models

From our EHR database, we extracted diagnosis codes , medication orders, and the most recent lab results, all of which occurred before the methylation samples were collected. We aggregated the ICD codes into higher-level phenotypes according to the phenotype code (Phecode) mapping proposed by Denny et al. [43, 44] and grouped individual medications by pharmaceutical subclass to increase generalizability and power.

We trained penalized linear models to predict clinical phenotypes for which there was a sufficient number of patient data available, which included 168 medication subclasses, 69 lab values, and 370 Phecodes. Using a Bonferroni-adjusted association test, the baseline and MRS models significantly imputed the usage of 69 and 88 medications, 18 and 33 labs, and 106 and 139 Phecodes respectively (Figure S1). We compared the performance of the MRS to a model that used both the PRS and baseline features on the same set of individuals, which significantly imputed the usage of 53 medications, 20 lab results, and 93 Phecodes. Notably, the baseline model imputed a greater number of medications and Phecodes than models that leveraged a PRS, which suggests that including genomic features may either add noise or our sample size may not have been sufficient to discover their effects for certain outcomes. We also show in Figure S2 that the baseline model gains some of its predictive power from genomics-derived features like ancestry PCs or estimated cell counts, and therefore a PRS or MRS may not offer a substantial improvement over these features for certain outcomes under the current sample sizes.

Next, we investigated outcomes for which genomics-based predictors add predictive power to the baseline features and, in such cases, the extent to which their inclusion improves predictive accuracy. On the outcomes for which the genomics-based predictors produced statistically significant imputations, we conducted a likelihood ratio test comparing an association test of the true outcome using the cross-validated baseline predictor alone, to a model that included the cross-validated baseline predictor as well as the cross-validated predictor that included both baseline and genomic features (Methods). The methylation significantly improved the baseline predictor for 54 medications, 29 labs, and 56 Phecodes, and led to a median increase of 10.74%, 141.52%, and 15.46% over the baseline predictor’s accuracy (AUC, *R*^2^) in each outcome, respectively (Figure 1). The genotypes significantly improved the baseline predictor for 8 medications, 3 labs, and 11 Phecodes, and led to a median increase of 18.42% over the baseline in the *R*^2^ of the labs, but a median decrease of 1.75% and 0.94% in AUC of the medications and Phecodes respectively (Figure 1). We note that our internal sample size is likely underpowered to discover small genetic effects and therefore suggest the contributions made by the genotypes may be due to additional ancestry signal that was not captured by the first few genetic PCs.

**Figure 1.**
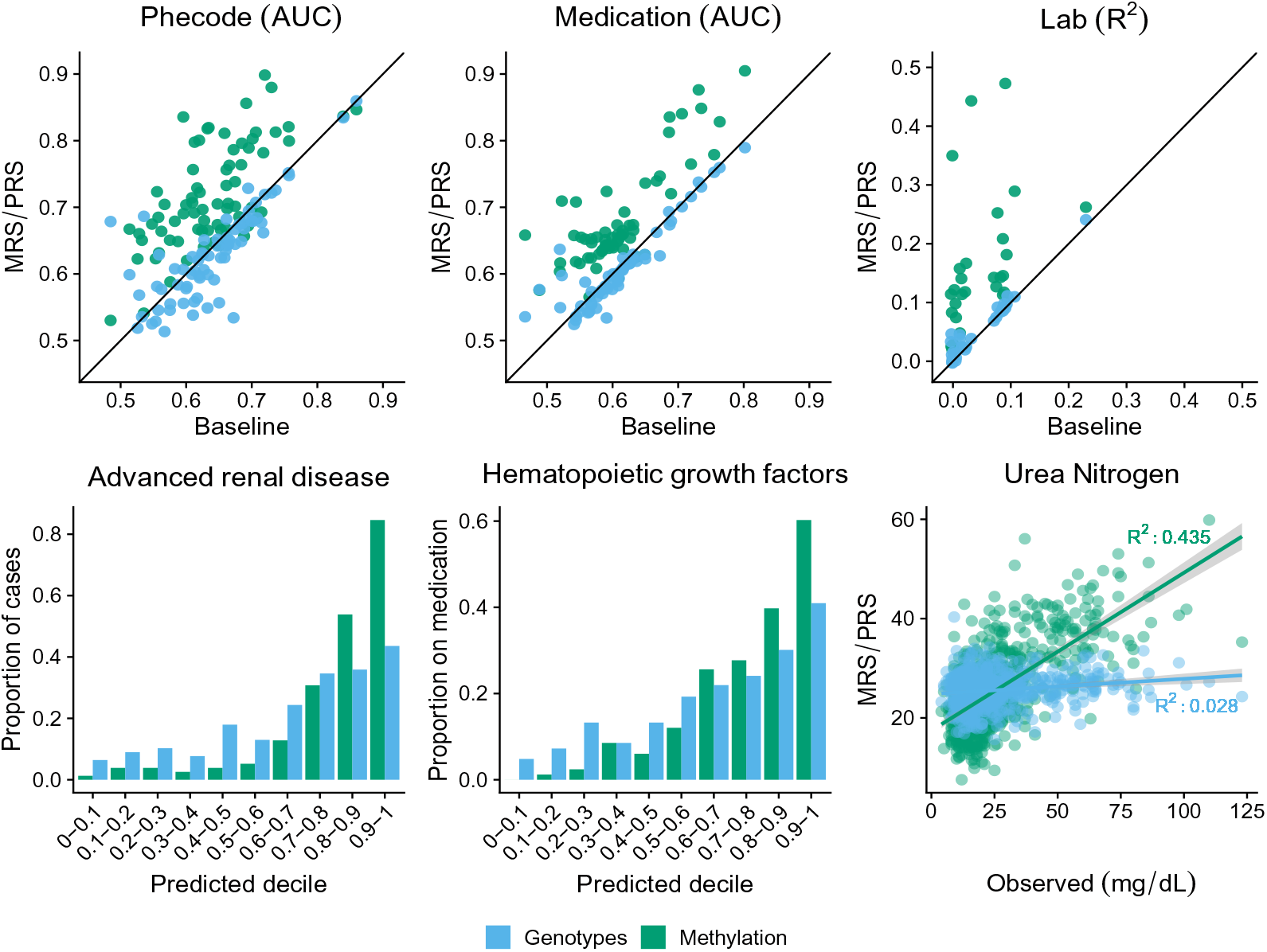
MRS increases imputation accuracy on a variety of outcomes. (Top) The performance of the PRS (blue) and MRS (green) imputations on the y-axis with the baseline model performance on the x-axis. The performance of binary phenotypes (Phecodes, medications) is measured using area under the ROC curve (AUC) and the performance of continuous phenotypes (lab results) is measured using proportion of variance explained (*R*^2^). Shown is the performance on the union of outcomes that were significantly improved over the baseline model by either the MRS or PRS and that were significantly imputed their corresonponding predictor (72 Phecodes, 59 medications, and 31 labs). (Bottom) The disease incidence as a function of the PRS (blue) and MRS (green) binned by deciles (left, middle); and the observed Urea Nitrogen lab result value plotted against its imputed value (right).

The medications that improved the greatest using methylation corresponded to drugs often prescribed to individuals with neutropenia (hematopoeitic growth factors, AUC baseline .706 95% CI [.661,.748] to AUC methylation .840 [.807,.871]) or chronic kidney disease (phosphate binder agents AUC from .731 [.683, .777] to .876 [.842, .907]). The lab panels best improved with the addition of the methylation-based predictor included those related to kidney function as well as cell counts (Urea nitrogen baseline adjusted *R*^2^ .032 [.007,.057] compared to .443 [.377,.509] with methylation, hemoglobin .107 [.063,.151] to .289 [.232,.346]). The addition of the genotype-based predictor improved the imputation of hematocrit (adjusted *R*^2^ from .077 [.041,.114] to .092 [.052,.132]) and total protein (adjusted *R*^2^ .094 [.047,.141] to .111 [.060,.162]), both of which are influenced by ancestry [45, 46]. In the context of Phecodes, methylation greatly increased the imputation of advanced renal disease over the baseline and genotype models (for example, AUC baseline .720 [.673,.762] to 0.898 [.867,.927] with methylation), and the genotype model increased the imputation of actinic keratosis (AUC from .694 [.631,.747] to .728 [.672,.784]).

Overall, when looking at the intersection of medications significantly imputed by either the methylation and genotypes or methylation and baseline, 92% were better imputed by methylation sites than genotypes (median 9.13% increase) and 78% were better imputed by methylation compared to the baseline (median 6.81% increase). Methylation improved the baseline imputation accuracy by over 15% for 14 medications. In the context of significantly imputed lab values, methylation explained more variability than the baseline (median 398% increase) and genotype (median 274% increase) predictors in 97% and 94% of the respective union of significantly imputed labs. For 22 labs, the percent increase of imputation accuracy was greater than 15% over the baseline model. Methylation was more accurate than the baseline (median 3.48% increase) or genotypes (median 6.58% increase) for 70% and 83% of each respective union of Phecodes. For 29 Phecodes, the methylation offered over a 15% increase in predictive accuracy compared to the baseline model. For a substantial proportion of outcomes, the MRS predictor added statistically significant predictive value over the PRS predictor (Figure S3). This was generally not true when comparing whether the PRS added predictive value over the MRS. For the imputation performance on the full list of phenotypes, see Supplemental Tables S2, S3, and S4. To see the number of CpGs selected for each MRS, see Supplemental Tables S5, S6, and S7.

Importantly, cell-type composition, age, sex, BMI, smoking status and ancestry provide sufficient power for the imputation of many EHR outcomes. We show explicitly in Supplementary Figure S4 that genomics derived features such as cell-type composition and ancestry PCs likely contribute to accurate imputation of several outcomes. In our analyses, we directly compared the power gained by methylation over the aforementioned set of baseline features. However, we note that obtaining these baseline features may be unnecessary as the methylation alone may capture their signal [27, 30, 32, 47, 48]. Further, previous reports have suggested that approaches that fit all methylation probes simultaneously with regularization may perform better when excluding latent confounders, such as cell type composition [49]. We therefore suggest that using the methylation alone is sufficient to replicate a substantial proportion of the associations generated from the baseline features alone.

### Using methylation risk scores improves imputation approaches

Due to significant heterogeneity in patient populations, the diagnosis and treatment process can vary widely between patients, causing many variables to be left unobserved. This sparse structure in the data must be reconciled before performing many downstream analyses, and the imputation accuracy of these unobserved variables is therefore crucial to subsequent steps. A commonly-used approach for imputation is matrix completion, for example, SoftImpute [39], where the data matrix is reconstructed from a low-rank representation. Often, one would jointly use demographic information, diagnosis codes, lab results, and medications to generate an estimate of the unobserved EHR values using an imputation method such as SoftImpute, and therefore we used this as our baseline imputation estimate [50].

To investigate whether methylation can add additional useful information to the imputation, we included the MRS values as part of imputation procedure and compared the performance to the estimates that do not take methylation data into account (see Methods). Specifically, we included cross-validated MRS values for diagnosis codes, lab results, medications, and demographics that were significantly imputed as 261 additional features (i.e. columns of the input matrix) in the imputation procedure. We randomly removed a subset of the observed lab results, including other labs that are ordered as part of the same lab panel(s), and imputed the masked values using the remaining observed values. The imputed values were then compared to the held-out, masked values to assess the quality of the imputation. In Figure 2, we show the imputation accuracy (*R*^2^ between the masked true and imputed values) for labs where the addition of cross-validated MRS to the baseline SoftImpute procedure explained significantly more variability. Of the 67 lab results considered, 25 (37.3%) were significantly better imputed by including the MRS values. Including the MRS values led to a median increase of 47.6% (95% CI 17.3%-90.9%) in the imputation *R*^2^ values.

**Figure 2.**
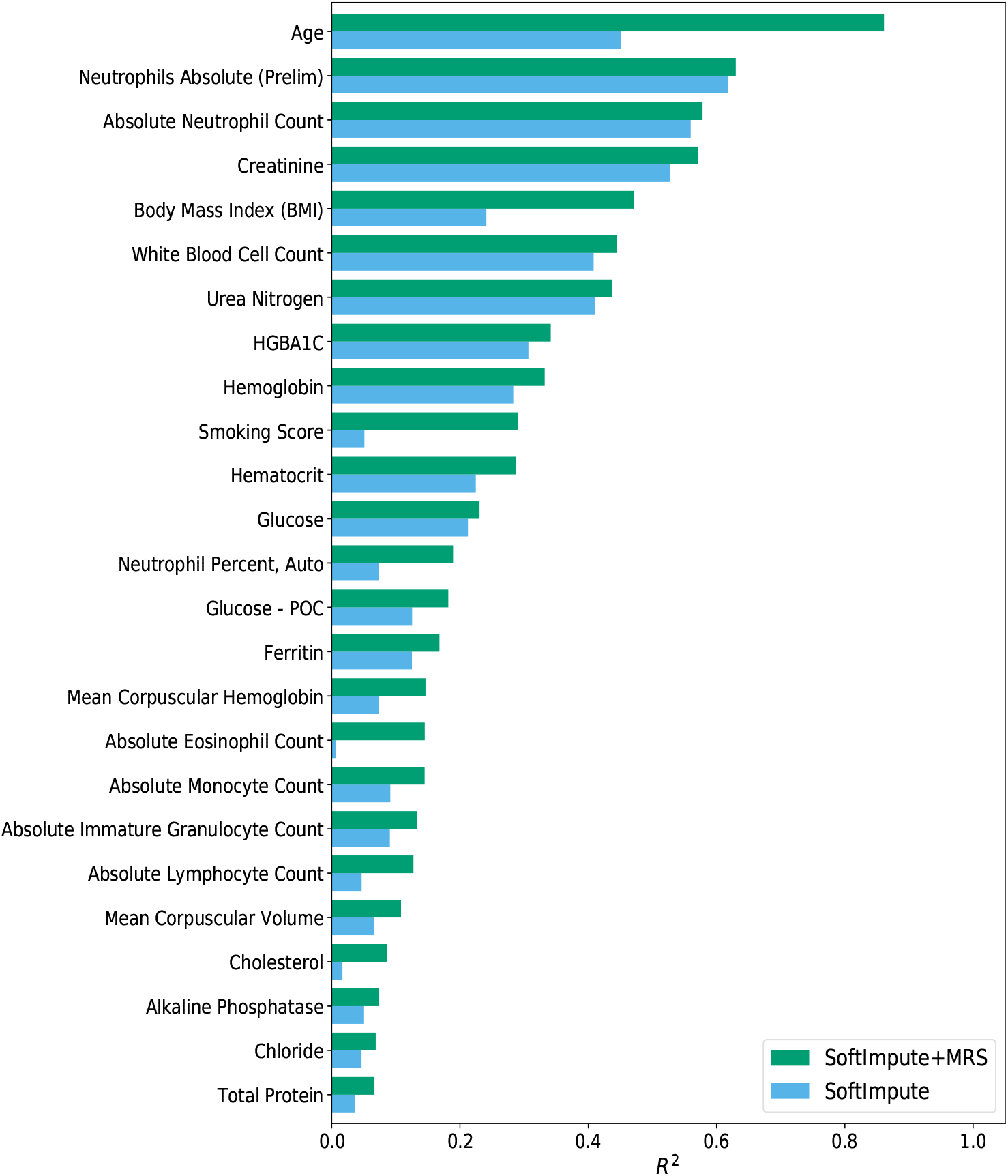
Improvement in lab result imputation performance by including MRS. For lab results that were significantly better imputed using a matrix completion imputation procedure that included the MRS values, we compare the quality of the imputed values (*R*^2^) using only the EHR data (SoftImpute) to the values generated when including the MRS values in addition to the EHR data (SoftImpute+MRS).

### Methylation risk scores will improve with larger sample sizes

In this study, our analyses of imputation accuracy were performed on 831 individuals’ methylation and genetic features. For many phenotypes, the genetic effects are relatively small and require large sample sizes to identify associations between genomic features and the outcome of interest. Consequently, in many biobanks the number of individuals with measured genomic features is several orders of magnitude larger than our sample size [1–3]. While the methylation data provided sufficient power to significantly impute numerous outcomes, there may remain much power to be gained by increasing the number of methylation samples to numbers approaching biobank-scale.

To determine the role of sample size in our imputation accuracy, we performed an experiment in which we downsampled the number of individuals in our data and trained models on the subsampled data. From the set of outcomes most accurately imputed by methylation and that also significantly improved the baseline’s imputation, we chose 10 medications, labs, and Phecodes on which to perform 10-fold cross-validation. For each sample size, we repeated the procedure 20 times to attempt to mitigate variance due to ascertainment effect. Though we selected features that had high accuracy using the full set of data, our results suggest that our models may become more accurate as the sample size increases (Figure 3; Figures S6,S7,S8). We further posit that there may be additional outcomes that will be significantly imputed as the number of methylation samples increases.

**Figure 3.**
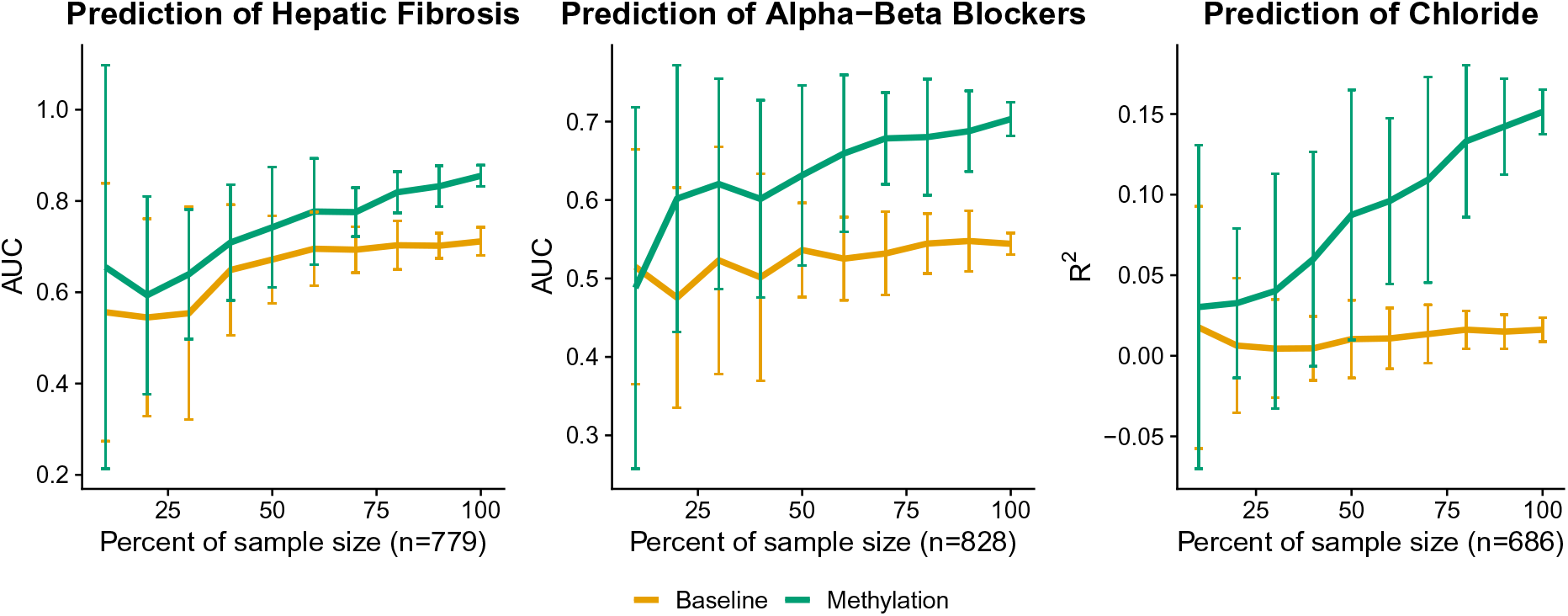
Imputation accuracy may improve with additional samples. We downsampled the number of individuals to evaluate the imputation performance as a function of sample size using a well-imputed medication, lab value, and Phecode. The performance is significantly affected by the number of individuals, suggesting that there is additional power to be gained with the addition of more methylation samples.

### Comparing MRS to UKBiobank PRS

As expected, due to a small sample size and the likely small effects of SNPs on phenotypes, the PRS developed using the UCLA cohort did not add substan-tial predictive power over the baseline features. Studies leveraging biobanks with sample sizes several magnitudes larger than the cohort at UCLA however, have shown non-zero heritability for a variety of phenotypes [1, 51–53]. Therefore, we sought to compare the MRS and PRS generated with the UCLA data to a polygenic risk score created using the UKBiobank data [54]. To do so, we obtained the genotype weights corresponding to 10 polygenic risk scores trained on the UKBiobank (Table S10) [52–55] data and imputed the external risk scores into our health record system using PLINK [56]. We included in the comparison labs that were significantly imputed by the baseline model and excluded labs that corresponded to cell counts or labs for which the internal PRS outperformed the external PRS (indicating a mismatch in the phenotypes or cryptic population structure that was unaccounted for by principal components). While the external polygenic risk score improved substantially the imputation performance relative to the internal polygenic risk score, it did not significantly outperform the methylation for any of the tested phenotypes (Figure 4). The methylation remained the best predictor in general—even when trained on fewer than 1000 samples—significantly outperforming the other models in the imputation of urea nitrogen, creatinine, hemoglobin, hematocrit, and albumin. The externally-derived polygenic risk score greatly outperformed both the internally-derived PRS and the MRS when predicting glycated hemoglobin (HGBA1C) and HDL levels, however the improvement was not significant. For detailed information on the external PRS and accession numbers, see Supplementary Table S10.

**Figure 4.**
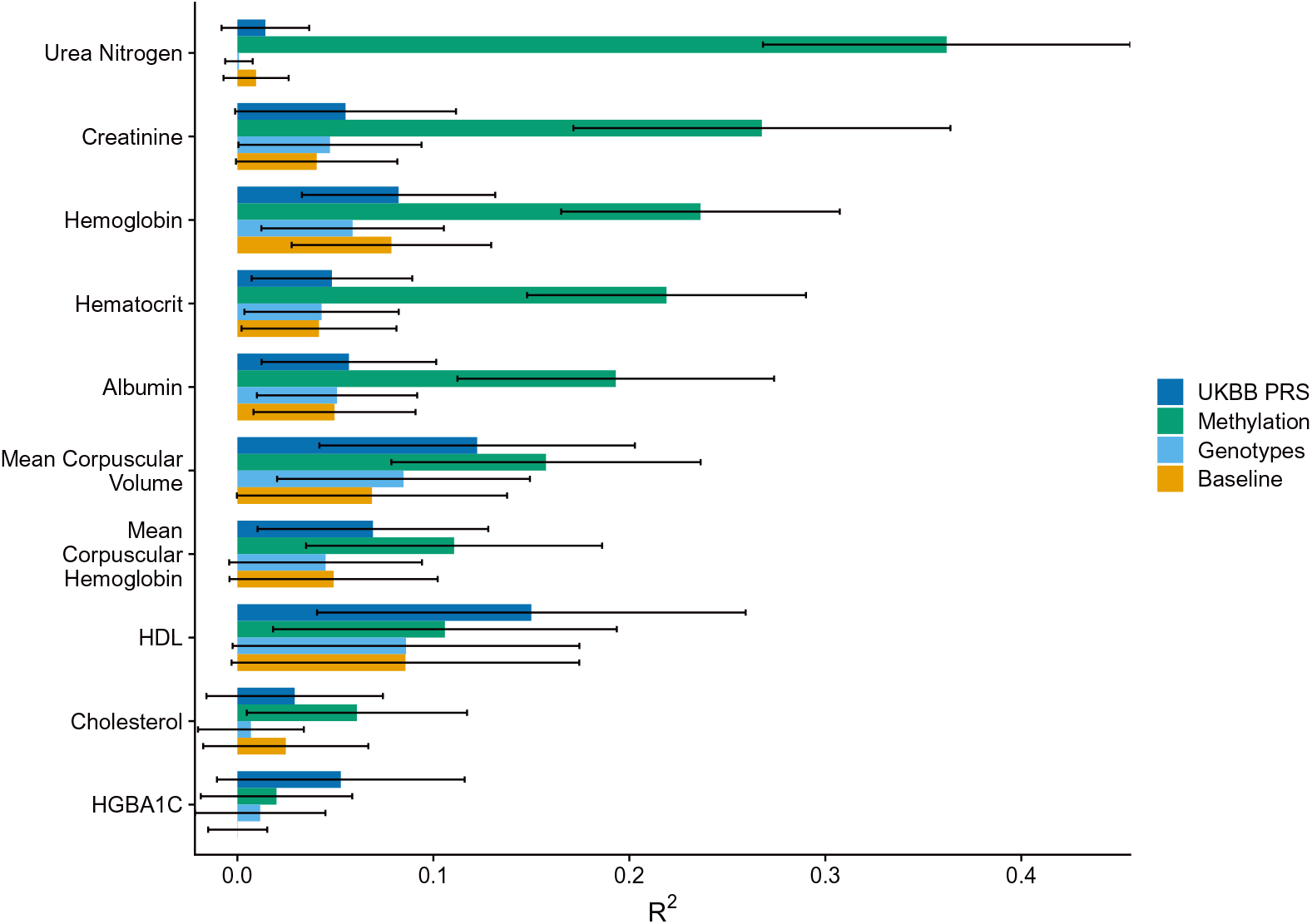
Labs as imputed by methylation, genotypes, and an externally-trained polygenic risk score. The cross-validated *R*^2^ between the true and imputed lab value on 541 unrelated patients of non-Hispanic-Latino white-identifying individuals using a baseline predictor as well as a baseline predictor with methylation, genotypes, and a PRS externally-trained from UKBiobank summary statistics. HDL corresponds to high-density lipoprotein cholesterol and HGBA1C to glycated hemoglobin.

Similarly to the analyses in which we examined whether predictors that leverage genomics offered predictive value over the baseline predictor, we examined whether our internal MRS and the externally trained PRS offer information that is complementary to the other. To do so, we measured the accuracy when using the MRS, external PRS, both risk scores, and as well as both risk scores and their interaction on the same set of labs as our original analysis. None of the models significantly outperformed the MRS alone (Figure S10). However, there was a significant interaction effect between the MRS and external PRS on creatinine (p=9.16e-05), as well as a nominally significant interaction effect on mean corpuscular hemoglobin (p=1.45e-02). As the interaction terms improved the accuracy for both outcomes, there may be added value in leveraging both MRS and PRS for imputation tasks, especially those that take advantage of non-linear effects.

### Evaluation of methylation risk scores across ancestral populations

Previous reports have suggested that a significant confounder to the application and versatility of polygenic risk scores is population structure, where a population-specific bias is induced that affects generalizability of PRS to different ancestries [57–59]. The collection of samples analyzed throughout this study is ethnically heterogeneous— individuals were self-identified as non-Hispanic/Latino European, Hispanic/Latino, Black, or Asian. Methylation data is also influenced by differences in population [60], and in particular the first several methylation principal components sufficiently capture population structure in European and African groups [61, 62]). Consequently, we examined the performance of the methylation risk scores within and across ancestral populations.

Primarily, after training the models on the entire heterogeneous set of samples, we examined the predictive performance within each ancestral population. When we examined the top 10 best-imputed (by MRS across the entire set of individuals) lab panels, medications, and Phecodes, only 10 of the entire 180 possible comparisons (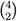 comparisons across 30 outcomes) displayed significant differences between the predictive performance within each population separately (Figures 5, S11, S12).

**Figure 5.**
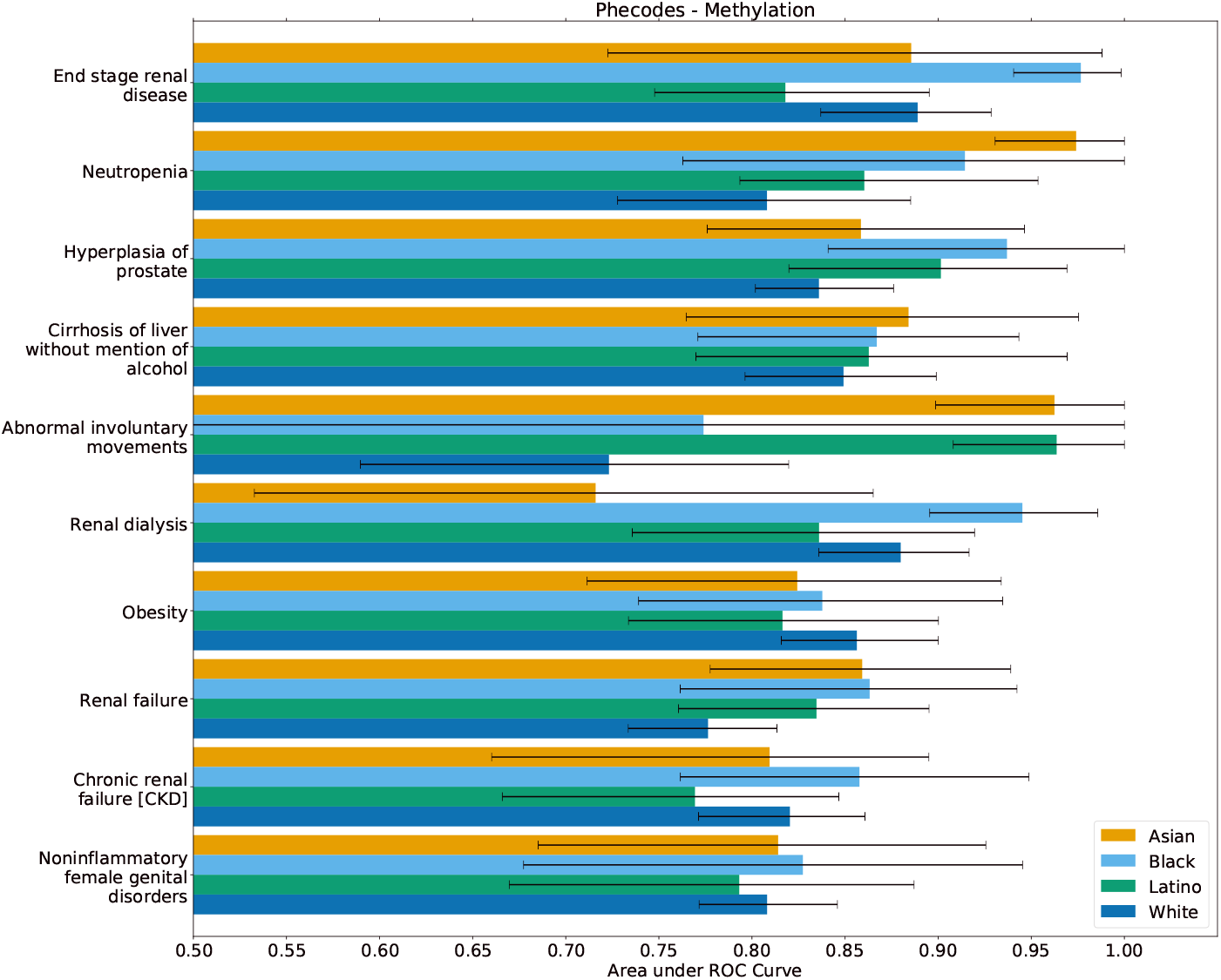
Best methylation-imputed Phecodes within ancestral populations. After training a model on the entire heterogeneous population of individuals, we evaluated the predictive performance within each population separately. We observed only 6 (of 60) significant differences between self-reported ancestral groupings.

In a second replication analysis we trained predictive models within ancestral groupings separately. As the individuals self-identified as either Black or Asian comprised less than 100 individuals in both groupings, we focused our analyses on Hispanic/Latino- and white-non-Hispanic/Latino-identifying individuals. We retrained models for the top 5 best-imputed (by MRS) medications, lab panels, and unique Phecodes on the Hispanic/Latino individuals and white non-Hispanic/Latino individuals alone and treated a prediction as significant if its association p-value was lower than .01. Creatinine, hemoglobin, and urea nitrogen replicated across both groupings, however, hematocrit and mean corpuscular hemoglobin did not replicate in the Latino/Hispanic grouping (Table 1). In the context of medications, CMV agents, osmotic diuretics, phosphate binder agents, hematopoietic growth factors, and immunosuppressive agents replicated within the white non-Hispanic/Latino population but only CMV and immunosuppressive agents replicated within the Hispanic/Latino population (Table 1). Finally, Phecodes corresponding to immunity deficiency, hypertensive renal disease and end-stage renal failure replicated within both groupings, however, neutropenia and anemia replicated only within the white non-Hispanic/Latino set of individuals (Table 1).

**Table 1.**
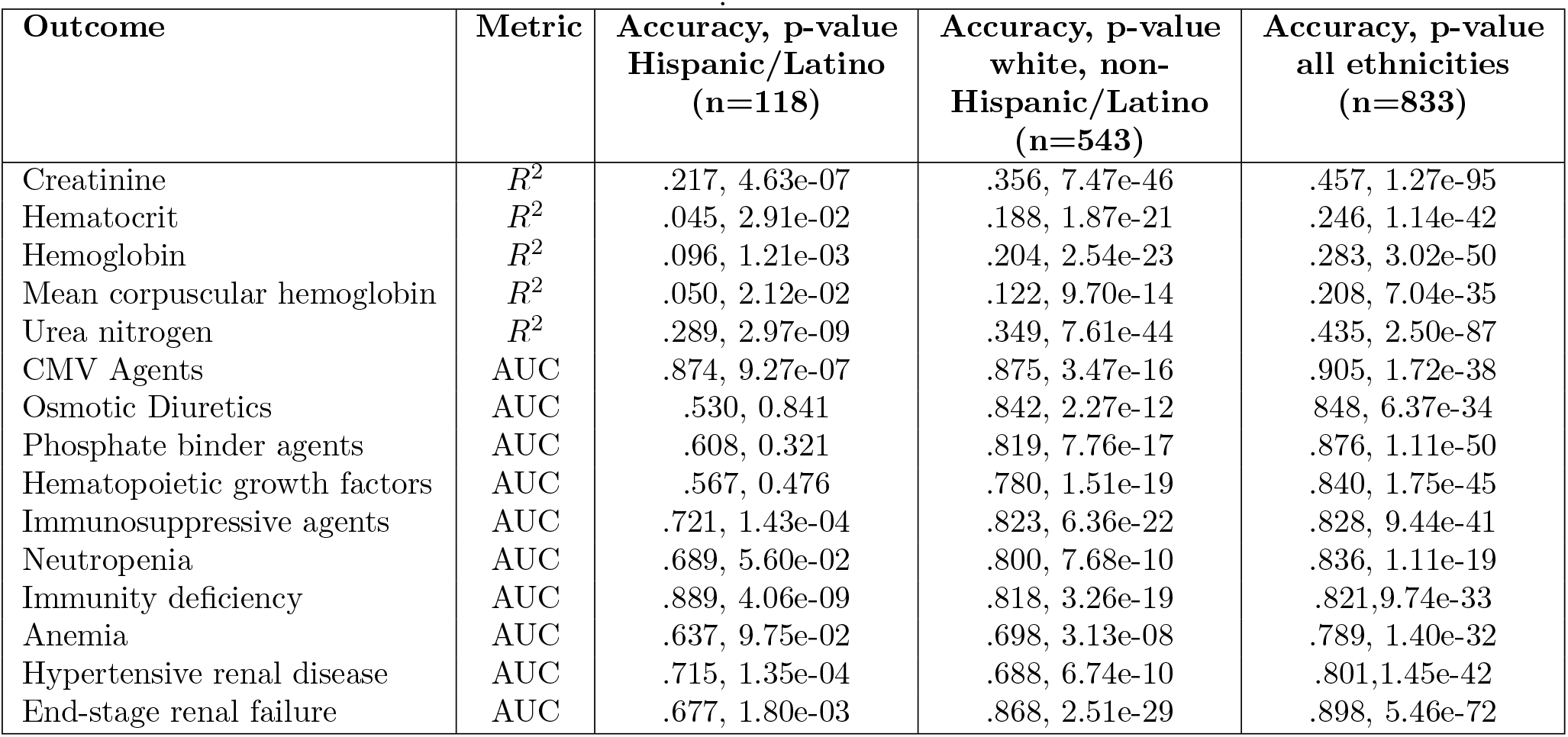
Replication statistics within ethnic groupings. Predictive accuracy (*R*^2^ and AUC) for MRS trained within only Latino/Hispanic-or white-non-Latino/Hispanic-identifying individuals compared to the accuracy trained on the entire, cross-ethnic cohort.

### Replication of methylation risk scores across external datasets

To evaluate the transferability of the MRS to a different population, we performed several experiments in which we imputed the MRS into external datasets. Primarily, we focused on imputation of kidney-related outcomes as they were the most accurately imputed in our own cohort. To do so, we leveraged a dataset that used the Human-Methylation27k array to measure the methylation of 194 individuals who had Type 1 Diabetes, 49.7% of whom had nephropathy (cases) [63]. We re-trained the models for a Phecode corresponding chronic renal disease as well as labs corresponding to creatinine and urea nitrogen on our in-house data, limiting our analysis to the 27,000 sites that belonged to the external dataset. The imputed chronic renal disease was significantly associated with nephropathy in the external dataset (p=8.32e-05, AUC=.684 [.615,.758]. Further, both of the imputed values for creatinine and urea nitrogen were significantly associated with nephropathy (p=5.11e-07, AUC=.739 [.670,.808] and p=3.71e-05 AUC=.693 [.619,.767], respectively). Importantly, when limiting our internal analysis to sites only on the 27k array, the association signal decreased (for chronic renal disease from p=6.81e-51 to p=3.13e-29, creatinine p=1.27e-95 to p=3.14e-62, and urea nitrogen p=2.50e-87 to p=8.44e-34). However, likely due to correlation between CpGs, the association tests for outcomes trained on the smaller set of sites were still significant.

Second, we expanded our replication analyses to include phenotypes that were unrelated to kidney function. In these analyses, we revisited epigenome-wide association studies (EWAS) of Schizophrenia [64] and Rheumatoid Arthritis [65] and imputed commonly prescribed medications for each dataset—for Schizophrenia we used phenothiazines, and for Rheumatoid Arthritis we used glucocorticosteroids. To ensure our MRS captured medication intake status and were not merely serving as proxies for the disease, we re-trained our models while conditioning on the trait of interest. The imputed phenothaizine intake was significantly associated with Schizophrenia case-control status (p=8.71e-04, AUC=.568 [.527,.611]) and the imputed glucocorticosteroids usage was significantly associated with Rheumatoid Arthritis case-control status (p=2.72e-07, AUC=.626 [.584,.669]. Weights for both medications were trained on CpGs corresponding to those present on the HumanMethylation450k array and also included their corresponding disease in the baseline set of covariates. Accordingly, the association signal of phenothiazines dropped from 1.14e-07 to 3.99e-05 and the performance of glucocorticoids dropped from 1.35e-16 to 1.82e-15 when compared to the MRS trained on the set of EPIC array CpGs and with the baseline features as covariates.

## 3 Methods

### Electronic Health Record Data

De-identified electronic health record data for this study was extracted from the perioperative data warehouse (PDW), a custom-built, robust data warehouse containing all patients who have undergone surgery at UCLA Health since the implementation of UCLA’s EMR (EPIC Systems, Madison, WI, USA) in March 2013. The PDW, which has been described previously [66], has a two-stage design. First, data are extracted from EPIC’s Clarity database into 29 tables organised around three distinct concepts: patients, surgical procedures, and health system encounters. Then, these data are used to populate a series of 4000 distinct measures and metrics such as procedure duration, admission ICD codes, lab results, and medication orders.

### Patient Ascertainment

Methylation and genotype samples were collected using blood from 831 patients as part of the UCLA ATLAS precision health initiative between October 26, 2016 and December 10, 2018 [67]. The samples were collected from patients before undergoing surgery with general anesthesia at UCLA Health, and the patients had not undergone surgery in the 30 days prior to blood sample collection. Of these patients, 302 were selected for inclusion based on the presence of acute kidney injury (AKI), defined as an Acute Kidney Injury Network (AKIN) classification of one or greater, after undergoing surgery. An additional 348 patients were risk-matched controls, with either glomerular filtration rate (GFR) less than or equal to 38 (210 patients), or GFR greater than 38 and a propensity risk score that matched case patients (348 patients). The propensity score was created using available EHR features such as age, weight, BMI, and other preoperative features that were measured in the hospital. Within the control group, we also performed a similar procedure ascertained on whether individuals were a heart attack case. Controls for heart attack patients were also selected using propensity scoring. Demographics of the patient population are further described in Table S1.

### Medication Usage

For each medication, a patient was labeled as using a medication if the electronic health record contained a medication order that occurred before the methylation sample collection date. Medications were grouped by pharmaceutical subclass using the Generic Product Identifier (GPI) hierarchical classification system codes. Any medications that were ordered in fewer than 5% of the patients were excluded from the analysis. In total, 168 pharmaceutical subclasses were considered in our analysis. The number of patients using medications from each subclass is shown in Supplemental Table S8. In Supplemental Table S9, we show for each pharmaceutical subclass the specific medication that patients in our cohort received.

### Lab Results

The most recent lab result prior to the methylation sample collection was extracted from the PDW for each patient. Any labs with a result date that occurred more than 365 days before the methylation sample collection date were excluded from the analyses. Additionally, labs for which there were less than 50 patients with valid results were excluded. We were left with a total of 69 lab values on which to run our models.

### Diagnosis Codes

International Classification of Diseases, Ninth Revision (ICD-9) and International Classification of Diseases, Tenth Revision (ICD-10) codes are a standard set of diagnosis codes, primarily used for billing purposes. While these codes provide a standardized methodology for describing a diagnosis, they are very specific. To map these specific diagnosis codes into meaningful, distinct diseases/traits, Denny et al. aggregated the ICD codes into phenotype codes (Phecodes) [43, 44]. Specifically, for each patient, we queried all diagnoses prior to the methylation sample collection date, and used the Phecode (version 1.2) mapping to aggregate ICD-9 and ICD-10 codes to unique, meaningful phenotypes. If a patient’s diagnosis record had both ICD-9 and ICD-10 labels, the ICD-10 to Phecode mapping was used instead of the ICD-9 to Phecode mapping. Each Phecode was treated as a binary variable, indicating the presence or absence of a relevant diagnosis code at any point in time before sample collection. We excluded rare Phecodes (occurrence less than 5% of the patients) and, in total, our cohort contained 370 unique Phecode phenotypes.

### Preprocessing of genotype data for cross-validation

We measured the genotypes for 831 individuals based on their DNA sampled from whole blood using the ATLAS genotype array. We preprocessed the genotype data using Beagle (d20) [68], PLINK (1.07) [56], and GCTA (1.93.2) [69]. We restricted the genotypes to autosomal variants, removed rare variants (MAF *< .*05), and filtered for variants that met Hardy-Weinberg equilibrium with p-value threshold 10*^−^*^6^. We also removed individuals and variants with more than 1% missing values. For the purpose of running cross-validation, we used Beagle to impute only any remaining missing values, but did not impute to an external dataset. We show that with our sample size and phenotypes evaluated, using genotypes imputed to an external reference does not significantly improve our results (Figure S14). In total we were left with 292,808 SNPs. To obtain principal components, we ran PCA using plink on the chipped genotypes.

### Preprocessing and imputation of genotype data for comparison to external models

We used a version of the ATLAS genotype data that was imputed to an external dataset, as detailed in [67]. Briefly, after performing quality control, genotypes were uploaded the Michigan Imputation Server [70]. The server phases the genotype data using Eagle v2.4 [71] and performs imputation using the TOPMed Freeze5 imputation panel [72] using minimac4[73]. We applied the same quality control and filters to the imputed genotypes as we did the chipped genotypes, and we were left with a total of 5,574,956 SNPs.

### Preprocessing of methylation array data

We measured methylation data for 831 individuals based on their DNA sampled from whole blood using the EPIC Illumina array. To generate beta-normalized methylation levels at each CpG, we ran the default pipeline of ENmix (1.22.0) [74] on the the raw probe data (IDAT files), which performs background correction, RELIC dye bias correction, and RCP probe-type bias adjustment. We removed from our analysis CpGs that coincided with SNP loci as well as CpGs on the sex chromosome. We also filtered out outlier samples, defined as having a PC score more than 4 standard deviations away from the average PC score in the first two principal components. In the imputation tasks, we removed sites with low variability (standard deviation *<* 0.02) leading to a total of 269,471 sites.

### Imputation using baseline medical features

To establish a baseline level of imputation performance, we constructed a set of features derived from basic patient information. We trained a simple linear (or logistic) model with 10-fold cross validation using an intercept and patients’ age, sex, BMI, methylation-based cell-type proportions (from the reference-based method of Houseman et al. [42]), self-reported ancestry, first ten genetic principal components, and smoking status (never, former or current). Importantly, we wished to establish how well an outcome (medication, Phecode, or lab value) could be imputed by using covariates (e.g. ancestry, age, smoking status) that are known to be captured by genomics.

### Imputation using a single penalized linear model

After establishing a baseline level of imputation performance, we performed penalized logistic and linear regression using either individuals’ methylation, genotypes, or both. More concretely, we fit 10-fold cross-validation using LASSO, elastic net and ridge regularization under the following two models:

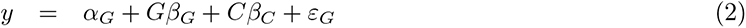

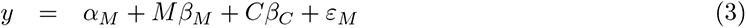

where *y* corresponds to the outcome, *α* the model-specific intercept, *G* the *n × s* genotypes, *M* the *n × c* methylation data, *β* the vector of length-*s* or -*c* effect sizes for the given explanatory variable, *C* and *β_C_* the covariates from the baseline model and their corresponding effect sizes, and *ε* the length *n* noise vector. We refer to models (2) and (3) as the PRS and MRS respectively, and note that they also include the baseline features. After fitting all three penalized linear models for a given datatype and outcome, we selected a final model as determined by the model with the highest cross-validated metric (AUC or *R*^2^ if the outcome was binary or continuous, respectively). We fit all penalized models using package *bigstatsr* [41]. We share MRS weights for outcomes that were significantly imputed at https://github.com/cozygene/EHR MRS UCLA. We also include details on the number of CpGs selected for each MRS in these analyses in Supplemental Tables S5, S6, and S7.

### Imputing lab results using EHR data and MRS values with softImpute

Imputing a partially-observed matrix of values is often formulated as a matrix-completion problem. In a matrix completion problem, the observed values of the matrix are used to estimate the values of the unobserved values by assuming that there is some underlying structure that is responsible for generating the data. For example, in the popular SoftImpute method [39], the data is assumed to be well-approximated by a low-rank representation, and the error between the observed values and the reconstructed values is minimized through a convex optimization procedure. However, since the unobserved values are, by definition, not observed, and therefore cannot be used to assess the imputation performance, the primary method for measuring the performance involves masking (removing) observed values and comparing the imputed values to these held-out, true values.

The EHR data used in the imputation procedure included demographic information, diagnosis codes, medication usage, and lab results, which were extracted from the EHR database using the previously described criteria. In addition to the EHR data, we also ran the imputation procedure while including relevant MRS values. Specifically, we included the MRS values for demographics, diagnosis codes, medication usage, and lab results that were imputed at a statistically significant level. These MRS values were added as additional observed features to the EHR matrix.

To estimate the imputation performance, we randomly masked 10% of the observed lab result values, and performed the imputation procedure (SoftImpute matrix completion) to generate estimates of the missing values. However, since labs are most often ordered in panels, for example a metabolic panel, if a lab is missing then typically other labs that are part of the same panel are also missing. We simulated a more realistic missingness scenario by, instead of masking out values only from a specific lab *l*, masking out all labs that are ordered as a panel that include lab *l*. This masking procedure was done per lab, using 10-fold cross-validation, such that 10% of the non-missing values of a particular lab result (and its associated lab panels) were masked (removed), and the remaining 90% of the observed values were used to complete the matrix. Matrix completion was performed using the SoftImpute algorithm, as implemented in the *fancyimpute* [75] python package (version 0.5.5). The proportion of variance explained (*R*^2^) of the true lab values by the imputed lab values was used to measure the imputation performance. Confidence intervals were derived using bootstrapping.

### Hypothesis testing

To determine whether an imputation was significant or whether one predictor offered significant additional explanatory signal, we conducted our hypothesis tests using a linear (logistic) regression framework. Primarily, after running cross-validation or generating a single predictor 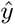 for an outcome *y*, we would test whether the imputation was significant by comparing it to *y*:

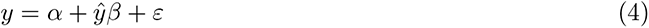

Where Equation (4) corresponded to linear regression when the outcome was continuous, and logistic regression when the outcome was binary, *α* was the intercept, and *β* was an effect size indicating association of the predictor with the outcome. Notably, by building our testing framework as a linear model, we can easily extend it to include additional predictors in order to test whether the additional predictors significantly improve the fit of the regression—or more simply, whether predictor 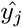 offers additional predictive power over 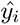 by conducting a likelihood ratio test of the following nested models:

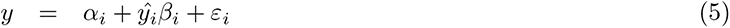

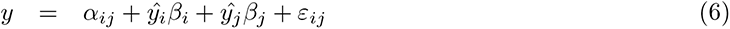

Where *i* and *j* index either the baseline, MRS, or PRS models. We corrected for multiple hypothesis tests within each outcome and method by using a Bonferroni adjustment at *α* level .05.

### Imputing external polygenic risk scores into the ATLAS cohort

We compared our in-house built risk scores to risk scores learned in the UKBiobank dataset[51, 53]. In both [51, 53] the authors construct their PRS using penalized regression akin to as we have done in our analyses. Notably, using penalized regression on individual-level genotypes allows one to automatically, optimally control for shrinkage and variable selection at the step of model generation[41, 76]. This is in contrast with many commonly used polygenic risk score tools such as LDPred[77] or PRSCS[78], that attempt to perform shrinkage or variable selection post-hoc on the level of summary statistics. After downloading the PRS from the PGS catalog[55] listed in Suppleplementary Table S10, we imputed PRS into our cohort using our imputed genotypes using the score function of Plink. To account for population structure, we limited our analysis to individuals who self-identified as white, and passed filtering using manual inspection of principal components (Figure S9).

### Ethical approval and patient consent

Retrospective data collection and analysis was approved by the UCLA IRB. All research was conducted in accordance with the tenets set forth in the Declaration of Helsinki.

### Data availability

The MRS weights for outcomes that were significantly imputed are located at: https://github.com/cozygene/EHR MRS UCLA. The raw UCLA datasets generated during and/or analyzed during the current study are not publicly available due to institutional restrictions on data sharing and privacy concerns.

## 4 Discussion

In this study, we provide a comprehensive investigation of the utility of methylation risk scores in a clinical setting. We used (to our knowledge) the largest methylation biobank cohort produced to date, which includes methylation, genotype, and comprehensive EHR data for all patients. We find that the MRS improved imputation performance over a baseline model by 10.65%, 156.31%, and 14.59% when predicting medication usage, lab panel values, and diagnosis codes respectively. These contributions are significantly more substantial than those obtained by PRS.

The vision of genomic biobanks is that the genomic data will be translated into improved clinical diagnosis and treatment decisions [12, 13, 79]. In practice, clinical decisions are not expected to be based solely on genomic information, but rather on the combination of the genomic, medical, and demographic information of the patient. While previous studies have used a limited number of key features as a baseline for imputation of a phenotype (e.g., age, sex, and major comorbidities) [50, 80–82], to the best of our knowledge, these studies did not take into account the entire familial-genetic or environmental history of the patients. Thus, the question of whether genomic data (methylation or genetics) can be used to improve imputation over the EHR data is critical in order to claim clinical relevance. Our results demonstrate that adding MRS to existing EHR-based imputation frameworks improve imputation accuracy by over 29% in a clinical context.

It is well appreciated that PRS are sensitive to the studied population, and it is often the case that a PRS developed for one ethnic group performs poorly on others [57, 59]. It is therefore important to evaluate the population effect on MRS performance. For this reason, we measured the transferability of our results across different populations, and we observe that the accuracy of the MRS was robust to population structure. This is likely driven by the diversity of the training cohort used, but also due to the fact the we are under-powered to discover subtle differences in imputation accuracy due to our sample sizes. Nonetheless, since we observed very few large differences in accuracy across populations, we are hopeful that our results will inspire future investigations to continue to recruit diverse cohorts and to examine these differences at length with greater sample sizes.

While our study was focused on methylation, there are many other possibilities for the introduction of genomic data in clinical settings. First and foremost, genetic data has been heavily studied by others and large biobanks including genetic data of patients already exists. However, other measurements such as RNA, microbiome, metabolomics, or proteomics may also be relevant. Some of these have logistic and cost considerations at scale. One of the advantages of methylation is that DNA biobanks already exist in large numbers, and the cost of measuring methylation is close to that of measuring genetic data. Moreover, different genomic measurements may provide different snapshots of the patient’s data, risk, or health status. Methylation, for example, is known to capture one’s smoking status[26], and may therefore be particularly useful for cases in which researchers intend to use self-reported features that may suffer from patient recall bias or honesty. Tangentially, while polygenic risk scores provide a lifetime risk for a patient, methylation risk scores may provide the current risk of the patient over the last few months [83–85], and other genomic information may provide risk with the resolution of days or hours (e.g., RNA or certain metabolomics [86–89]). Nonetheless, owing to the dynamic nature of methylation, it is currently unclear what the range or duration of the methylation risk score are. Furthermore, while methylation patterns are associated with outcomes, it is generally unknown if they cause a disease or are a response to a disease [90].

To assist the research community in investigating methylation in the context of disease, we provide the MRS predictors for all significantly predicted outcomes at https://github.com/cozygene/EHR MRS UCLA. While our samples were ascertained on kidney and heart disease, we show that our weights successfully replicated across three internal datasets, including studies of Rheumatoid Arthritis and Schizophrenia.

Consequently, our weights may be used by researchers and clinicians in different ways. For example, in many epigenome-wide association studies (EWAS), in which associations between specific methylation CpG sites and a phenotype are studied, one may wish to account for patients’ comorbidities and medications, which are often not available to the study. Using the MRS database, the researchers leveraging EWAS will be able to incorporate such covariates into their model.

There are multiple potential next steps for the examination of methylation in clinical contexts. First, in this work we focused our attention on the imputation of the phenotypes, or in other words, the inference as to whether the patient is currently diagnosed with a disease. We hope that our findings will be able to be translated to the inference of future clinical events, i.e., prediction of future deterioration or disease occurrence. Second, our analyses did not focus on generating models for a specific patient demographic (e.g. only senior patients) and we were limited to methylation collected from blood samples. As methylation is known to vary across age and tissue type, models may be improved by focusing on individuals of a specific demographic, or by assaying a tissue relevant for a given phenotype (e.g. liver tissue for metabolic disorders. Third, although our evaluation is across the largest dataset which includes both EHR, methylation, and genotype data, the sample size of our study is still moderate compared to genetic studies that are performed on biobanks. Indeed, we demonstrate that for some of the phenotypes, an increase in sample size will likely lead to a substantially improved imputation accuracy (Figure 3). Moreover, larger sample size data may be able to reveal the quantity or contribution of genetics verses methylation to the MRS imputation accuracy [49]. In light of our results, as well as the fact that many biobanks have already obtained blood or DNA samples, we recommend that future biobanks consider measuring methylation in addition to the genotypes across a large number of patients.

### Competing Interests

I.H. is the president of Clarity Healthcare Analytics Inc, a company that assists hospitals with extracting and using data from their electronic medical records. The company currently owns the rights to the PDW software that was used to extract data from the electronic health record. I.H. receives research funding from Merck Pharmaceuticals. M.C. is a consultant for Edwards Lifesciences (Irvine, CA) and Masimo Corp (Irvine, CA), and has funded research from Edwards Lifesciences and Masimo Corp. He is also the founder of Sironis and he owns patents and receives royalties for closed loop hemodynamic management technologies that have been licensed to Edwards Lifesciences. E.H. is senior vice president of AI/ML at OptumLabs (Minnetonka, MN). The other authors declare no competing interests concerning this article.

### Author Contribution

IPH, E.H., N.Z., B.H., and M.T. conceived of study. M.T. and B.H. performed data analysis under significant contribution and input from S.S., N.R., J.N.C., M.C. and I.H. IPH, N.R., E.H., M.C., I.H., J.N.C. contributed to data acquisition and design. All authors contributed to manuscript writing.

## Data Availability

Methylation risk scores are available at https://github.com/cozygene/EHR_MRS_UCLA

---

**Figure S1.**
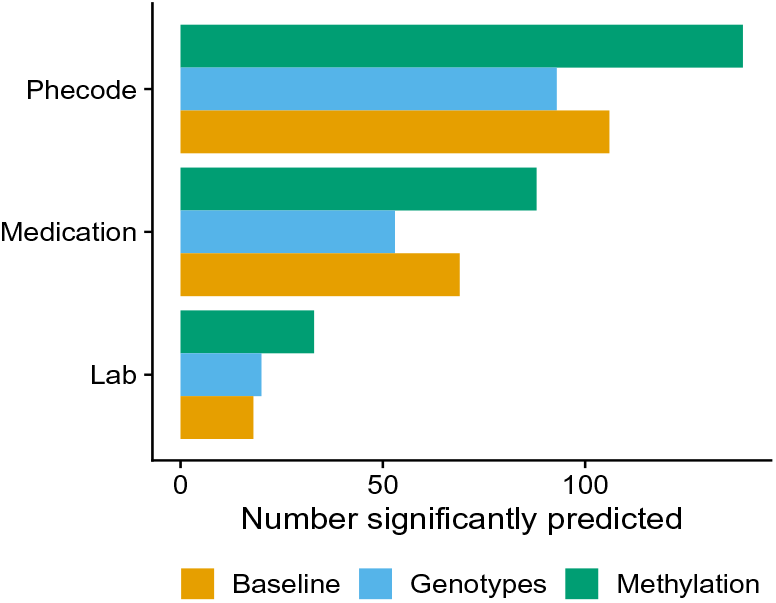
Significantly predicted outcomes per data type. Total number of significantly predicted outcomes when using the baseline alone, as well as including either set of genomic features in addition to the baseline. We used an association test of the cross-validated predictors and the true outcome and adjusted for multiple testing using Bonferroni correction at a nominal threshold of 0.05.

**Figure S2.**
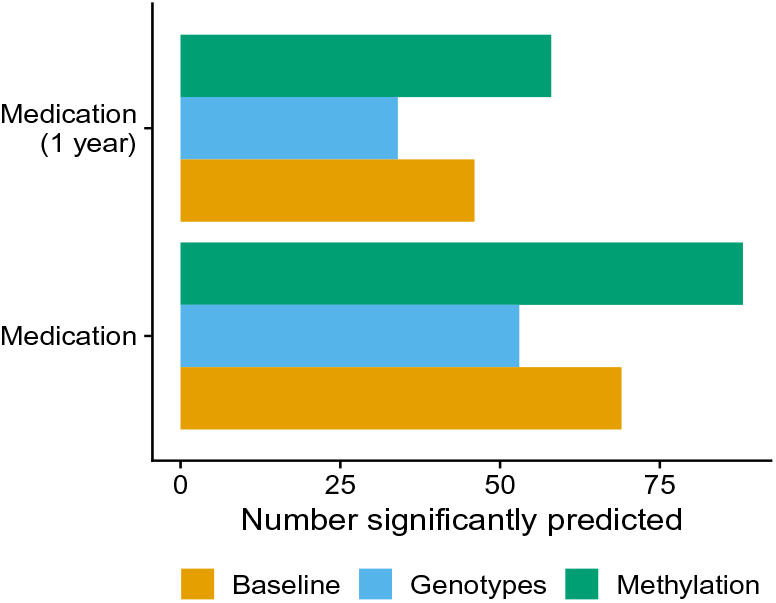
Significantly predicted medications prescribed within one year of collection date or EHR history. Total number of significantly predicted medications when using the baseline alone, as well as including either set of genomic features in addition to the baseline. We used an association test of the cross-validated predictors and the true outcome and adjusted for multiple testing using Bonferroni correction at a nominal threshold of 0.05. Using a patient’s entire EHR history to generate their list of medications resulted in more significant associations than using just the year prior to sample collection date.

**Figure S3.**
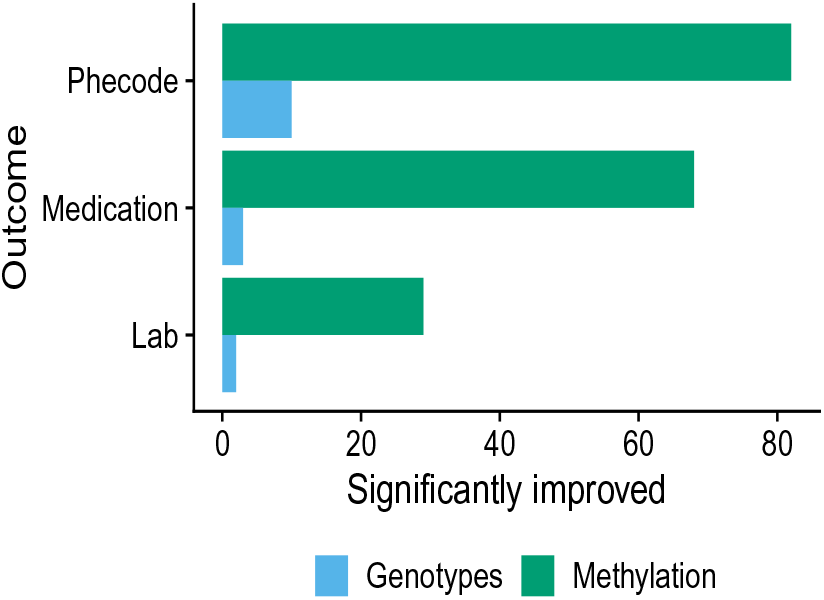
Number of outcomes significantly improved when adding the corresponding predictor. Similarly to the tests in which we surveyed whether the MRS or PRS predictors added predictive power over the baseline predictor, we conducted an analysis in which we examined whether the MRS predictor adds predictive power over the PRS predictor (Green “Methylation”), and whether the PRS predictor adds predictive power over the MRS predictor (Blue “Genotypes”). The MRS predictor improved the performance of the PRS predictor for a substantial number of outcomes

**Figure S4.**
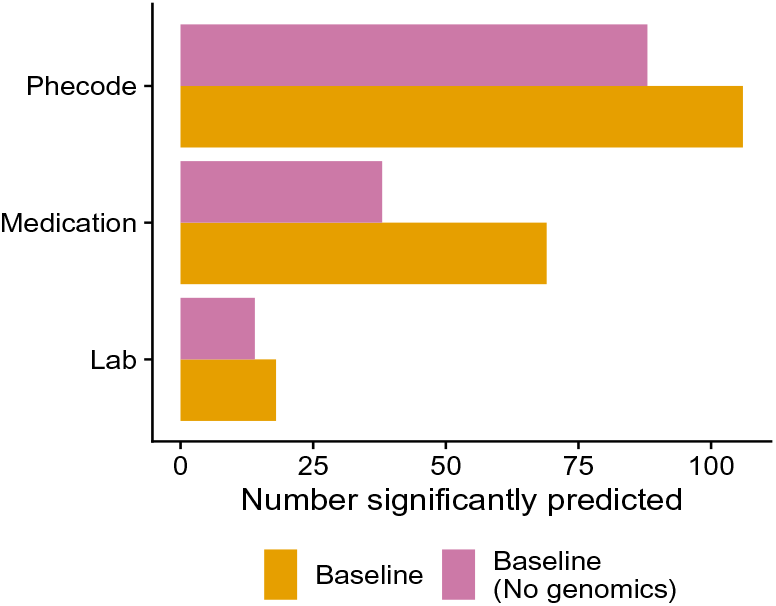
Significantly predicted outcomes across baseline models. Total number of significantly predicted outcomes when using the baseline model with and without variables derived from genomics (ancestry principal components, methylation-estimated cell-type composition estimates). We used an association test of the cross-validated predictors and the true outcome and adjusted for multiple testing using Bonferroni correction at a nominal threshold of 0.05.

**Figure S5.**
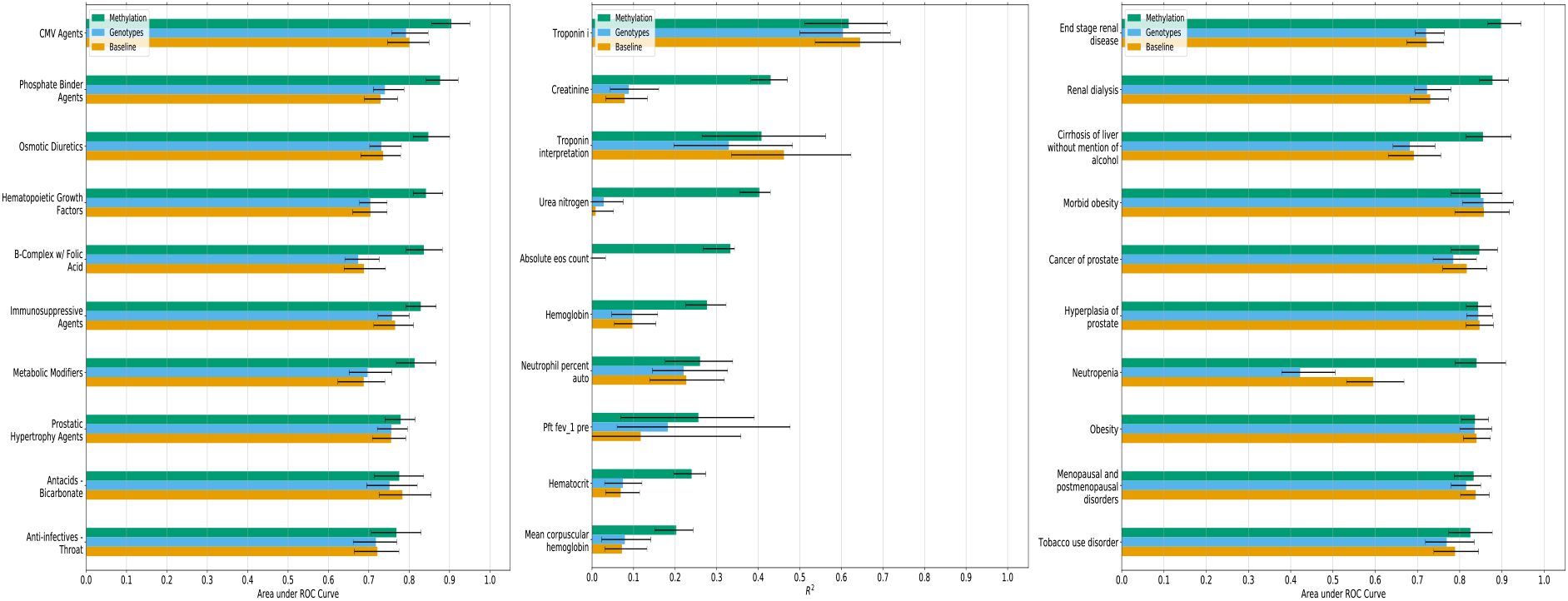
Significantly-predicted outcomes per data type The top 10 methylation-predicted (Left) medications, (Middle) labs, and (Right) Phecodes, with Baseline and Genotype prediction performance results for comparison.

**Table S1.**
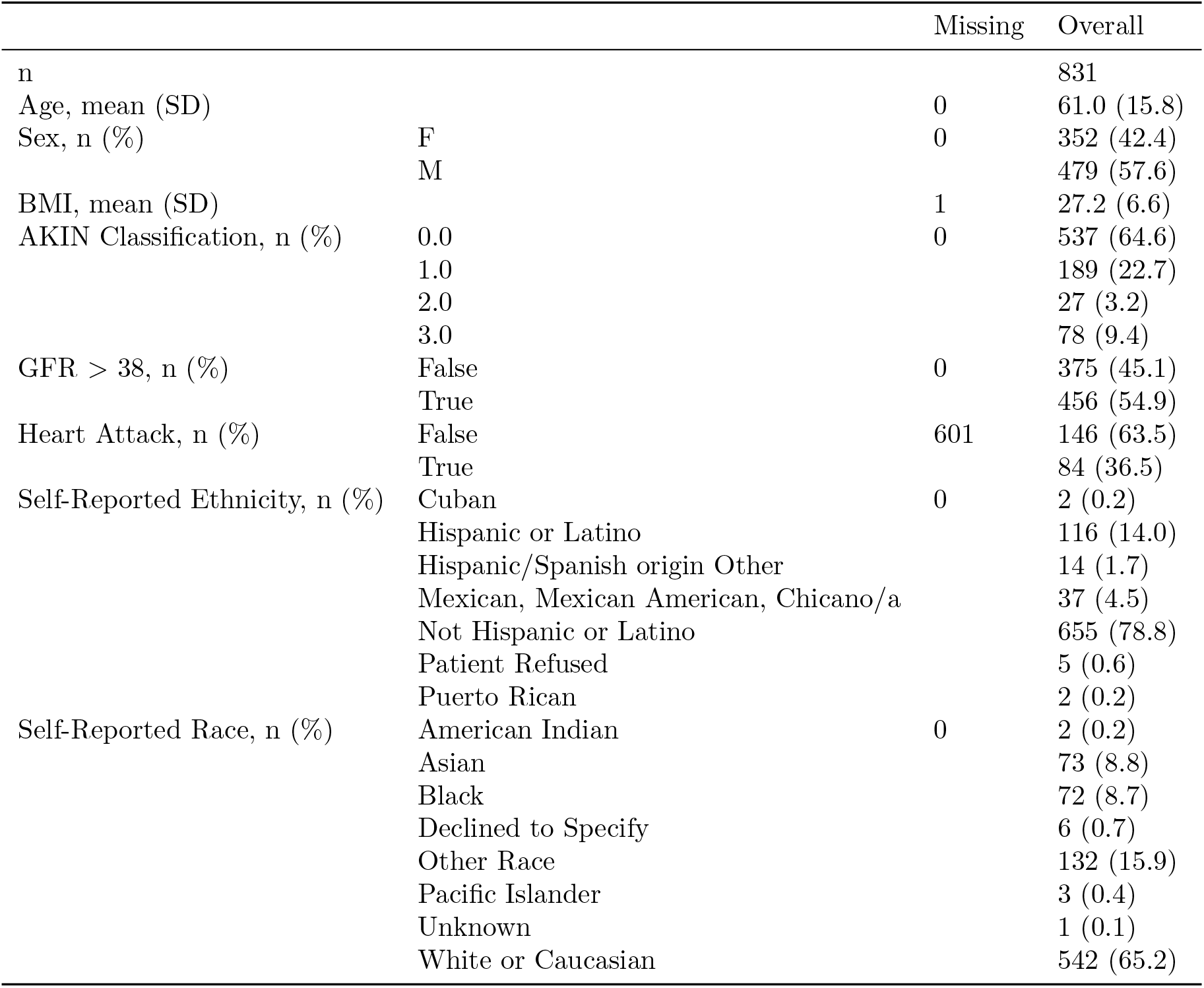
Cohort patient demographics. AKIN is the Acute Kidney Injury Network Classification, BMI is Body Mass Index, GFR is glomerular filtration rate.

|  |  | Missing | Overall |
| --- | --- | --- | --- |
| n |  |  | 831 |
| Age, mean (SD) |  | 0 | 61.0 (15.8) |
| Sex, n (%) | F | 0 | 352 (42.4) |
|  | M |  | 479 (57.6) |
| BMI, mean (SD) |  | 1 | 27.2 (6.6) |
| AKIN Classification, n (%) | 0.0 | 0 | 537 (64.6) |
|  | 1.0 |  | 189 (22.7) |
|  | 2.0 |  | 27 (3.2) |
|  | 3.0 |  | 78 (9.4) |
| GFR > 38, n (%) | False | 0 | 375 (45.1) |
|  | True |  | 456 (54.9) |
| Heart Attack, n (%) | False | 601 | 146 (63.5) |
|  | True |  | 84 (36.5) |
| Self-Reported Ethnicity, n (%) | Cuban | 0 | 2 (0.2) |
|  | Hispanic or Latino |  | 116 (14.0) |
|  | Hispanic/Spanish origin Other |  | 14 (1.7) |
|  | Mexican, Mexican American, Chicano/a |  | 37 (4.5) |
|  | Not Hispanic or Latino |  | 655 (78.8) |
|  | Patient Refused |  | 5 (0.6) |
|  | Puerto Rican |  | 2 (0.2) |
| Self-Reported Race, n (%) | American Indian | 0 | 2 (0.2) |
|  | Asian |  | 73 (8.8) |
|  | Black |  | 72 (8.7) |
|  | Declined to Specify |  | 6 (0.7) |
|  | Other Race |  | 132 (15.9) |
|  | Pacific Islander |  | 3 (0.4) |
|  | Unknown |  | 1 (0.1) |
|  | White or Caucasian |  | 542 (65.2) |

**Table S2.**
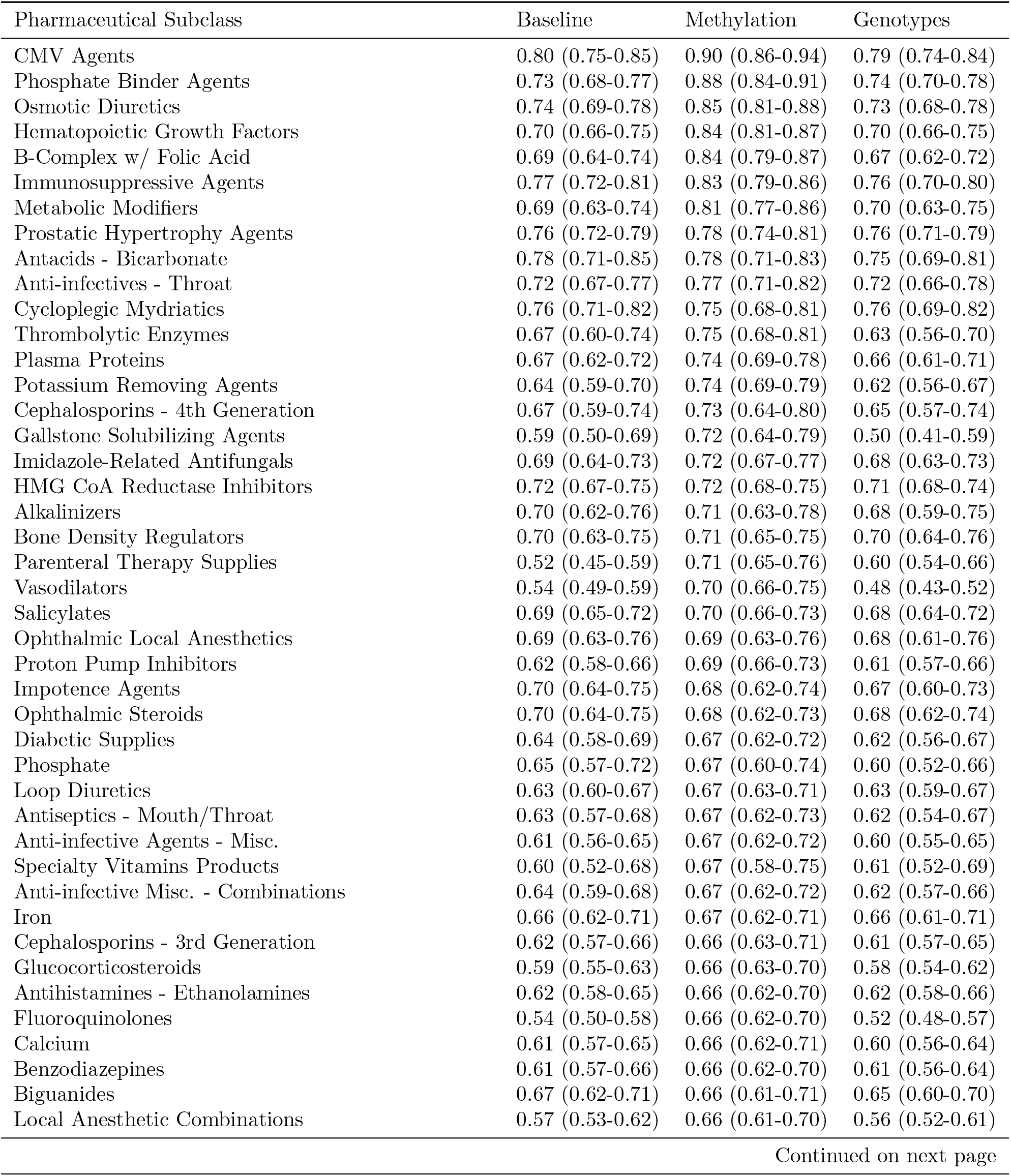
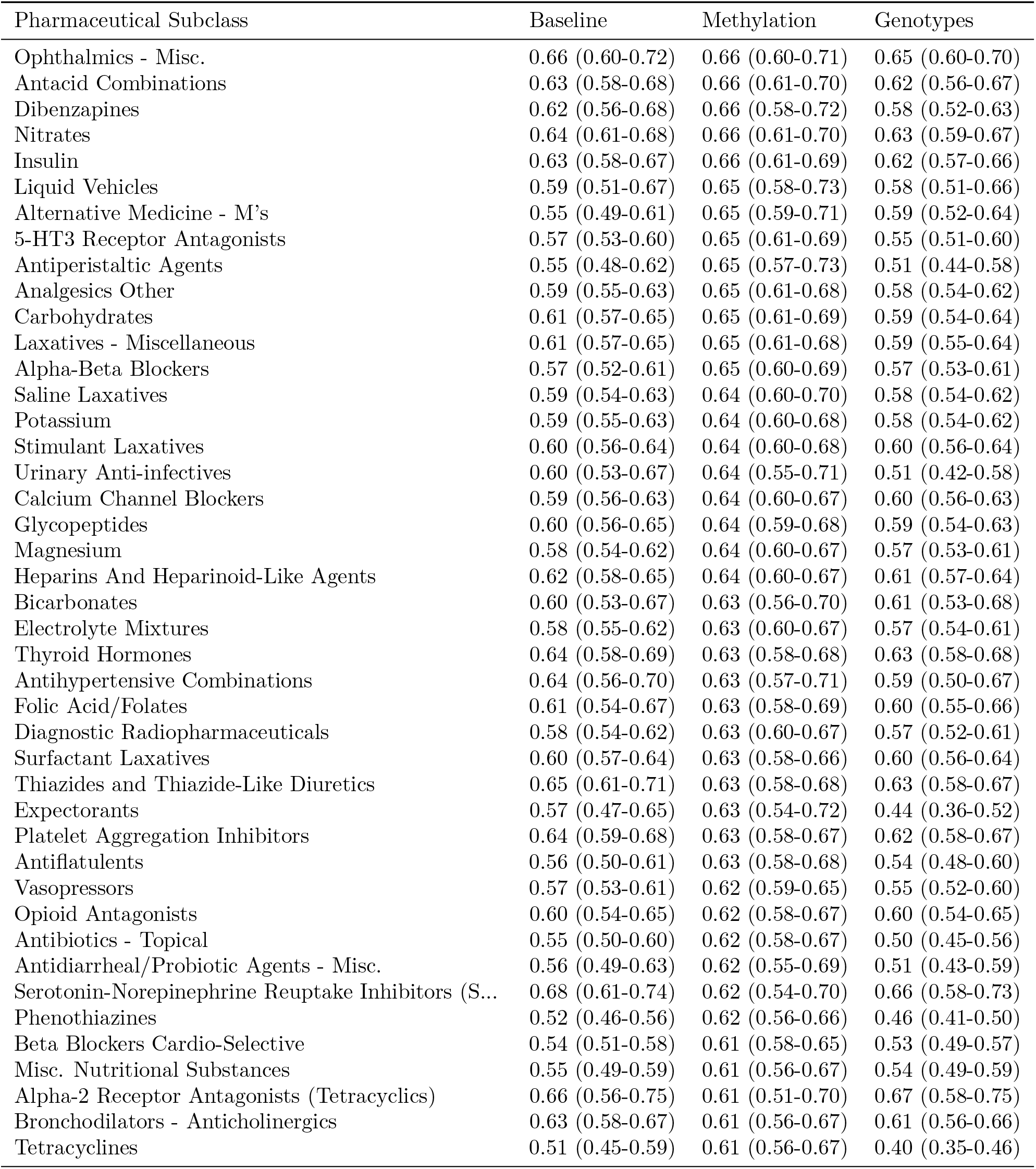
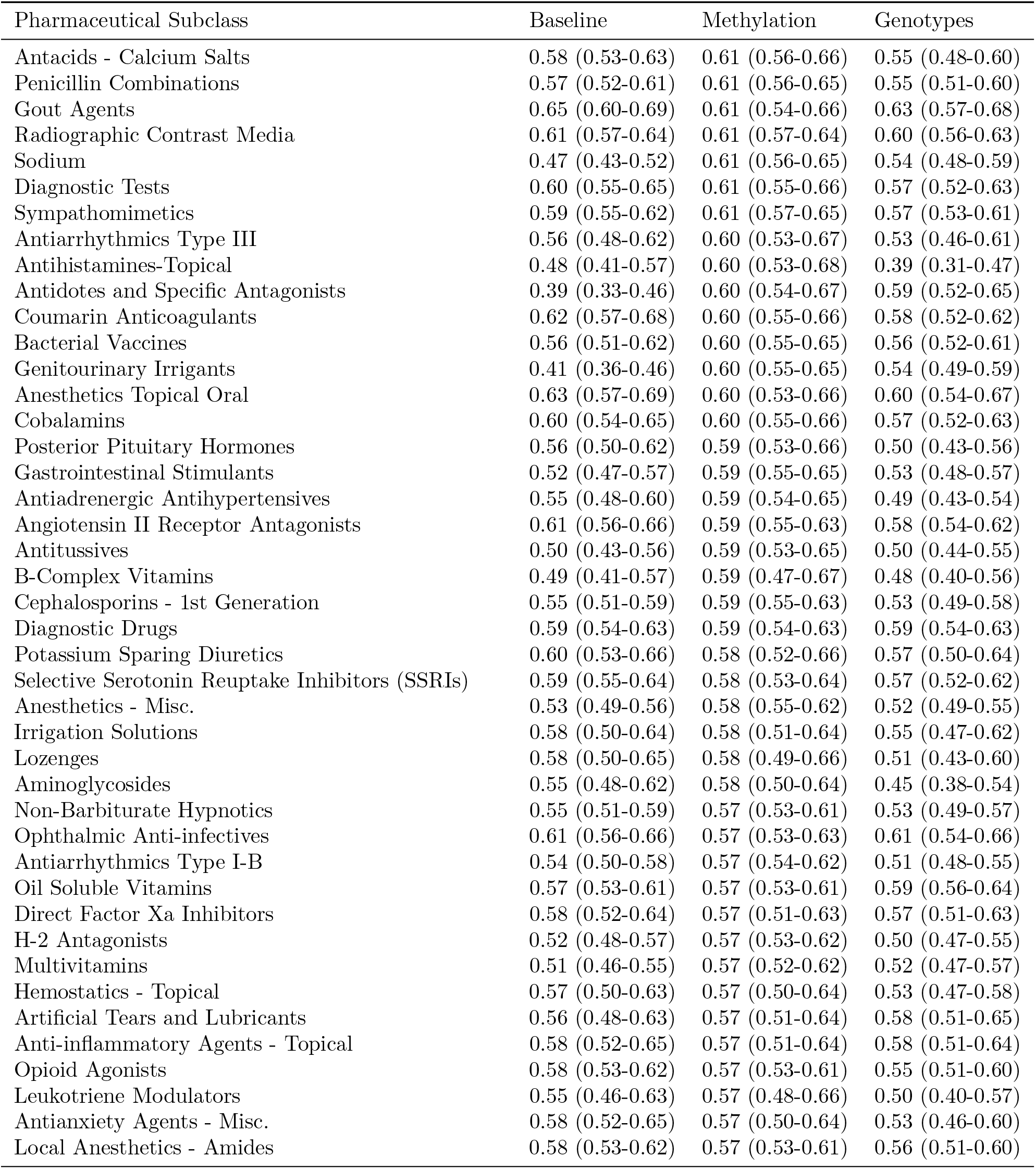
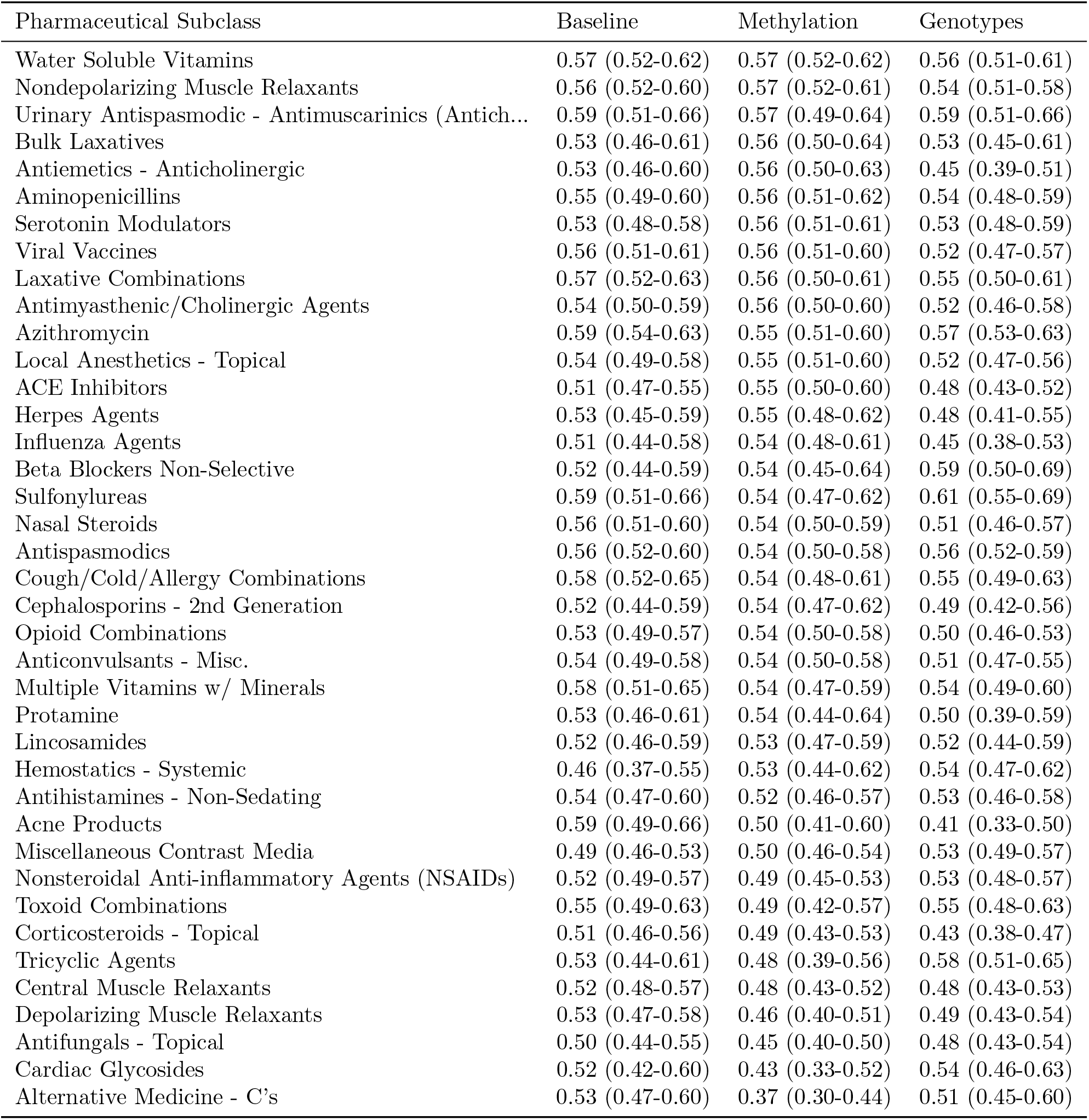
Mean (95% con_dence interval) area under the ROC curve for predicting medication usage, grouped by pharmaceutical subclass, using the baseline, methylation data, and genotype data. Con_dence intervals determined using bootstrapping.

| Pharmaceutical Subclass | Baseline | Methylation | Genotypes |
| --- | --- | --- | --- |
| CMV Agents | 0.80 (0.75-0.85) | 0.90 (0.86-0.94) | 0.79 (0.74-0.84) |
| Phosphate Binder Agents | 0.73 (0.68-0.77) | 0.88 (0.84-0.91) | 0.74 (0.70-0.78) |
| Osmotic Diuretics | 0.74 (0.69-0.78) | 0.85 (0.81-0.88) | 0.73 (0.68-0.78) |
| Hematopoietic Growth Factors | 0.70 (0.66-0.75) | 0.84 (0.81-0.87) | 0.70 (0.66-0.75) |
| B-Complex w/ Folic Acid | 0.69 (0.64-0.74) | 0.84 (0.79-0.87) | 0.67 (0.62-0.72) |
| Immunosuppressive Agents | 0.77 (0.72-0.81) | 0.83 (0.79-0.86) | 0.76 (0.70-0.80) |
| Metabolic Modifiers | 0.69 (0.63-0.74) | 0.81 (0.77-0.86) | 0.70 (0.63-0.75) |
| Prostatic Hypertrophy Agents | 0.76 (0.72-0.79) | 0.78 (0.74-0.81) | 0.76 (0.71-0.79) |
| Antacids - Bicarbonate | 0.78 (0.71-0.85) | 0.78 (0.71-0.83) | 0.75 (0.69-0.81) |
| Anti-infectives - Throat | 0.72 (0.67-0.77) | 0.77 (0.71-0.82) | 0.72 (0.66-0.78) |
| Cycloplegic Mydriatics | 0.76 (0.71-0.82) | 0.75 (0.68-0.81) | 0.76 (0.69-0.82) |
| Thrombolytic Enzymes | 0.67 (0.60-0.74) | 0.75 (0.68-0.81) | 0.63 (0.56-0.70) |
| Plasma Proteins | 0.67 (0.62-0.72) | 0.74 (0.69-0.78) | 0.66 (0.61-0.71) |
| Potassium Removing Agents | 0.64 (0.59-0.70) | 0.74 (0.69-0.79) | 0.62 (0.56-0.67) |
| Cephalosporins - 4th Generation | 0.67 (0.59-0.74) | 0.73 (0.64-0.80) | 0.65 (0.57-0.74) |
| Gallstone Solubilizing Agents | 0.59 (0.50-0.69) | 0.72 (0.64-0.79) | 0.50 (0.41-0.59) |
| Imidazole-Related Antifungals | 0.69 (0.64-0.73) | 0.72 (0.67-0.77) | 0.68 (0.63-0.73) |
| HMG CoA Reductase Inhibitors | 0.72 (0.67-0.75) | 0.72 (0.68-0.75) | 0.71 (0.68-0.74) |
| Alkalinizers | 0.70 (0.62-0.76) | 0.71 (0.63-0.78) | 0.68 (0.59-0.75) |
| Bone Density Regulators | 0.70 (0.63-0.75) | 0.71 (0.65-0.75) | 0.70 (0.64-0.76) |
| Parenteral Therapy Supplies | 0.52 (0.45-0.59) | 0.71 (0.65-0.76) | 0.60 (0.54-0.66) |
| Vasodilators | 0.54 (0.49-0.59) | 0.70 (0.66-0.75) | 0.48 (0.43-0.52) |
| Salicylates | 0.69 (0.65-0.72) | 0.70 (0.66-0.73) | 0.68 (0.64-0.72) |
| Ophthalmic Local Anesthetics | 0.69 (0.63-0.76) | 0.69 (0.63-0.76) | 0.68 (0.61-0.76) |
| Proton Pump Inhibitors | 0.62 (0.58-0.66) | 0.69 (0.66-0.73) | 0.61 (0.57-0.66) |
| Impotence Agents | 0.70 (0.64-0.75) | 0.68 (0.62-0.74) | 0.67 (0.60-0.73) |
| Ophthalmic Steroids | 0.70 (0.64-0.75) | 0.68 (0.62-0.73) | 0.68 (0.62-0.74) |
| Diabetic Supplies | 0.64 (0.58-0.69) | 0.67 (0.62-0.72) | 0.62 (0.56-0.67) |
| Phosphate | 0.65 (0.57-0.72) | 0.67 (0.60-0.74) | 0.60 (0.52-0.66) |
| Loop Diuretics | 0.63 (0.60-0.67) | 0.67 (0.63-0.71) | 0.63 (0.59-0.67) |
| Antiseptics - Mouth/Throat | 0.63 (0.57-0.68) | 0.67 (0.62-0.73) | 0.62 (0.54-0.67) |
| Anti-infective Agents - Misc. | 0.61 (0.56-0.65) | 0.67 (0.62-0.72) | 0.60 (0.55-0.65) |
| Specialty Vitamins Products | 0.60 (0.52-0.68) | 0.67 (0.58-0.75) | 0.61 (0.52-0.69) |
| Anti-infective Misc. - Combinations | 0.64 (0.59-0.68) | 0.67 (0.62-0.72) | 0.62 (0.57-0.66) |
| Iron | 0.66 (0.62-0.71) | 0.67 (0.62-0.71) | 0.66 (0.61-0.71) |
| Cephalosporins - 3rd Generation | 0.62 (0.57-0.66) | 0.66 (0.63-0.71) | 0.61 (0.57-0.65) |
| Glucocorticosteroids | 0.59 (0.55-0.63) | 0.66 (0.63-0.70) | 0.58 (0.54-0.62) |
| Antihistamines - Ethanolamines | 0.62 (0.58-0.65) | 0.66 (0.62-0.70) | 0.62 (0.58-0.66) |
| Fluoroquinolones | 0.54 (0.50-0.58) | 0.66 (0.62-0.70) | 0.52 (0.48-0.57) |
| Calcium | 0.61 (0.57-0.65) | 0.66 (0.62-0.71) | 0.60 (0.56-0.64) |
| Benzodiazepines | 0.61 (0.57-0.66) | 0.66 (0.62-0.70) | 0.61 (0.56-0.64) |
| Biguanides | 0.67 (0.62-0.71) | 0.66 (0.61-0.71) | 0.65 (0.60-0.70) |
| Local Anesthetic Combinations | 0.57 (0.53-0.62) | 0.66 (0.61-0.70) | 0.56 (0.52-0.61) |
| Ophthalmics - Misc. | 0.66 (0.60-0.72) | 0.66 (0.60-0.71) | 0.65 (0.60-0.70) |
| Antacid Combinations | 0.63 (0.58-0.68) | 0.66 (0.61-0.70) | 0.62 (0.56-0.67) |
| Dibenzapines | 0.62 (0.56-0.68) | 0.66 (0.58-0.72) | 0.58 (0.52-0.63) |
| Nitrates | 0.64 (0.61-0.68) | 0.66 (0.61-0.70) | 0.63 (0.59-0.67) |
| Insulin | 0.63 (0.58-0.67) | 0.66 (0.61-0.69) | 0.62 (0.57-0.66) |
| Liquid Vehicles | 0.59 (0.51-0.67) | 0.65 (0.58-0.73) | 0.58 (0.51-0.66) |
| Alternative Medicine - M's | 0.55 (0.49-0.61) | 0.65 (0.59-0.71) | 0.59 (0.52-0.64) |
| 5-HT3 Receptor Antagonists | 0.57 (0.53-0.60) | 0.65 (0.61-0.69) | 0.55 (0.51-0.60) |
| Antiperistaltic Agents | 0.55 (0.48-0.62) | 0.65 (0.57-0.73) | 0.51 (0.44-0.58) |
| Analgesics Other | 0.59 (0.55-0.63) | 0.65 (0.61-0.68) | 0.58 (0.54-0.62) |
| Carbohydrates | 0.61 (0.57-0.65) | 0.65 (0.61-0.69) | 0.59 (0.54-0.64) |
| Laxatives - Miscellaneous | 0.61 (0.57-0.65) | 0.65 (0.61-0.68) | 0.59 (0.55-0.64) |
| Alpha-Beta Blockers | 0.57 (0.52-0.61) | 0.65 (0.60-0.69) | 0.57 (0.53-0.61) |
| Saline Laxatives | 0.59 (0.54-0.63) | 0.64 (0.60-0.70) | 0.58 (0.54-0.62) |
| Potassium | 0.59 (0.55-0.63) | 0.64 (0.60-0.68) | 0.58 (0.54-0.62) |
| Stimulant Laxatives | 0.60 (0.56-0.64) | 0.64 (0.60-0.68) | 0.60 (0.56-0.64) |
| Urinary Anti-infectives | 0.60 (0.53-0.67) | 0.64 (0.55-0.71) | 0.51 (0.42-0.58) |
| Calcium Channel Blockers | 0.59 (0.56-0.63) | 0.64 (0.60-0.67) | 0.60 (0.56-0.63) |
| Glycopeptides | 0.60 (0.56-0.65) | 0.64 (0.59-0.68) | 0.59 (0.54-0.63) |
| Magnesium | 0.58 (0.54-0.62) | 0.64 (0.60-0.67) | 0.57 (0.53-0.61) |
| Heparins And Heparinoid-Like Agents | 0.62 (0.58-0.65) | 0.64 (0.60-0.67) | 0.61 (0.57-0.64) |
| Bicarbonates | 0.60 (0.53-0.67) | 0.63 (0.56-0.70) | 0.61 (0.53-0.68) |
| Electrolyte Mixtures | 0.58 (0.55-0.62) | 0.63 (0.60-0.67) | 0.57 (0.54-0.61) |
| Thyroid Hormones | 0.64 (0.58-0.69) | 0.63 (0.58-0.68) | 0.63 (0.58-0.68) |
| Antihypertensive Combinations | 0.64 (0.56-0.70) | 0.63 (0.57-0.71) | 0.59 (0.50-0.67) |
| Folic Acid/Folates | 0.61 (0.54-0.67) | 0.63 (0.58-0.69) | 0.60 (0.55-0.66) |
| Diagnostic Radiopharmaceuticals | 0.58 (0.54-0.62) | 0.63 (0.60-0.67) | 0.57 (0.52-0.61) |
| Surfactant Laxatives | 0.60 (0.57-0.64) | 0.63 (0.58-0.66) | 0.60 (0.56-0.64) |
| Thiazides and Thiazide-Like Diuretics | 0.65 (0.61-0.71) | 0.63 (0.58-0.68) | 0.63 (0.58-0.67) |
| Expectorants | 0.57 (0.47-0.65) | 0.63 (0.54-0.72) | 0.44 (0.36-0.52) |
| Platelet Aggregation Inhibitors | 0.64 (0.59-0.68) | 0.63 (0.58-0.67) | 0.62 (0.58-0.67) |
| Antiflatulents | 0.56 (0.50-0.61) | 0.63 (0.58-0.68) | 0.54 (0.48-0.60) |
| Vasopressors | 0.57 (0.53-0.61) | 0.62 (0.59-0.65) | 0.55 (0.52-0.60) |
| Opioid Antagonists | 0.60 (0.54-0.65) | 0.62 (0.58-0.67) | 0.60 (0.54-0.65) |
| Antibiotics - Topical | 0.55 (0.50-0.60) | 0.62 (0.58-0.67) | 0.50 (0.45-0.56) |
| Antidiarrheal/Probiotic Agents - Misc. | 0.56 (0.49-0.63) | 0.62 (0.55-0.69) | 0.51 (0.43-0.59) |
| Serotonin-Norepinephrine Reuptake Inhibitors (S... | 0.68 (0.61-0.74) | 0.62 (0.54-0.70) | 0.66 (0.58-0.73) |
| Phenothiazines | 0.52 (0.46-0.56) | 0.62 (0.56-0.66) | 0.46 (0.41-0.50) |
| Beta Blockers Cardio-Selective | 0.54 (0.51-0.58) | 0.61 (0.58-0.65) | 0.53 (0.49-0.57) |
| Misc. Nutritional Substances | 0.55 (0.49-0.59) | 0.61 (0.56-0.67) | 0.54 (0.49-0.59) |
| Alpha-2 Receptor Antagonists (Tetracyclics) | 0.66 (0.56-0.75) | 0.61 (0.51-0.70) | 0.67 (0.58-0.75) |
| Bronchodilators - Anticholinergics | 0.63 (0.58-0.67) | 0.61 (0.56-0.67) | 0.61 (0.56-0.66) |
| Tetracyclines | 0.51 (0.45-0.59) | 0.61 (0.56-0.67) | 0.40 (0.35-0.46) |
| Antacids - Calcium Salts | 0.58 (0.53-0.63) | 0.61 (0.56-0.66) | 0.55 (0.48-0.60) |
| Penicillin Combinations | 0.57 (0.52-0.61) | 0.61 (0.56-0.65) | 0.55 (0.51-0.60) |
| Gout Agents | 0.65 (0.60-0.69) | 0.61 (0.54-0.66) | 0.63 (0.57-0.68) |
| Radiographic Contrast Media | 0.61 (0.57-0.64) | 0.61 (0.57-0.64) | 0.60 (0.56-0.63) |
| Sodium | 0.47 (0.43-0.52) | 0.61 (0.56-0.65) | 0.54 (0.48-0.59) |
| Diagnostic Tests | 0.60 (0.55-0.65) | 0.61 (0.55-0.66) | 0.57 (0.52-0.63) |
| Sympathomimetics | 0.59 (0.55-0.62) | 0.61 (0.57-0.65) | 0.57 (0.53-0.61) |
| Antiarrhythmics Type III | 0.56 (0.48-0.62) | 0.60 (0.53-0.67) | 0.53 (0.46-0.61) |
| Antihistamines-Topical | 0.48 (0.41-0.57) | 0.60 (0.53-0.68) | 0.39 (0.31-0.47) |
| Antidotes and Specific Antagonists | 0.39 (0.33-0.46) | 0.60 (0.54-0.67) | 0.59 (0.52-0.65) |
| Coumarin Anticoagulants | 0.62 (0.57-0.68) | 0.60 (0.55-0.66) | 0.58 (0.52-0.62) |
| Bacterial Vaccines | 0.56 (0.51-0.62) | 0.60 (0.55-0.65) | 0.56 (0.52-0.61) |
| Genitourinary Irrigants | 0.41 (0.36-0.46) | 0.60 (0.55-0.65) | 0.54 (0.49-0.59) |
| Anesthetics Topical Oral | 0.63 (0.57-0.69) | 0.60 (0.53-0.66) | 0.60 (0.54-0.67) |
| Cobalamins | 0.60 (0.54-0.65) | 0.60 (0.55-0.66) | 0.57 (0.52-0.63) |
| Posterior Pituitary Hormones | 0.56 (0.50-0.62) | 0.59 (0.53-0.66) | 0.50 (0.43-0.56) |
| Gastrointestinal Stimulants | 0.52 (0.47-0.57) | 0.59 (0.55-0.65) | 0.53 (0.48-0.57) |
| Antiadrenergic Antihypertensives | 0.55 (0.48-0.60) | 0.59 (0.54-0.65) | 0.49 (0.43-0.54) |
| Angiotensin II Receptor Antagonists | 0.61 (0.56-0.66) | 0.59 (0.55-0.63) | 0.58 (0.54-0.62) |
| Antitussives | 0.50 (0.43-0.56) | 0.59 (0.53-0.65) | 0.50 (0.44-0.55) |
| B-Complex Vitamins | 0.49 (0.41-0.57) | 0.59 (0.47-0.67) | 0.48 (0.40-0.56) |
| Cephalosporins - 1st Generation | 0.55 (0.51-0.59) | 0.59 (0.55-0.63) | 0.53 (0.49-0.58) |
| Diagnostic Drugs | 0.59 (0.54-0.63) | 0.59 (0.54-0.63) | 0.59 (0.54-0.63) |
| Potassium Sparing Diuretics | 0.60 (0.53-0.66) | 0.58 (0.52-0.66) | 0.57 (0.50-0.64) |
| Selective Serotonin Reuptake Inhibitors (SSRIs) | 0.59 (0.55-0.64) | 0.58 (0.53-0.64) | 0.57 (0.52-0.62) |
| Anesthetics - Misc. | 0.53 (0.49-0.56) | 0.58 (0.55-0.62) | 0.52 (0.49-0.55) |
| Irrigation Solutions | 0.58 (0.50-0.64) | 0.58 (0.51-0.64) | 0.55 (0.47-0.62) |
| Lozenges | 0.58 (0.50-0.65) | 0.58 (0.49-0.66) | 0.51 (0.43-0.60) |
| Aminoglycosides | 0.55 (0.48-0.62) | 0.58 (0.50-0.64) | 0.45 (0.38-0.54) |
| Non-Barbiturate Hypnotics | 0.55 (0.51-0.59) | 0.57 (0.53-0.61) | 0.53 (0.49-0.57) |
| Ophthalmic Anti-infectives | 0.61 (0.56-0.66) | 0.57 (0.53-0.63) | 0.61 (0.54-0.66) |
| Antiarrhythmics Type I-B | 0.54 (0.50-0.58) | 0.57 (0.54-0.62) | 0.51 (0.48-0.55) |
| Oil Soluble Vitamins | 0.57 (0.53-0.61) | 0.57 (0.53-0.61) | 0.59 (0.56-0.64) |
| Direct Factor Xa Inhibitors | 0.58 (0.52-0.64) | 0.57 (0.51-0.63) | 0.57 (0.51-0.63) |
| H-2 Antagonists | 0.52 (0.48-0.57) | 0.57 (0.53-0.62) | 0.50 (0.47-0.55) |
| Multivitamins | 0.51 (0.46-0.55) | 0.57 (0.52-0.62) | 0.52 (0.47-0.57) |
| Hemostatics - Topical | 0.57 (0.50-0.63) | 0.57 (0.50-0.64) | 0.53 (0.47-0.58) |
| Artificial Tears and Lubricants | 0.56 (0.48-0.63) | 0.57 (0.51-0.64) | 0.58 (0.51-0.65) |
| Anti-inflammatory Agents - Topical | 0.58 (0.52-0.65) | 0.57 (0.51-0.64) | 0.58 (0.51-0.64) |
| Opioid Agonists | 0.58 (0.53-0.62) | 0.57 (0.53-0.61) | 0.55 (0.51-0.60) |
| Leukotriene Modulators | 0.55 (0.46-0.63) | 0.57 (0.48-0.66) | 0.50 (0.40-0.57) |
| Antianxiety Agents - Misc. | 0.58 (0.52-0.65) | 0.57 (0.50-0.64) | 0.53 (0.46-0.60) |
| Local Anesthetics - Amides | 0.58 (0.53-0.62) | 0.57 (0.53-0.61) | 0.56 (0.51-0.60) |
| Water Soluble Vitamins | 0.57 (0.52-0.62) | 0.57 (0.52-0.62) | 0.56 (0.51-0.61) |
| Nondepolarizing Muscle Relaxants | 0.56 (0.52-0.60) | 0.57 (0.52-0.61) | 0.54 (0.51-0.58) |
| Urinary Antispasmodic - Antimuscarinics (Antich... | 0.59 (0.51-0.66) | 0.57 (0.49-0.64) | 0.59 (0.51-0.66) |
| Bulk Laxatives | 0.53 (0.46-0.61) | 0.56 (0.50-0.64) | 0.53 (0.45-0.61) |
| Antiemetics - Anticholinergic | 0.53 (0.46-0.60) | 0.56 (0.50-0.63) | 0.45 (0.39-0.51) |
| Aminopenicillins | 0.55 (0.49-0.60) | 0.56 (0.51-0.62) | 0.54 (0.48-0.59) |
| Serotonin Modulators | 0.53 (0.48-0.58) | 0.56 (0.51-0.61) | 0.53 (0.48-0.59) |
| Viral Vaccines | 0.56 (0.51-0.61) | 0.56 (0.51-0.60) | 0.52 (0.47-0.57) |
| Laxative Combinations | 0.57 (0.52-0.63) | 0.56 (0.50-0.61) | 0.55 (0.50-0.61) |
| Antimyasthenic/Cholinergic Agents | 0.54 (0.50-0.59) | 0.56 (0.50-0.60) | 0.52 (0.46-0.58) |
| Azithromycin | 0.59 (0.54-0.63) | 0.55 (0.51-0.60) | 0.57 (0.53-0.63) |
| Local Anesthetics - Topical | 0.54 (0.49-0.58) | 0.55 (0.51-0.60) | 0.52 (0.47-0.56) |
| ACE Inhibitors | 0.51 (0.47-0.55) | 0.55 (0.50-0.60) | 0.48 (0.43-0.52) |
| Herpes Agents | 0.53 (0.45-0.59) | 0.55 (0.48-0.62) | 0.48 (0.41-0.55) |
| Influenza Agents | 0.51 (0.44-0.58) | 0.54 (0.48-0.61) | 0.45 (0.38-0.53) |
| Beta Blockers Non-Selective | 0.52 (0.44-0.59) | 0.54 (0.45-0.64) | 0.59 (0.50-0.69) |
| Sulfonylureas | 0.59 (0.51-0.66) | 0.54 (0.47-0.62) | 0.61 (0.55-0.69) |
| Nasal Steroids | 0.56 (0.51-0.60) | 0.54 (0.50-0.59) | 0.51 (0.46-0.57) |
| Antispasmodics | 0.56 (0.52-0.60) | 0.54 (0.50-0.58) | 0.56 (0.52-0.59) |
| Cough/Cold/Allergy Combinations | 0.58 (0.52-0.65) | 0.54 (0.48-0.61) | 0.55 (0.49-0.63) |
| Cephalosporins - 2nd Generation | 0.52 (0.44-0.59) | 0.54 (0.47-0.62) | 0.49 (0.42-0.56) |
| Opioid Combinations | 0.53 (0.49-0.57) | 0.54 (0.50-0.58) | 0.50 (0.46-0.53) |
| Anticonvulsants - Misc. | 0.54 (0.49-0.58) | 0.54 (0.50-0.58) | 0.51 (0.47-0.55) |
| Multiple Vitamins w/ Minerals | 0.58 (0.51-0.65) | 0.54 (0.47-0.59) | 0.54 (0.49-0.60) |
| Protamine | 0.53 (0.46-0.61) | 0.54 (0.44-0.64) | 0.50 (0.39-0.59) |
| Lincosamides | 0.52 (0.46-0.59) | 0.53 (0.47-0.59) | 0.52 (0.44-0.59) |
| Hemostatics - Systemic | 0.46 (0.37-0.55) | 0.53 (0.44-0.62) | 0.54 (0.47-0.62) |
| Antihistamines - Non-Sedating | 0.54 (0.47-0.60) | 0.52 (0.46-0.57) | 0.53 (0.46-0.58) |
| Acne Products | 0.59 (0.49-0.66) | 0.50 (0.41-0.60) | 0.41 (0.33-0.50) |
| Miscellaneous Contrast Media | 0.49 (0.46-0.53) | 0.50 (0.46-0.54) | 0.53 (0.49-0.57) |
| Nonsteroidal Anti-inflammatory Agents (NSAIDs) | 0.52 (0.49-0.57) | 0.49 (0.45-0.53) | 0.53 (0.48-0.57) |
| Toxoid Combinations | 0.55 (0.49-0.63) | 0.49 (0.42-0.57) | 0.55 (0.48-0.63) |
| Corticosteroids - Topical | 0.51 (0.46-0.56) | 0.49 (0.43-0.53) | 0.43 (0.38-0.47) |
| Tricyclic Agents | 0.53 (0.44-0.61) | 0.48 (0.39-0.56) | 0.58 (0.51-0.65) |
| Central Muscle Relaxants | 0.52 (0.48-0.57) | 0.48 (0.43-0.52) | 0.48 (0.43-0.53) |
| Depolarizing Muscle Relaxants | 0.53 (0.47-0.58) | 0.46 (0.40-0.51) | 0.49 (0.43-0.54) |
| Antifungals - Topical | 0.50 (0.44-0.55) | 0.45 (0.40-0.50) | 0.48 (0.43-0.54) |
| Cardiac Glycosides | 0.52 (0.42-0.60) | 0.43 (0.33-0.52) | 0.54 (0.46-0.63) |
| Alternative Medicine - C's | 0.53 (0.47-0.60) | 0.37 (0.30-0.44) | 0.51 (0.45-0.60) |

**Table S3.**
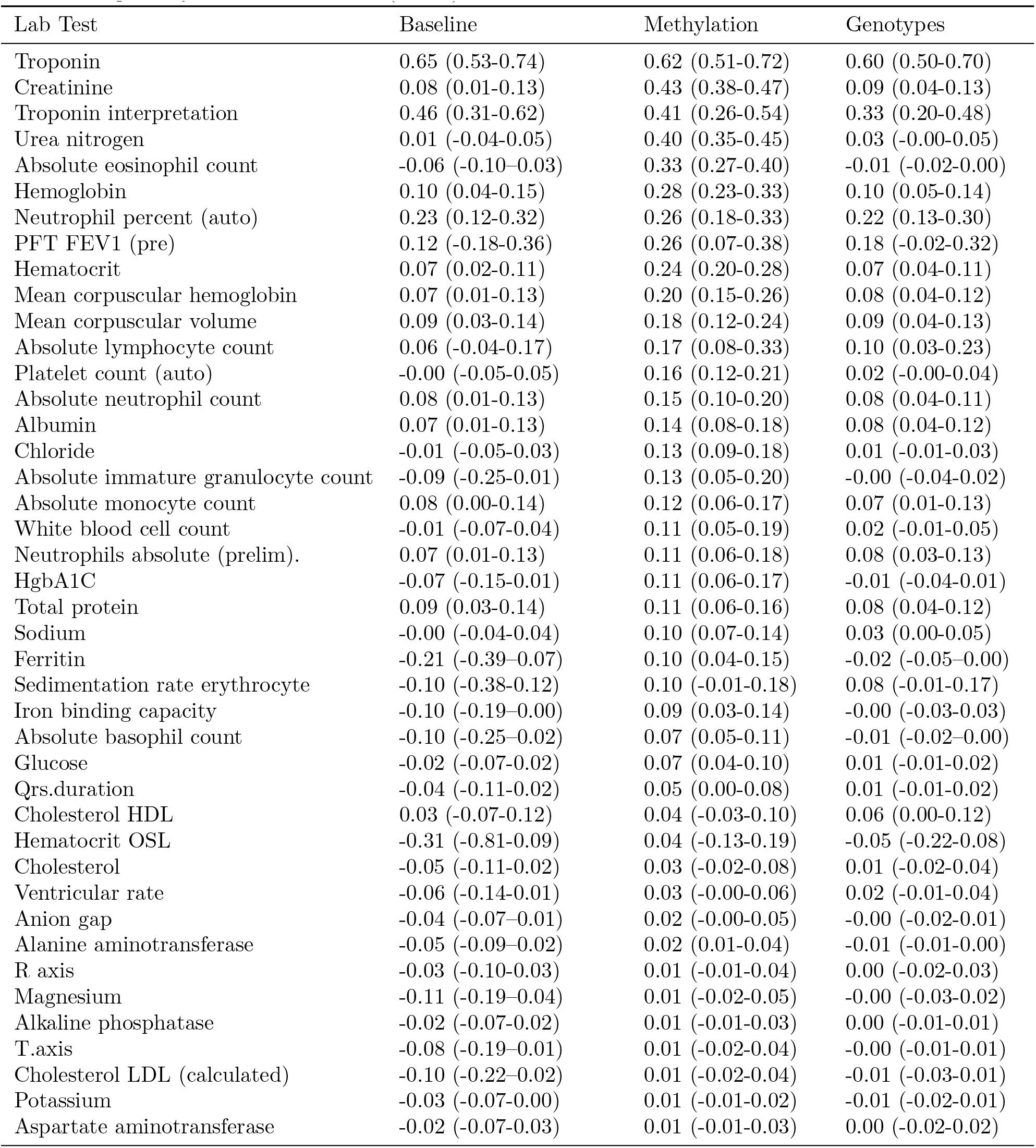
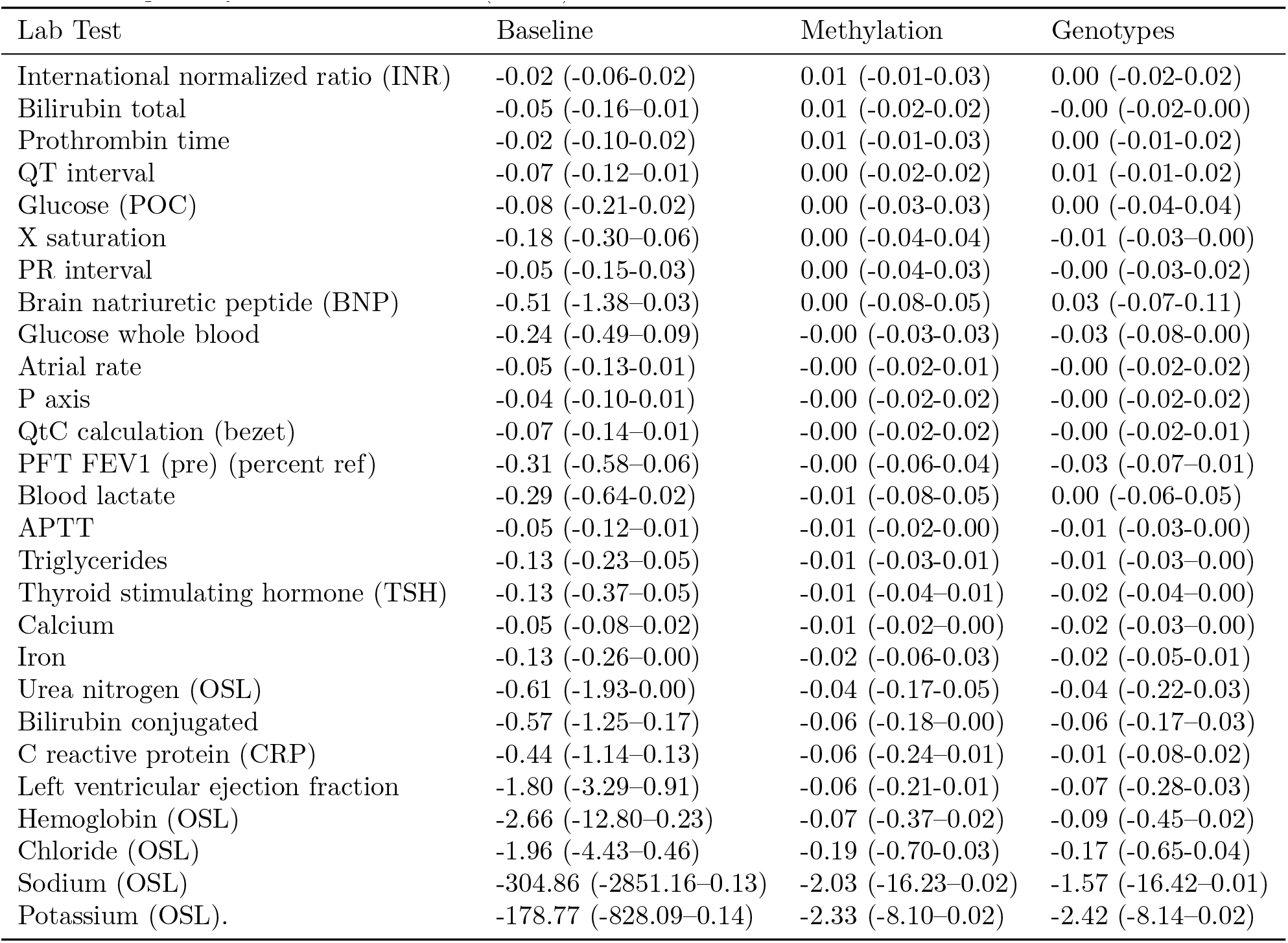
Mean (95% con_dence interval) R2 for predicting the most recent lab result using the baseline, methylation data, and genotype data. Con_dence intervals determined using bootstrapping. Activated Partial Thromboplastin Time (APTT); Point of care (POC); Pulmonary function test (PFT); Forced expiratory volume in 1 second (FEV1)

| Lab Test | Baseline | Methylation | Genotypes |
| --- | --- | --- | --- |
| Troponin | 0.65 (0.53-0.74) | 0.62 (0.51-0.72) | 0.60 (0.50-0.70) |
| Creatinine | 0.08 (0.01-0.13) | 0.43 (0.38-0.47) | 0.09 (0.04-0.13) |
| Troponin interpretation | 0.46 (0.31-0.62) | 0.41 (0.26-0.54) | 0.33 (0.20-0.48) |
| Urea nitrogen | 0.01 (-0.04-0.05) | 0.40 (0.35-0.45) | 0.03 (-0.00-0.05) |
| Absolute eosinophil count | -0.06 (-0.10-0.03) | 0.33 (0.27-0.40) | -0.01 (-0.02-0.00) |
| Hemoglobin | 0.10 (0.04-0.15) | 0.28 (0.23-0.33) | 0.10 (0.05-0.14) |
| Neutrophil percent (auto) | 0.23 (0.12-0.32) | 0.26 (0.18-0.33) | 0.22 (0.13-0.30) |
| PFT FEV1 (pre) | 0.12 (-0.18-0.36) | 0.26 (0.07-0.38) | 0.18 (-0.02-0.32) |
| Hematocrit | 0.07 (0.02-0.11) | 0.24 (0.20-0.28) | 0.07 (0.04-0.11) |
| Mean corpuscular hemoglobin | 0.07 (0.01-0.13) | 0.20 (0.15-0.26) | 0.08 (0.04-0.12) |
| Mean corpuscular volume | 0.09 (0.03-0.14) | 0.18 (0.12-0.24) | 0.09 (0.04-0.13) |
| Absolute lymphocyte count | 0.06 (-0.04-0.17) | 0.17 (0.08-0.33) | 0.10 (0.03-0.23) |
| Platelet count (auto) | -0.00 (-0.05-0.05) | 0.16 (0.12-0.21) | 0.02 (-0.00-0.04) |
| Absolute neutrophil count | 0.08 (0.01-0.13) | 0.15 (0.10-0.20) | 0.08 (0.04-0.11) |
| Albumin | 0.07 (0.01-0.13) | 0.14 (0.08-0.18) | 0.08 (0.04-0.12) |
| Chloride | -0.01 (-0.05-0.03) | 0.13 (0.09-0.18) | 0.01 (-0.01-0.03) |
| Absolute immature granulocyte count | -0.09 (-0.25-0.01) | 0.13 (0.05-0.20) | -0.00 (-0.04-0.02) |
| Absolute monocyte count | 0.08 (0.00-0.14) | 0.12 (0.06-0.17) | 0.07 (0.01-0.13) |
| White blood cell count | -0.01 (-0.07-0.04) | 0.11 (0.05-0.19) | 0.02 (-0.01-0.05) |
| Neutrophils absolute (prelim). | 0.07 (0.01-0.13) | 0.11 (0.06-0.18) | 0.08 (0.03-0.13) |
| HgbA1C | -0.07 (-0.15-0.01) | 0.11 (0.06-0.17) | -0.01 (-0.04-0.01) |
| Total protein | 0.09 (0.03-0.14) | 0.11 (0.06-0.16) | 0.08 (0.04-0.12) |
| Sodium | -0.00 (-0.04-0.04) | 0.10 (0.07-0.14) | 0.03 (0.00-0.05) |
| Ferritin | -0.21 (-0.39-0.07) | 0.10 (0.04-0.15) | -0.02 (-0.05-0.00) |
| Sedimentation rate erythrocyte | -0.10 (-0.38-0.12) | 0.10 (-0.01-0.18) | 0.08 (-0.01-0.17) |
| Iron binding capacity | -0.10 (-0.19-0.00) | 0.09 (0.03-0.14) | -0.00 (-0.03-0.03) |
| Absolute basophil count | -0.10 (-0.25-0.02) | 0.07 (0.05-0.11) | -0.01 (-0.02-0.00) |
| Glucose | -0.02 (-0.07-0.02) | 0.07 (0.04-0.10) | 0.01 (-0.01-0.02) |
| Qrs.duration | -0.04 (-0.11-0.02) | 0.05 (0.00-0.08) | 0.01 (-0.01-0.02) |
| Cholesterol HDL | 0.03 (-0.07-0.12) | 0.04 (-0.03-0.10) | 0.06 (0.00-0.12) |
| Hematocrit OSL | -0.31 (-0.81-0.09) | 0.04 (-0.13-0.19) | -0.05 (-0.22-0.08) |
| Cholesterol | -0.05 (-0.11-0.02) | 0.03 (-0.02-0.08) | 0.01 (-0.02-0.04) |
| Ventricular rate | -0.06 (-0.14-0.01) | 0.03 (-0.00-0.06) | 0.02 (-0.01-0.04) |
| Anion gap | -0.04 (-0.07-0.01) | 0.02 (-0.00-0.05) | -0.00 (-0.02-0.01) |
| Alanine aminotransferase | -0.05 (-0.09-0.02) | 0.02 (0.01-0.04) | -0.01 (-0.01-0.00) |
| R axis | -0.03 (-0.10-0.03) | 0.01 (-0.01-0.04) | 0.00 (-0.02-0.03) |
| Magnesium | -0.11 (-0.19-0.04) | 0.01 (-0.02-0.05) | -0.00 (-0.03-0.02) |
| Alkaline phosphatase | -0.02 (-0.07-0.02) | 0.01 (-0.01-0.03) | 0.00 (-0.01-0.01) |
| T.axis | -0.08 (-0.19-0.01) | 0.01 (-0.02-0.04) | -0.00 (-0.01-0.01) |
| Cholesterol LDL (calculated) | -0.10 (-0.22-0.02) | 0.01 (-0.02-0.04) | -0.01 (-0.03-0.01) |
| Potassium | -0.03 (-0.07-0.00) | 0.01 (-0.01-0.02) | -0.01 (-0.02-0.01) |
| Aspartate aminotransferase | -0.02 (-0.07-0.03) | 0.01 (-0.01-0.03) | 0.00 (-0.02-0.02) |
| International normalized ratio (INR) | -0.02 (-0.06-0.02) | 0.01 (-0.01-0.03) | 0.00 (-0.02-0.02) |
| Bilirubin total | -0.05 (-0.16-0.01) | 0.01 (-0.02-0.02) | -0.00 (-0.02-0.00) |
| Prothrombin time | -0.02 (-0.10-0.02) | 0.01 (-0.01-0.03) | 0.00 (-0.01-0.02) |
| QT interval | -0.07 (-0.12-0.01) | 0.00 (-0.02-0.02) | 0.01 (-0.01-0.02) |
| Glucose (POC) | -0.08 (-0.21-0.02) | 0.00 (-0.03-0.03) | 0.00 (-0.04-0.04) |
| X saturation | -0.18 (-0.30-0.06) | 0.00 (-0.04-0.04) | -0.01 (-0.03-0.00) |
| PR interval | -0.05 (-0.15-0.03) | 0.00 (-0.04-0.03) | -0.00 (-0.03-0.02) |
| Brain natriuretic peptide (BNP) | -0.51 (-1.38-0.03) | 0.00 (-0.08-0.05) | 0.03 (-0.07-0.11) |
| Glucose whole blood | -0.24 (-0.49-0.09) | -0.00 (-0.03-0.03) | -0.03 (-0.08-0.00) |
| Atrial rate | -0.05 (-0.13-0.01) | -0.00 (-0.02-0.01) | -0.00 (-0.02-0.02) |
| P axis | -0.04 (-0.10-0.01) | -0.00 (-0.02-0.02) | -0.00 (-0.02-0.02) |
| QtC calculation (bezet) | -0.07 (-0.14-0.01) | -0.00 (-0.02-0.02) | -0.00 (-0.02-0.01) |
| PFT FEV1 (pre) (percent ref) | -0.31 (-0.58-0.06) | -0.00 (-0.06-0.04) | -0.03 (-0.07-0.01) |
| Blood lactate | -0.29 (-0.64-0.02) | -0.01 (-0.08-0.05) | 0.00 (-0.06-0.05) |
| APTT | -0.05 (-0.12-0.01) | -0.01 (-0.02-0.00) | -0.01 (-0.03-0.00) |
| Triglycerides | -0.13 (-0.23-0.05) | -0.01 (-0.03-0.01) | -0.01 (-0.03-0.00) |
| Thyroid stimulating hormone (TSH) | -0.13 (-0.37-0.05) | -0.01 (-0.04-0.01) | -0.02 (-0.04-0.00) |
| Calcium | -0.05 (-0.08-0.02) | -0.01 (-0.02-0.00) | -0.02 (-0.03-0.00) |
| Iron | -0.13 (-0.26-0.00) | -0.02 (-0.06-0.03) | -0.02 (-0.05-0.01) |
| Urea nitrogen (OSL) | -0.61 (-1.93-0.00) | -0.04 (-0.17-0.05) | -0.04 (-0.22-0.03) |
| Bilirubin conjugated | -0.57 (-1.25-0.17) | -0.06 (-0.18-0.00) | -0.06 (-0.17-0.03) |
| C reactive protein (CRP) | -0.44 (-1.14-0.13) | -0.06 (-0.24-0.01) | -0.01 (-0.08-0.02) |
| Left ventricular ejection fraction | -1.80 (-3.29-0.91) | -0.06 (-0.21-0.01) | -0.07 (-0.28-0.03) |
| Hemoglobin (OSL) | -2.66 (-12.80-0.23) | -0.07 (-0.37-0.02) | -0.09 (-0.45-0.02) |
| Chloride (OSL) | -1.96 (-4.43-0.46) | -0.19 (-0.70-0.03) | -0.17 (-0.65-0.04) |
| Sodium (OSL) | -304.86 (-2851.16-0.13) | -2.03 (-16.23-0.02) | -1.57 (-16.42-0.01) |
| Potassium (OSL). | -178.77 (-828.09-0.14) | -2.33 (-8.10-0.02) | -2.42 (-8.14-0.02) |

**Table S4.**
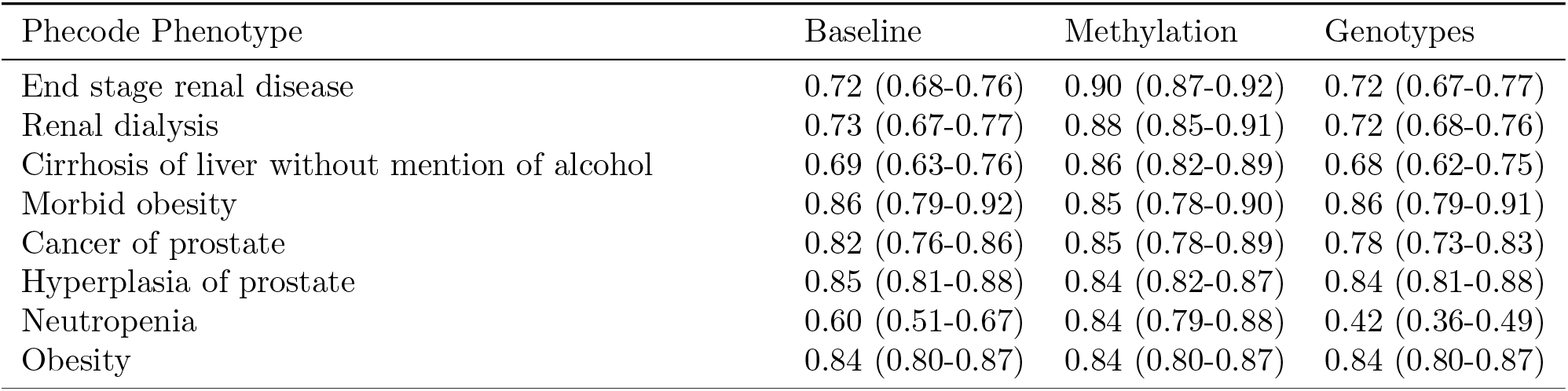
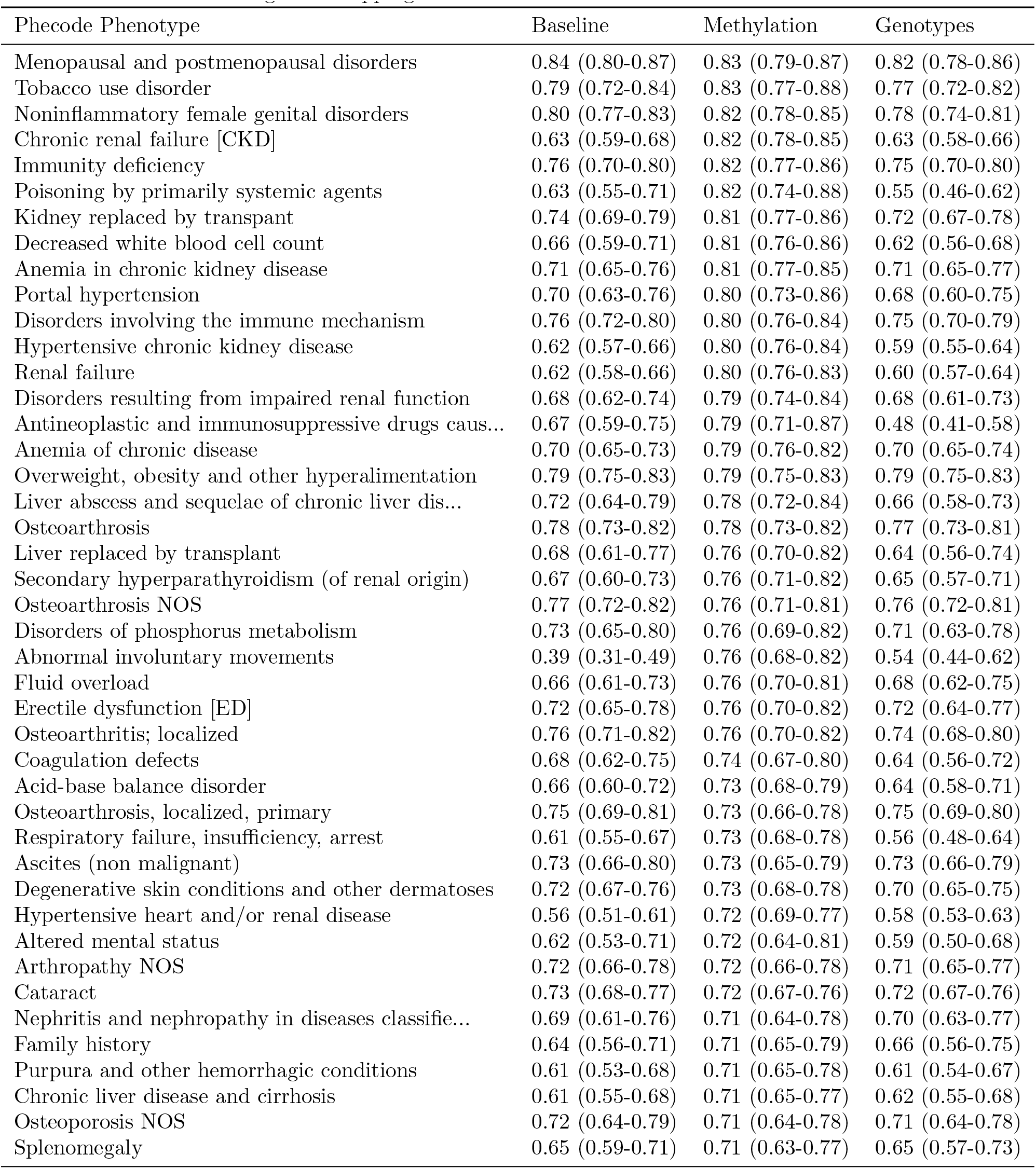
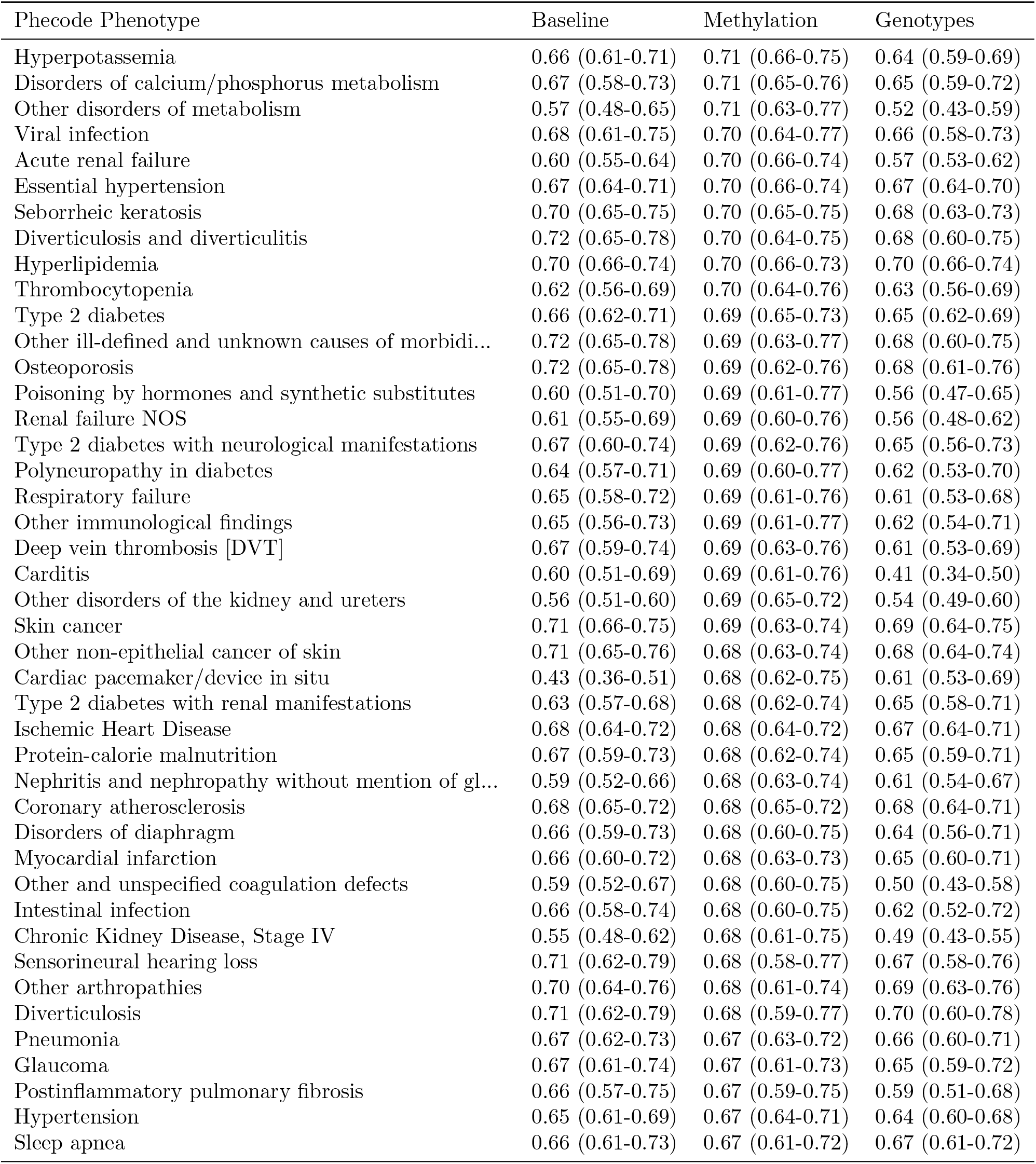
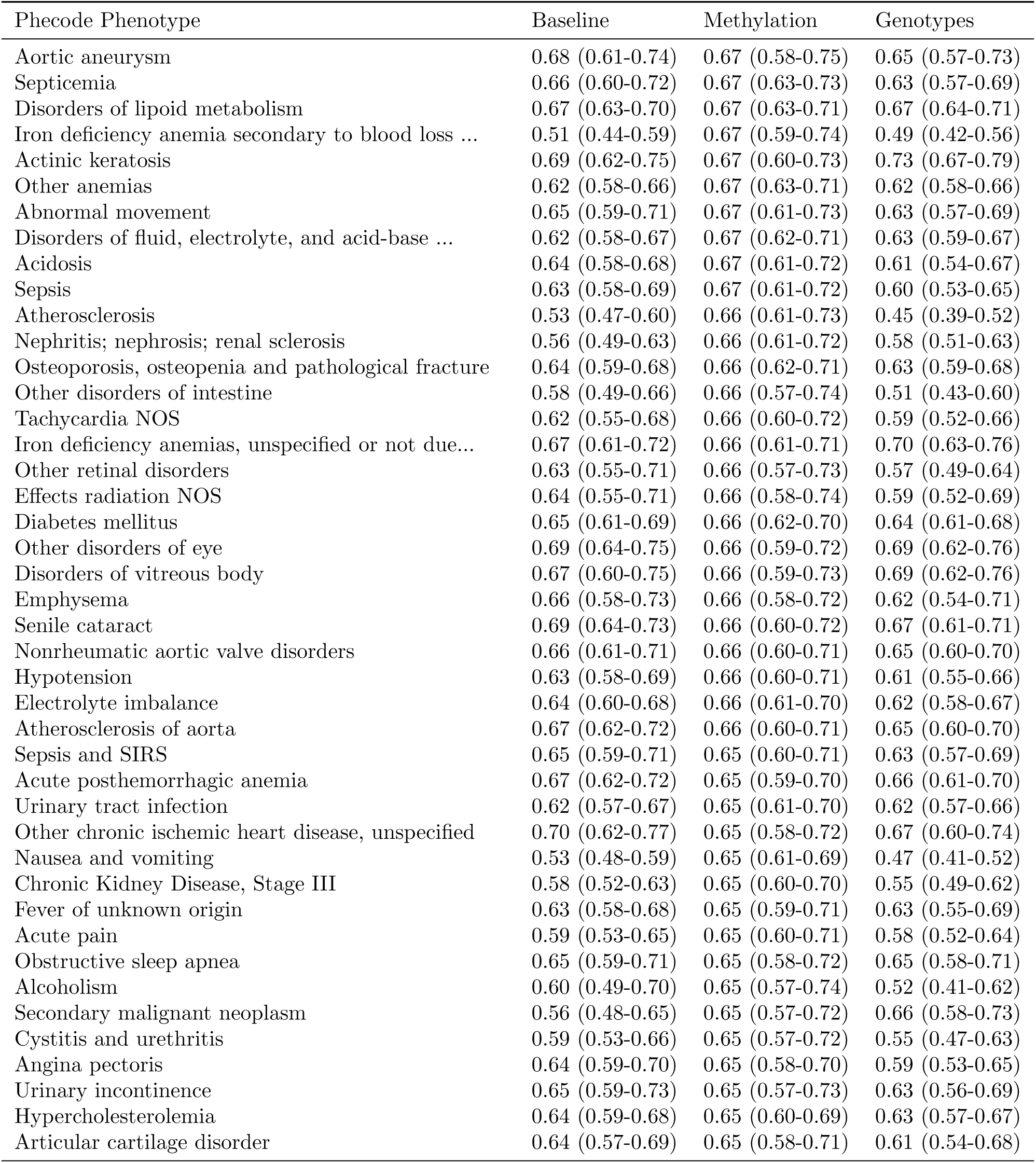
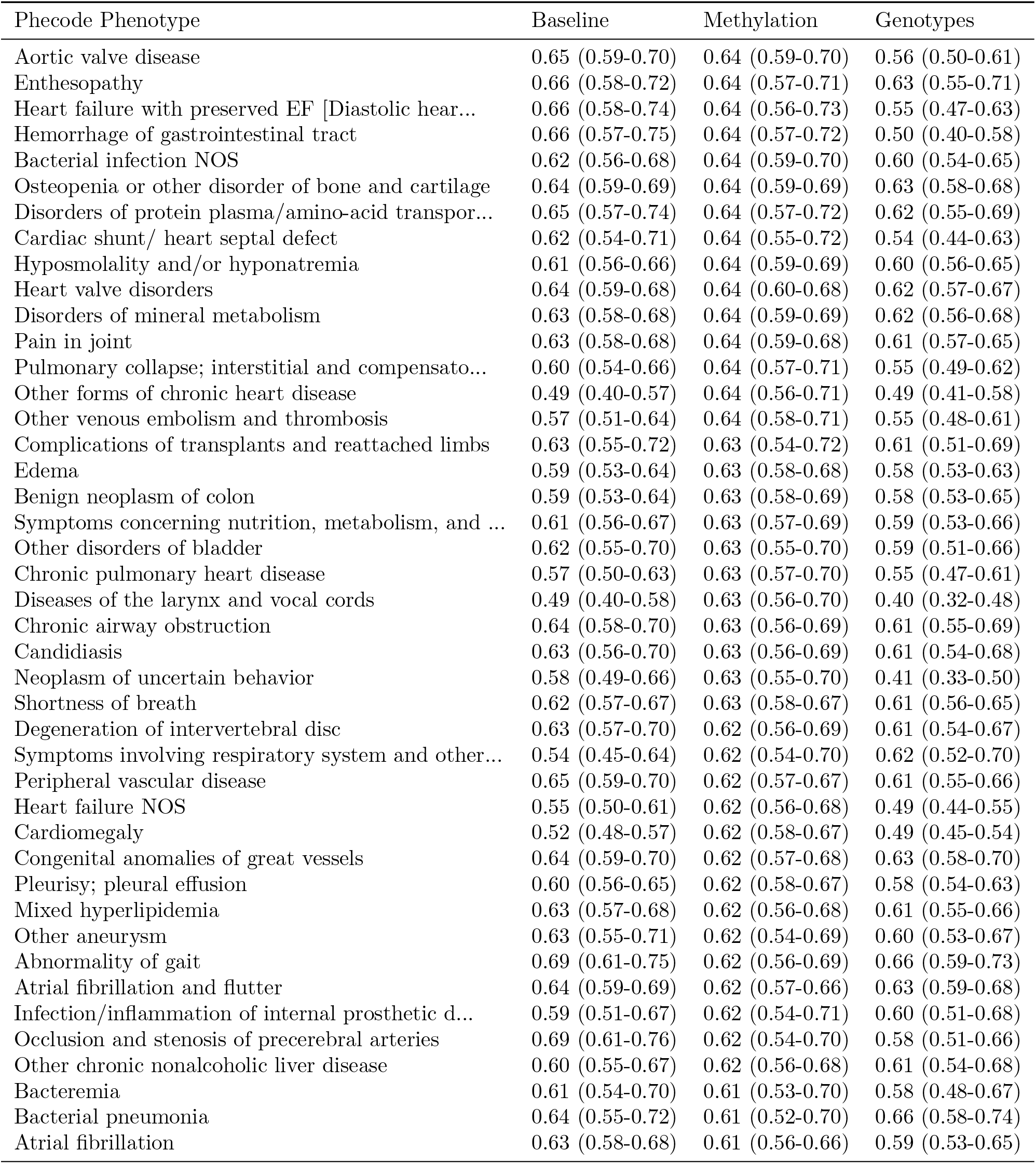
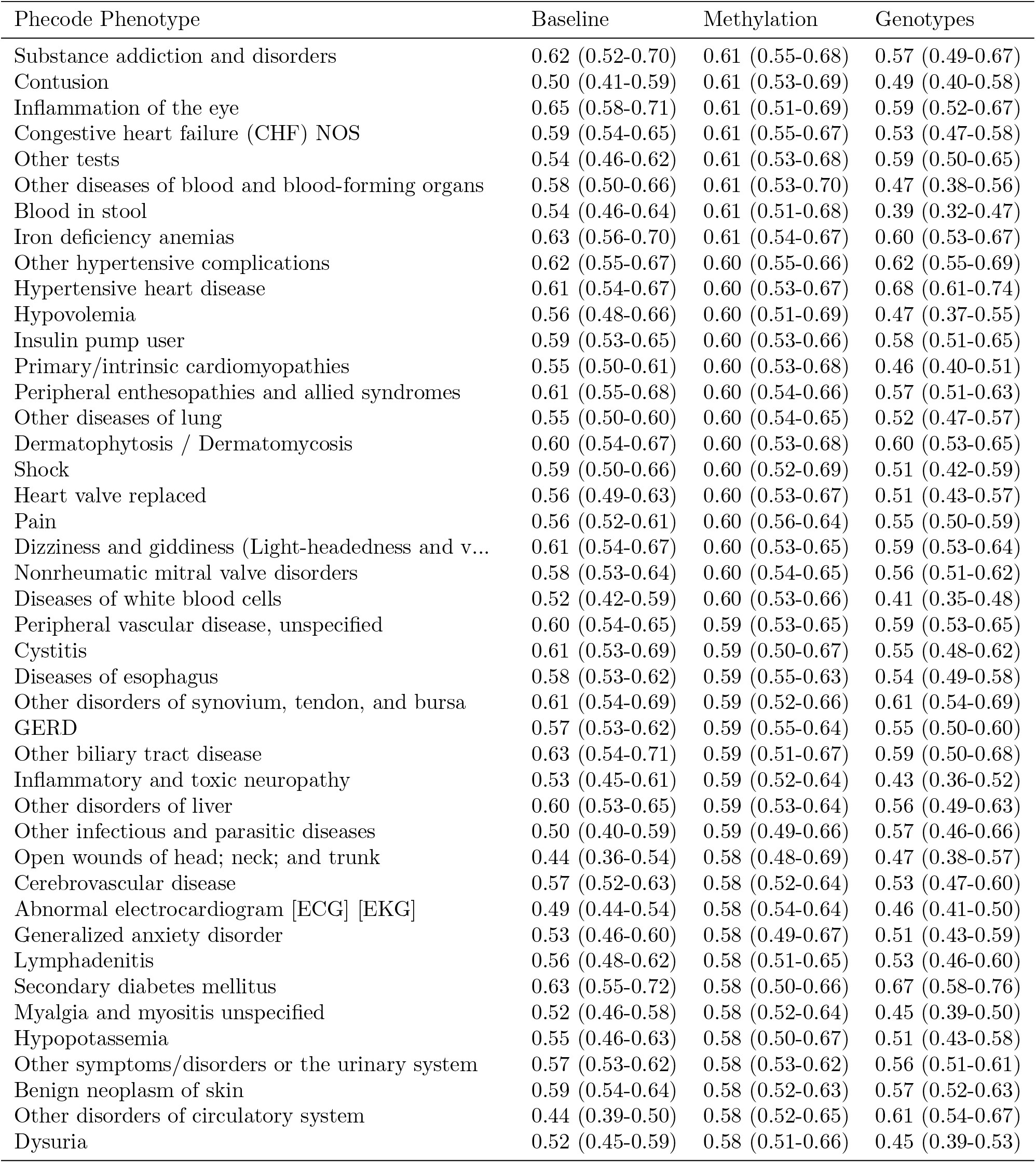
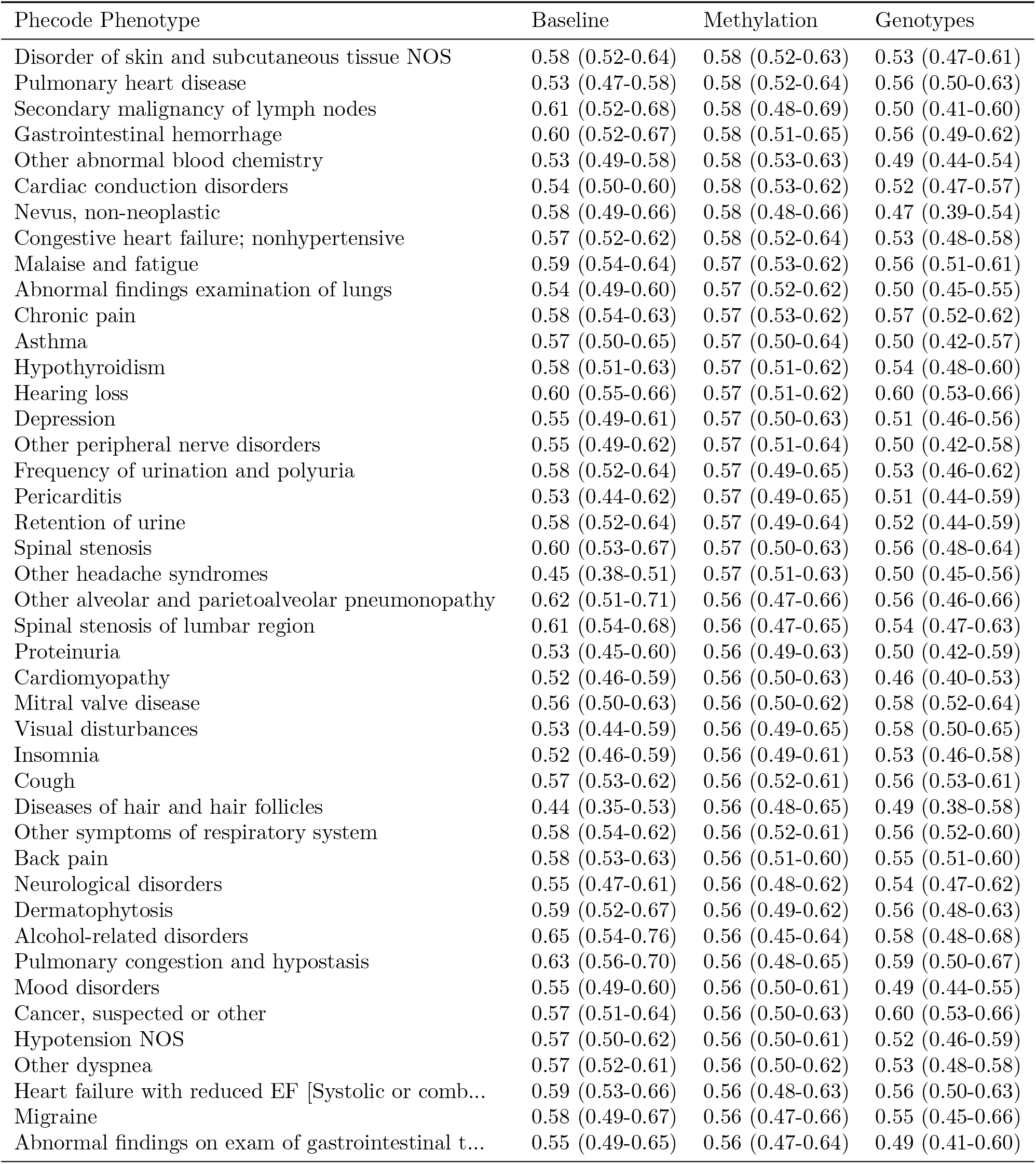
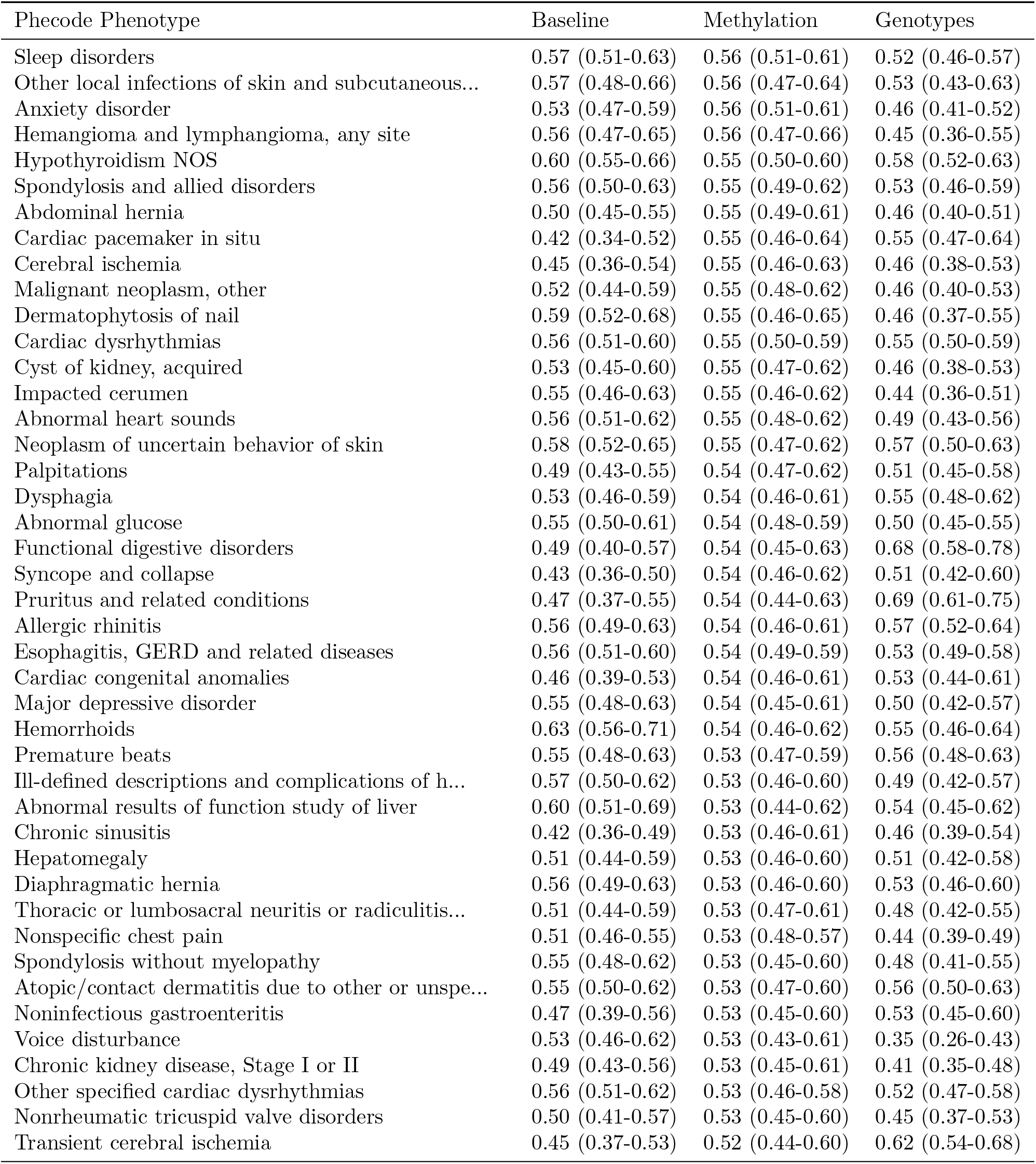
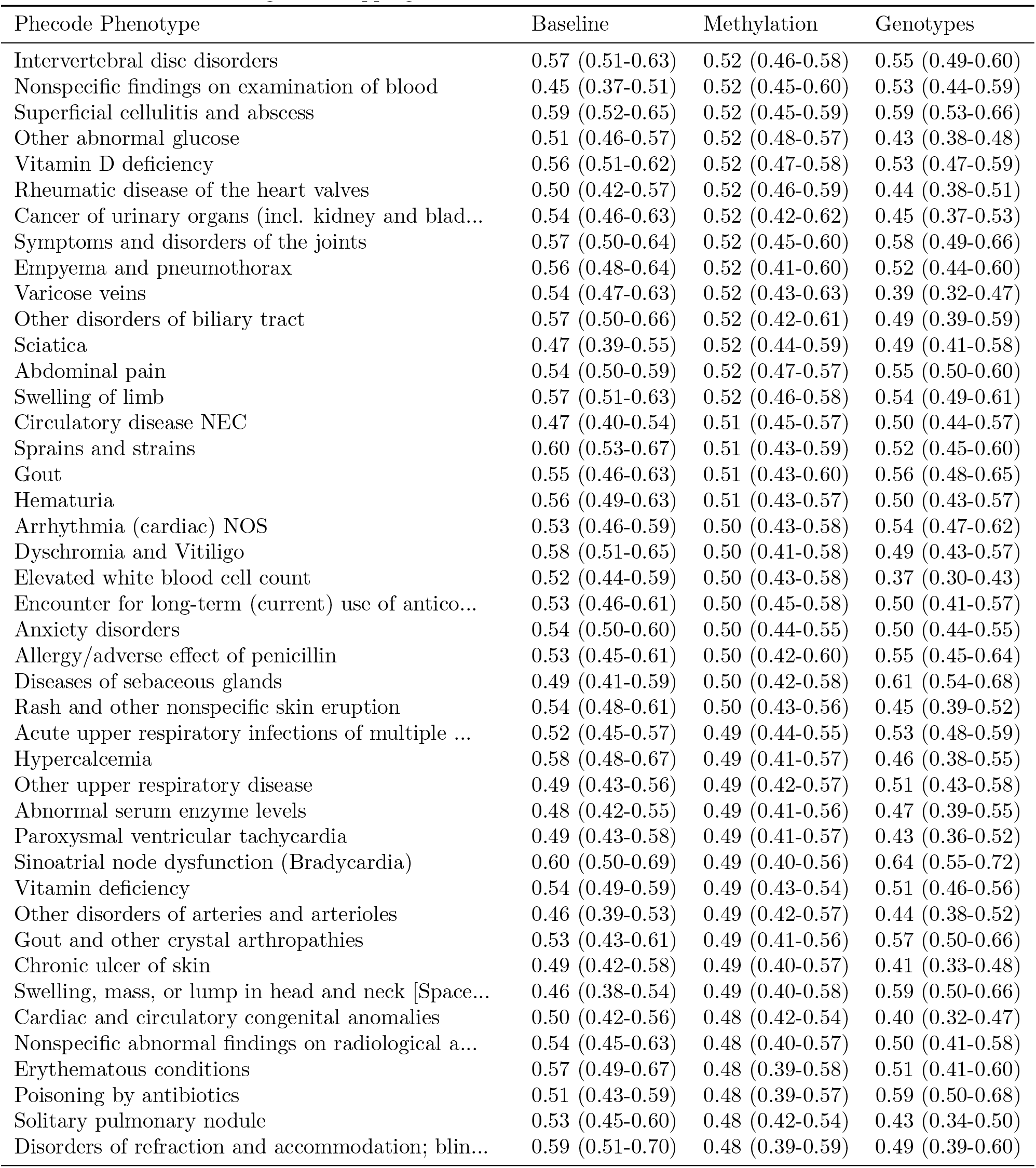
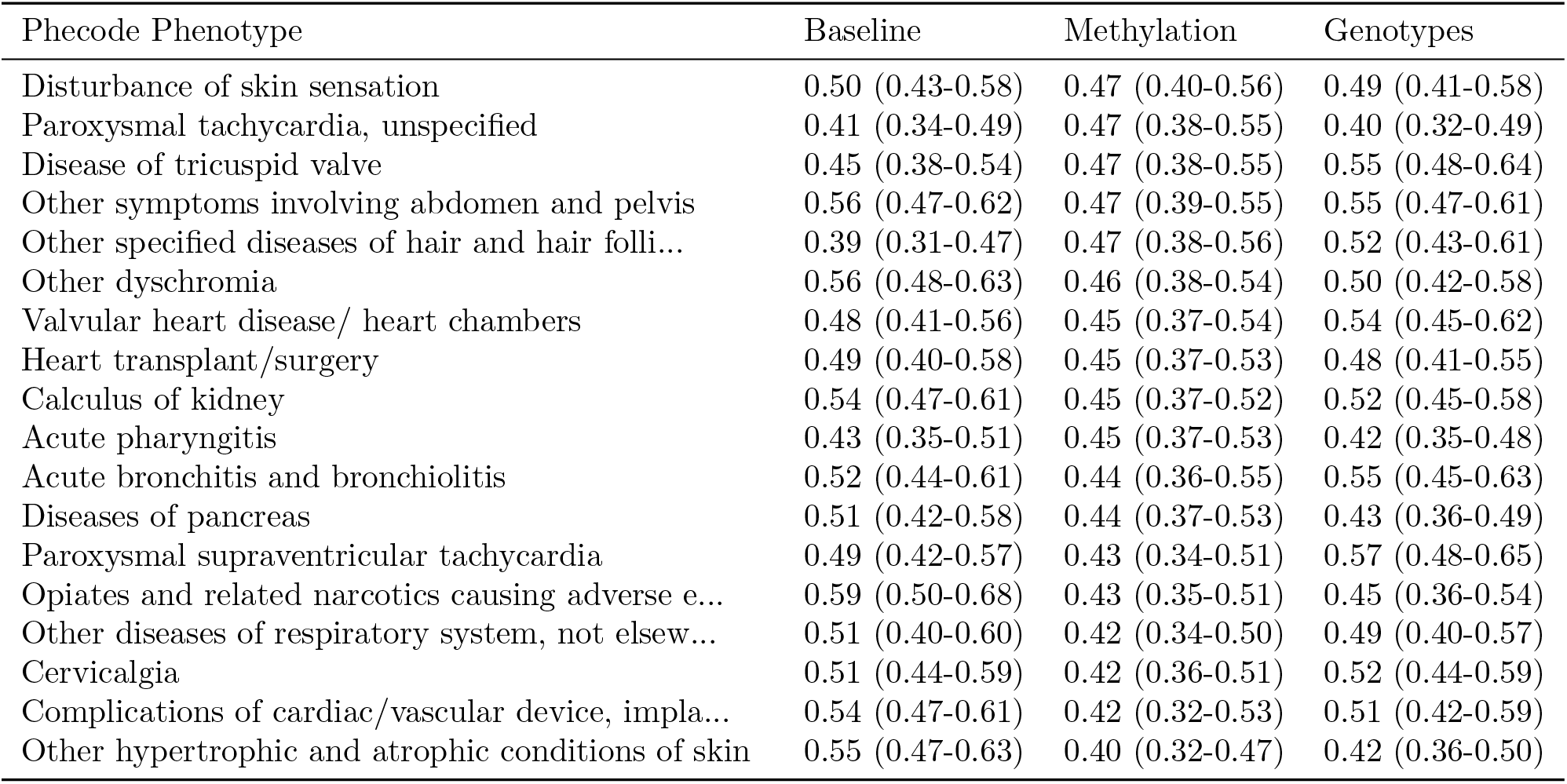
Mean (95% confidence interval) area under the ROC curve for predicting patient diagnoses, grouped into Phecode phenotypes, using the baseline, methylation data, and genotype data. Confidence intervals determined using bootstrapping.

**Table S5.**
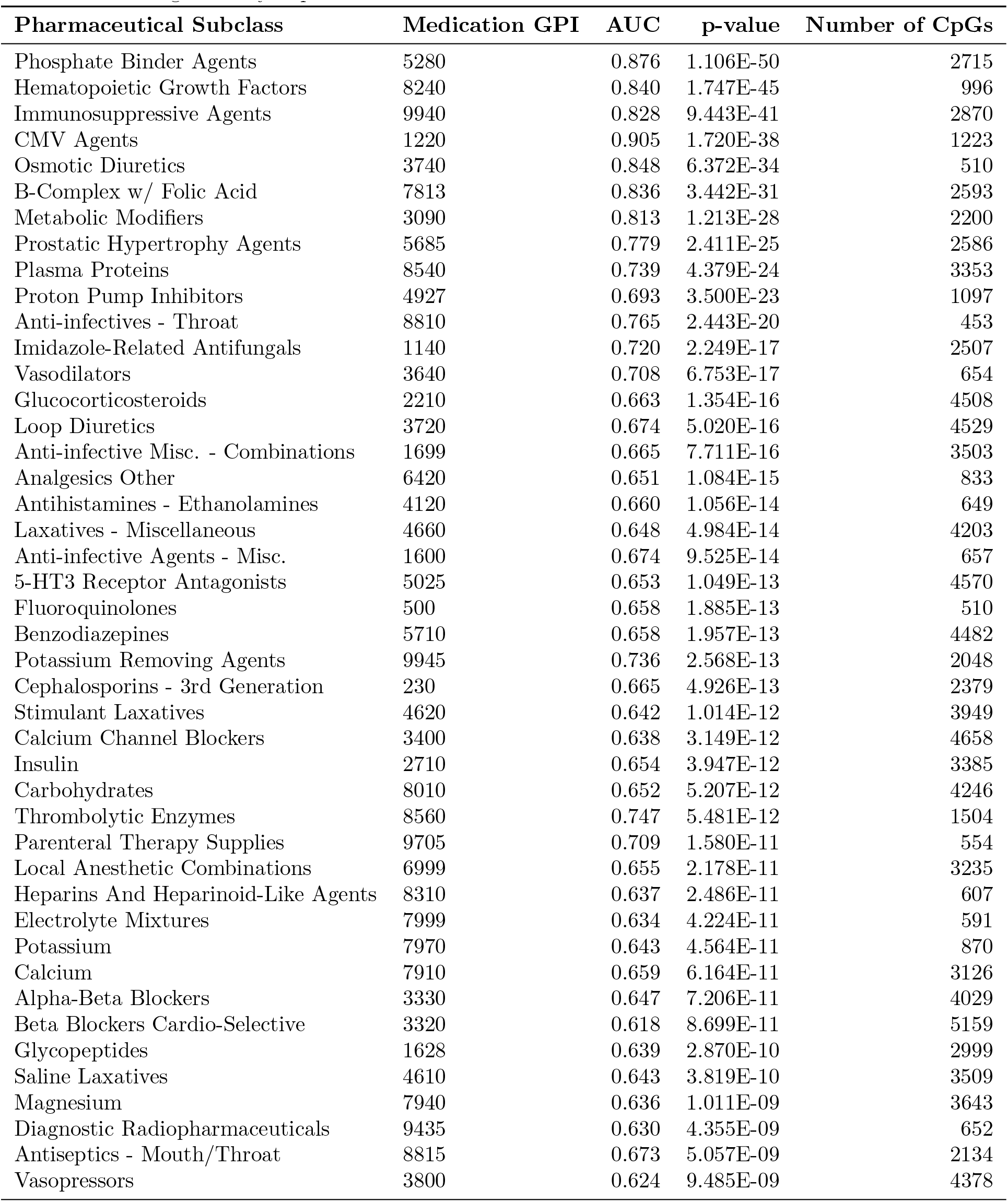
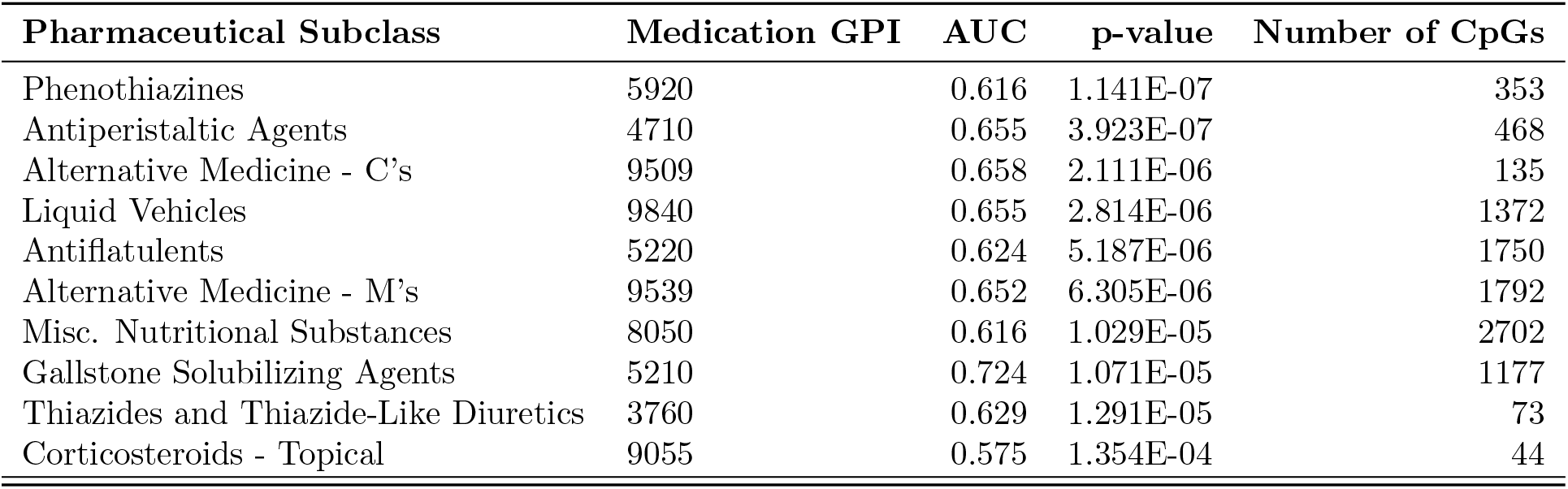
The imputation accuracy, p-value and number of CpGs selected for signi_cantly imputed MRS that also signi_cantly improved over the baseline model.

| Pharmaceutical Subclass | Medication GPI | AUC | p-value | Number of CpGs |
| --- | --- | --- | --- | --- |
| Phosphate Binder Agents | 5280 | 0.876 | 1.106E-50 | 2715 |
| Hematopoietic Growth Factors | 8240 | 0.840 | 1.747E-45 | 996 |
| Immunosuppressive Agents | 9940 | 0.828 | 9.443E-41 | 2870 |
| CMV Agents | 1220 | 0.905 | 1.720E-38 | 1223 |
| Osmotic Diuretics | 3740 | 0.848 | 6.372E-34 | 510 |
| B-Complex w/ Folic Acid | 7813 | 0.836 | 3.442E-31 | 2593 |
| Metabolic Modifiers | 3090 | 0.813 | 1.213E-28 | 2200 |
| Prostatic Hypertrophy Agents | 5685 | 0.779 | 2.411E-25 | 2586 |
| Plasma Proteins | 8540 | 0.739 | 4.379E-24 | 3353 |
| Proton Pump Inhibitors | 4927 | 0.693 | 3.500E-23 | 1097 |
| Anti-infectives - Throat | 8810 | 0.765 | 2.443E-20 | 453 |
| Imidazole-Related Antifungals | 1140 | 0.720 | 2.249E-17 | 2507 |
| Vasodilators | 3640 | 0.708 | 6.753E-17 | 654 |
| Glucocorticosteroids | 2210 | 0.663 | 1.354E-16 | 4508 |
| Loop Diuretics | 3720 | 0.674 | 5.020E-16 | 4529 |
| Anti-infective Misc. - Combinations | 1699 | 0.665 | 7.711E-16 | 3503 |
| Analgesics Other | 6420 | 0.651 | 1.084E-15 | 833 |
| Antihistamines - Ethanolamines | 4120 | 0.660 | 1.056E-14 | 649 |
| Laxatives - Miscellaneous | 4660 | 0.648 | 4.984E-14 | 4203 |
| Anti-infective Agents - Misc. | 1600 | 0.674 | 9.525E-14 | 657 |
| 5-HT3 Receptor Antagonists | 5025 | 0.653 | 1.049E-13 | 4570 |
| Fluoroquinolones | 500 | 0.658 | 1.885E-13 | 510 |
| Benzodiazepines | 5710 | 0.658 | 1.957E-13 | 4482 |
| Potassium Removing Agents | 9945 | 0.736 | 2.568E-13 | 2048 |
| Cephalosporins - 3rd Generation | 230 | 0.665 | 4.926E-13 | 2379 |
| Stimulant Laxatives | 4620 | 0.642 | 1.014E-12 | 3949 |
| Calcium Channel Blockers | 3400 | 0.638 | 3.149E-12 | 4658 |
| Insulin | 2710 | 0.654 | 3.947E-12 | 3385 |
| Carbohydrates | 8010 | 0.652 | 5.207E-12 | 4246 |
| Thrombolytic Enzymes | 8560 | 0.747 | 5.481E-12 | 1504 |
| Parenteral Therapy Supplies | 9705 | 0.709 | 1.580E-11 | 554 |
| Local Anesthetic Combinations | 6999 | 0.655 | 2.178E-11 | 3235 |
| Heparins And Heparinoid-Like Agents | 8310 | 0.637 | 2.486E-11 | 607 |
| Electrolyte Mixtures | 7999 | 0.634 | 4.224E-11 | 591 |
| Potassium | 7970 | 0.643 | 4.564E-11 | 870 |
| Calcium | 7910 | 0.659 | 6.164E-11 | 3126 |
| Alpha-Beta Blockers | 3330 | 0.647 | 7.206E-11 | 4029 |
| Beta Blockers Cardio-Selective | 3320 | 0.618 | 8.699E-11 | 5159 |
| Glycopeptides | 1628 | 0.639 | 2.870E-10 | 2999 |
| Saline Laxatives | 4610 | 0.643 | 3.819E-10 | 3509 |
| Magnesium | 7940 | 0.636 | 1.011E-09 | 3643 |
| Diagnostic Radiopharmaceuticals | 9435 | 0.630 | 4.355E-09 | 652 |
| Antiseptics - Mouth/Throat | 8815 | 0.673 | 5.057E-09 | 2134 |
| Vasopressors | 3800 | 0.624 | 9.485E-09 | 4378 |
| Phenothiazines | 5920 | 0.616 | 1.141E-07 | 353 |
| Antiperistaltic Agents | 4710 | 0.655 | 3.923E-07 | 468 |
| Alternative Medicine - C's | 9509 | 0.658 | 2.111E-06 | 135 |
| Liquid Vehicles | 9840 | 0.655 | 2.814E-06 | 1372 |
| Antiflatulents | 5220 | 0.624 | 5.187E-06 | 1750 |
| Alternative Medicine - M's | 9539 | 0.652 | 6.305E-06 | 1792 |
| Misc. Nutritional Substances | 8050 | 0.616 | 1.029E-05 | 2702 |
| Gallstone Solubilizing Agents | 5210 | 0.724 | 1.071E-05 | 1177 |
| Thiazides and Thiazide-Like Diuretics | 3760 | 0.629 | 1.291E-05 | 73 |
| Corticosteroids - Topical | 9055 | 0.575 | 1.354E-04 | 44 |

**Table S6.** The imputation accuracy, p-value and number of CpGs selected for significantly imputed MRS that also significantly improved over the baseline model.

| Lab | $R^2$ | p-value | Number of CpGs |
| --- | --- | --- | --- |
| Creatinine | 0.457 | 1.266E-95 | 32364 |
| Urea Nitrogen | 0.435 | 2.502E-87 | 4218 |
| Absolute Eos Count | 0.352 | 3.960E-51 | 3602 |
| Hemoglobin | 0.284 | 3.024E-50 | 25995 |
| Hematocrit | 0.246 | 1.144E-42 | 1827 |
| Neutrophil Percent, Auto | 0.264 | 1.139E-36 | 897 |
| Mean Corpuscular Hemoglobin | 0.208 | 7.037E-35 | 33151 |
| Mean Corpuscular Volume | 0.183 | 1.439E-30 | 60383 |
| Platelet Count, Auto | 0.168 | 1.956E-28 | 15362 |
| Chloride | 0.141 | 1.493E-24 | 44943 |
| Albumin | 0.143 | 6.472E-22 | 6020 |
| Absolute Immature Gran Count | 0.155 | 1.545E-20 | 1705 |
| White Blood Cell Count | 0.120 | 3.405E-20 | 29995 |
| Absolute Neut Count | 0.149 | 4.974E-20 | 19924 |
| Sodium | 0.115 | 5.060E-20 | 35598 |
| Absolute Lymphocyte Count | 0.128 | 2.920E-17 | 10575 |
| Absolute Mono Count | 0.121 | 2.579E-16 | 3675 |
| Neutrophils Abs (Prelim) | 0.116 | 1.263E-15 | 18192 |
| Glucose | 0.075 | 4.033E-13 | 973 |
| Absolute Baso Count | 0.086 | 7.020E-12 | 566 |
| Hgb A1c - Hplc | 0.125 | 2.588E-11 | 902 |
| Ferritin | 0.122 | 1.331E-07 | 40620 |
| Iron Binding Capacity | 0.104 | 8.842E-07 | 7121 |
| Qrs Duration | 0.052 | 1.722E-06 | 4731 |
| Anion Gap | 0.027 | 1.729E-05 | 29869 |
| Alanine Aminotransferase | 0.028 | 2.801E-05 | 546 |
| Cholesterol, Hdl | 0.054 | 4.612E-05 | 60 |
| Ventricular Rate | 0.033 | 1.499E-04 | 92303 |
| Bilirubin, Total | 0.022 | 3.094E-04 | 535 |

**Table S7.**
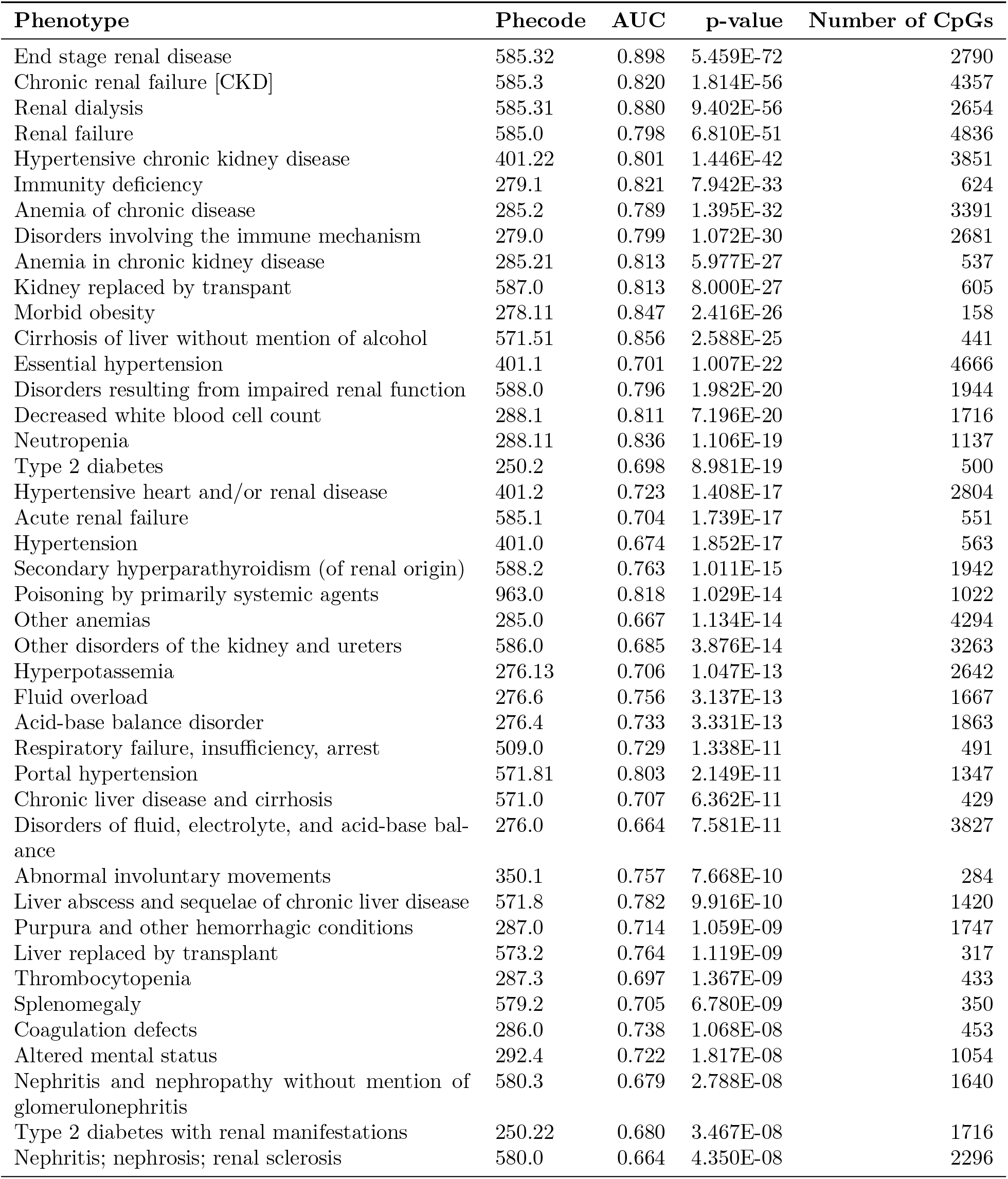
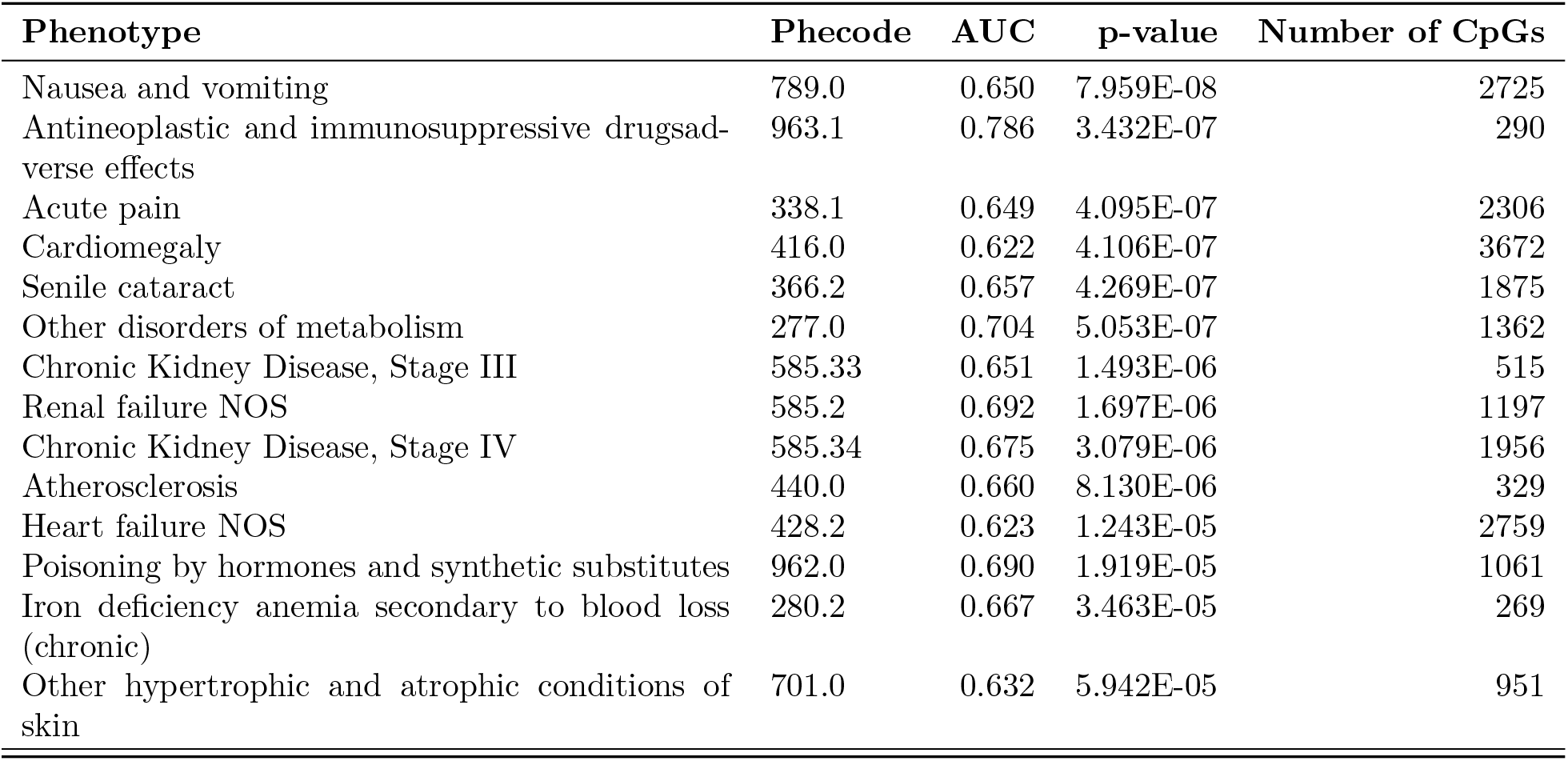
The imputation accuracy, p-value and number of CpGs selected for signi_cantly imputed MRS that also signi_cantly improved over the baseline model.

| Phenotype | Phecode | AUC | p-value | Number of CpGs |
| --- | --- | --- | --- | --- |
| End stage renal disease | 585.32 | 0.898 | 5.459E-72 | 2790 |
| Chronic renal failure [CKD] | 585.3 | 0.820 | 1.814E-56 | 4357 |
| Renal dialysis | 585.31 | 0.880 | 9.402E-56 | 2654 |
| Renal failure | 585.0 | 0.798 | 6.810E-51 | 4836 |
| Hypertensive chronic kidney disease | 401.22 | 0.801 | 1.446E-42 | 3851 |
| Immunity deficiency | 279.1 | 0.821 | 7.942E-33 | 624 |
| Anemia of chronic disease | 285.2 | 0.789 | 1.395E-32 | 3391 |
| Disorders involving the immune mechanism | 279.0 | 0.799 | 1.072E-30 | 2681 |
| Anemia in chronic kidney disease | 285.21 | 0.813 | 5.977E-27 | 537 |
| Kidney replaced by transpant | 587.0 | 0.813 | 8.000E-27 | 605 |
| Morbid obesity | 278.11 | 0.847 | 2.416E-26 | 158 |
| Cirrhosis of liver without mention of alcohol | 571.51 | 0.856 | 2.588E-25 | 441 |
| Essential hypertension | 401.1 | 0.701 | 1.007E-22 | 4666 |
| Disorders resulting from impaired renal function | 588.0 | 0.796 | 1.982E-20 | 1944 |
| Decreased white blood cell count | 288.1 | 0.811 | 7.196E-20 | 1716 |
| Neutropenia | 288.11 | 0.836 | 1.106E-19 | 1137 |
| Type 2 diabetes | 250.2 | 0.698 | 8.981E-19 | 500 |
| Hypertensive heart and/or renal disease | 401.2 | 0.723 | 1.408E-17 | 2804 |
| Acute renal failure | 585.1 | 0.704 | 1.739E-17 | 551 |
| Hypertension | 401.0 | 0.674 | 1.852E-17 | 563 |
| Secondary hyperparathyroidism (of renal origin) | 588.2 | 0.763 | 1.011E-15 | 1942 |
| Poisoning by primarily systemic agents | 963.0 | 0.818 | 1.029E-14 | 1022 |
| Other anemias | 285.0 | 0.667 | 1.134E-14 | 4294 |
| Other disorders of the kidney and ureters | 586.0 | 0.685 | 3.876E-14 | 3263 |
| Hyperpotassemia | 276.13 | 0.706 | 1.047E-13 | 2642 |
| Fluid overload | 276.6 | 0.756 | 3.137E-13 | 1667 |
| Acid-base balance disorder | 276.4 | 0.733 | 3.331E-13 | 1863 |
| Respiratory failure, insufficiency, arrest | 509.0 | 0.729 | 1.338E-11 | 491 |
| Portal hypertension | 571.81 | 0.803 | 2.149E-11 | 1347 |
| Chronic liver disease and cirrhosis | 571.0 | 0.707 | 6.362E-11 | 429 |
| Disorders of fluid, electrolyte, and acid-base balance | 276.0 | 0.664 | 7.581E-11 | 3827 |
| Abnormal involuntary movements | 350.1 | 0.757 | 7.668E-10 | 284 |
| Liver abscess and sequelae of chronic liver disease | 571.8 | 0.782 | 9.916E-10 | 1420 |
| Purpura and other hemorrhagic conditions | 287.0 | 0.714 | 1.059E-09 | 1747 |
| Liver replaced by transplant | 573.2 | 0.764 | 1.119E-09 | 317 |
| Thrombocytopenia | 287.3 | 0.697 | 1.367E-09 | 433 |
| Splenomegaly | 579.2 | 0.705 | 6.780E-09 | 350 |
| Coagulation defects | 286.0 | 0.738 | 1.068E-08 | 453 |
| Altered mental status | 292.4 | 0.722 | 1.817E-08 | 1054 |
| Nephritis and nephropathy without mention of glomerulonephritis | 580.3 | 0.679 | 2.788E-08 | 1640 |
| Type 2 diabetes with renal manifestations | 250.22 | 0.680 | 3.467E-08 | 1716 |
| Nephritis; nephrosis; renal sclerosis | 580.0 | 0.664 | 4.350E-08 | 2296 |
| Nausea and vomiting | 789.0 | 0.650 | 7.959E-08 | 2725 |
| Antineoplastic and immunosuppressive drugsad-verse effects | 963.1 | 0.786 | 3.432E-07 | 290 |
| Acute pain | 338.1 | 0.649 | 4.095E-07 | 2306 |
| Cardiomegaly | 416.0 | 0.622 | 4.106E-07 | 3672 |
| Senile cataract | 366.2 | 0.657 | 4.269E-07 | 1875 |
| Other disorders of metabolism | 277.0 | 0.704 | 5.053E-07 | 1362 |
| Chronic Kidney Disease, Stage III | 585.33 | 0.651 | 1.493E-06 | 515 |
| Renal failure NOS | 585.2 | 0.692 | 1.697E-06 | 1197 |
| Chronic Kidney Disease, Stage IV | 585.34 | 0.675 | 3.079E-06 | 1956 |
| Atherosclerosis | 440.0 | 0.660 | 8.130E-06 | 329 |
| Heart failure NOS | 428.2 | 0.623 | 1.243E-05 | 2759 |
| Poisoning by hormones and synthetic substitutes | 962.0 | 0.690 | 1.919E-05 | 1061 |
| Iron deficiency anemia secondary to blood loss (chronic) | 280.2 | 0.667 | 3.463E-05 | 269 |
| Other hypertrophic and atrophic conditions of skin | 701.0 | 0.632 | 5.942E-05 | 951 |

**Table S8.** Number of samples with reported usage of medications in the pharmaceutical subclasses. Pharmaceutical subclasses are sorted by number of samples.

| Pharmaceutical Subclass | Number of Samples (Percent) |
| --- | --- |
| Sodium | 699 (80.9%) |
| Opioid Agonists | 639 (74.0%) |
| Local Anesthetics - Amides | 589 (68.2%) |
| Non-Barbiturate Hypnotics | 584 (67.6%) |
| 5-HT<sub>3</sub> Receptor Antagonists | 549 (63.5%) |
| Analgesics Other | 544 (63.0%) |
| Radiographic Contrast Media | 535 (61.9%) |
| Anesthetics - Misc. | 507 (58.7%) |
| Glucocorticosteroids | 499 (57.8%) |
| Salicylates | 459 (53.1%) |
| Heparins And Heparinoid-Like Agents | 459 (53.1%) |
| Opioid Combinations | 458 (53.0%) |
| HMG CoA Reductase Inhibitors | 456 (52.8%) |
| Proton Pump Inhibitors | 443 (51.3%) |
| Oil Soluble Vitamins | 434 (50.2%) |
| Vasopressors | 421 (48.7%) |
| Surfactant Laxatives | 398 (46.1%) |
| Electrolyte Mixtures | 390 (45.1%) |
| Antiarrhythmics Type I-B | 383 (44.3%) |
| Beta Blockers Cardio-Selective | 383 (44.3%) |
| Cephalosporins - 1st Generation | 369 (42.7%) |
| Calcium Channel Blockers | 367 (42.5%) |
| Loop Diuretics | 346 (40.0%) |
| Miscellaneous Contrast Media | 341 (39.5%) |
| Nondepolarizing Muscle Relaxants | 336 (38.9%) |
| Fluoroquinolones | 327 (37.8%) |
| Stimulant Laxatives | 326 (37.7%) |
| Nonsteroidal Anti-inflammatory Agents (NSAIDs) | 313 (36.2%) |
| Sympathomimetics | 308 (35.6%) |
| Antihistamines - Ethanolamines | 301 (34.8%) |
| Laxatives - Miscellaneous | 293 (33.9%) |
| Magnesium | 290 (33.6%) |
| Local Anesthetics - Topical | 280 (32.4%) |
| Potassium | 277 (32.1%) |
| Insulin | 269 (31.1%) |
| Benzodiazepines | 265 (30.7%) |
| Diagnostic Radiopharmaceuticals | 264 (30.6%) |
| Anticonvulsants - Misc. | 260 (30.1%) |
| Carbohydrates | 252 (29.2%) |
| Saline Laxatives | 250 (28.9%) |
| Antispasmodics | 250 (28.9%) |
| H<sub>2</sub> Antagonists | 232 (26.9%) |
| Angiotensin II Receptor Antagonists | 231 (26.7%) |
| ACE Inhibitors | 225 (26.0%) |
| Penicillin Combinations | 219 (25.3%) |
| Cephalosporins - 3rd Generation | 217 (25.1%) |
| Nitrates | 215 (24.9%) |
| Glycopeptides | 213 (24.7%) |
| Alpha-Beta Blockers | 210 (24.3%) |
| Calcium | 207 (24.0%) |
| Multivitamins | 207 (24.0%) |
| Local Anesthetic Combinations | 200 (23.1%) |
| Anti-infective Misc. - Combinations | 198 (22.9%) |
| Anti-infective Agents - Misc. | 193 (22.3%) |
| Plasma Proteins | 190 (22.0%) |
| Diagnostic Drugs | 189 (21.9%) |
| Water Soluble Vitamins | 188 (21.8%) |
| Phenothiazines | 184 (21.3%) |
| Gastrointestinal Stimulants | 182 (21.1%) |
| Corticosteroids - Topical | 182 (21.1%) |
| Central Muscle Relaxants | 181 (20.9%) |
| Viral Vaccines | 175 (20.3%) |
| Iron | 170 (19.7%) |
| Vasodilators | 168 (19.4%) |
| Antibiotics - Topical | 166 (19.2%) |
| Hematopoietic Growth Factors | 165 (19.1%) |
| Azithromycin | 164 (19.0%) |
| Antacids - Calcium Salts | 164 (19.0%) |
| Antimyasthenic/Cholinergic Agents | 158 (18.3%) |
| Nasal Steroids | 158 (18.3%) |
| Selective Serotonin Reuptake Inhibitors (SSRIs) | 157 (18.2%) |
| Thiazides and Thiazide-Like Diuretics | 157 (18.2%) |
| Misc. Nutritional Substances | 155 (17.9%) |
| Opioid Antagonists | 155 (17.9%) |
| Platelet Aggregation Inhibitors | 154 (17.8%) |
| Thyroid Hormones | 149 (17.2%) |
| Antifungals - Topical | 149 (17.2%) |
| Bacterial Vaccines | 144 (16.7%) |
| Immunosuppressive Agents | 142 (16.4%) |
| Phosphate Binder Agents | 140 (16.2%) |
| Serotonin Modulators | 136 (15.7%) |
| Laxative Combinations | 136 (15.7%) |
| Biguanides | 135 (15.6%) |
| Depolarizing Muscle Relaxants | 135 (15.6%) |
| Genitourinary Irrigants | 134 (15.5%) |
| Prostatic Hypertrophy Agents | 134 (15.5%) |
| Bronchodilators - Anticholinergics | 131 (15.2%) |
| Antiflatulents | 130 (15.0%) |
| Antacid Combinations | 127 (14.7%) |
| Aminopenicillins | 126 (14.6%) |
| Imidazole-Related Antifungals | 125 (14.5%) |
| Diagnostic Tests | 120 (13.9%) |
| Cobalamins | 118 (13.7%) |
| Folic Acid/Folates | 116 (13.4%) |
| B-Complex w/ Folic Acid | 116 (13.4%) |
| Antihistamines - Non-Sedating | 113 (13.1%) |
| Anesthetics Topical Oral | 108 (12.5%) |
| Diabetic Supplies | 107 (12.4%) |
| Osmotic Diuretics | 106 (12.3%) |
| Tetracyclines | 105 (12.2%) |
| Multiple Vitamins w/ Minerals | 105 (12.2%) |
| Ophthalmic Anti-infectives | 104 (12.0%) |
| Metabolic Modifiers | 102 (11.8%) |
| Potassium Removing Agents | 102 (11.8%) |
| Potassium Sparing Diuretics | 101 (11.7%) |
| Hemostatics - Topical | 101 (11.7%) |
| Ophthalmics - Misc. | 101 (11.7%) |
| Gout Agents | 100 (11.6%) |
| Alternative Medicine - M's | 99 (11.5%) |
| Parenteral Therapy Supplies | 99 (11.5%) |
| Cough/Cold/Allergy Combinations | 99 (11.5%) |
| Antiseptics - Mouth/Throat | 98 (11.3%) |
| Direct Factor Xa Inhibitors | 97 (11.2%) |
| Anti-infectives - Throat | 94 (10.9%) |
| Anti-inflammatory Agents - Topical | 93 (10.8%) |
| Coumarin Anticoagulants | 92 (10.6%) |
| Posterior Pituitary Hormones | 91 (10.5%) |
| Antidotes and Specific Antagonists | 90 (10.4%) |
| Antiadrenergic Antihypertensives | 90 (10.4%) |
| Ophthalmic Steroids | 90 (10.4%) |
| Antitussives | 88 (10.2%) |
| Lincosamides | 84 (9.7%) |
| Dibenzapines | 83 (9.6%) |
| Bone Density Regulators | 81 (9.4%) |
| Antianxiety Agents - Misc. | 80 (9.3%) |
| Phosphate | 78 (9.0%) |
| Antiemetics - Anticholinergic | 77 (8.9%) |
| Antiperistaltic Agents | 76 (8.8%) |
| Herpes Agents | 76 (8.8%) |
| Bicarbonates | 75 (8.7%) |
| Liquid Vehicles | 72 (8.3%) |
| Antiarrhythmics Type III | 72 (8.3%) |
| Artificial Tears and Lubricants | 71 (8.2%) |
| Antidiarrheal/Probiotic Agents - Misc. | 71 (8.2%) |
| Toxoid Combinations | 70 (8.1%) |
| Urinary Antispasmodic - Antimuscarinics (Antich... | 67 (7.8%) |
| Lozenges | 67 (7.8%) |
| CMV Agents | 66 (7.6%) |
| Thrombolytic Enzymes | 66 (7.6%) |
| Impotence Agents | 65 (7.5%) |
| Alternative Medicine - C's | 64 (7.4%) |
| Sulfonylureas | 63 (7.3%) |
| Antihypertensive Combinations | 63 (7.3%) |
| Specialty Vitamins Products | 63 (7.3%) |
| Aminoglycosides | 61 (7.1%) |
| Cephalosporins - 2nd Generation | 60 (6.9%) |
| Alkalinizers | 59 (6.8%) |
| Opioid Partial Agonists | 73 (6.8%) |
| Urinary Anti-infectives | 58 (6.7%) |
| Irrigation Solutions | 58 (6.7%) |
| Influenza Agents | 57 (6.6%) |
| Expectorants | 57 (6.6%) |
| Beta Blockers Non-Selective | 56 (6.5%) |
| Tricyclic Agents | 56 (6.5%) |
| Serotonin-Norepinephrine Reuptake Inhibitors (S... | 56 (6.5%) |
| Cephalosporins - 4th Generation | 55 (6.4%) |
| Antihistamines-Topical | 55 (6.4%) |
| Antacids - Bicarbonate | 54 (6.2%) |
| Bulk Laxatives | 53 (6.1%) |
| Alpha-2 Receptor Antagonists (Tetracyclics) | 52 (6.0%) |
| Ophthalmic Local Anesthetics | 49 (5.7%) |
| Hemostatics - Systemic | 49 (5.7%) |
| Zinc | 48 (5.6%) |
| Dipeptidyl Peptidase-4 (DPP-4) Inhibitors | 47 (5.4%) |
| Gallstone Solubilizing Agents | 47 (5.4%) |
| Cycloplegic Mydriatics | 47 (5.4%) |
| Protamine | 58 (5.4%) |
| Butyrophenones | 46 (5.3%) |
| Antidepressants - Misc. | 45 (5.2%) |
| Mucolytics | 45 (5.2%) |
| Leukotriene Modulators | 44 (5.1%) |
| B-Complex Vitamins | 44 (5.1%) |
| Acne Products | 44 (5.1%) |

**Table S9.**
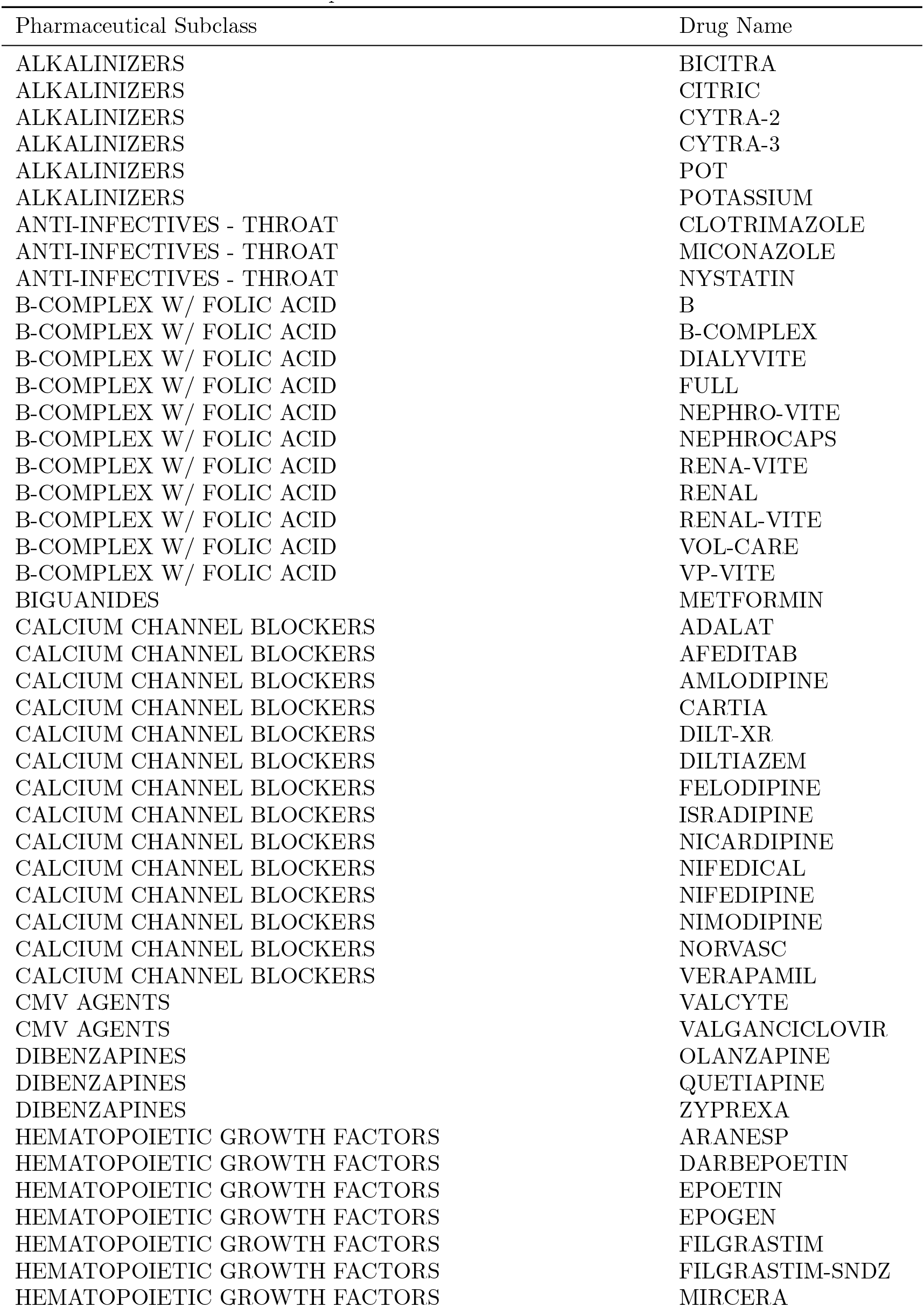
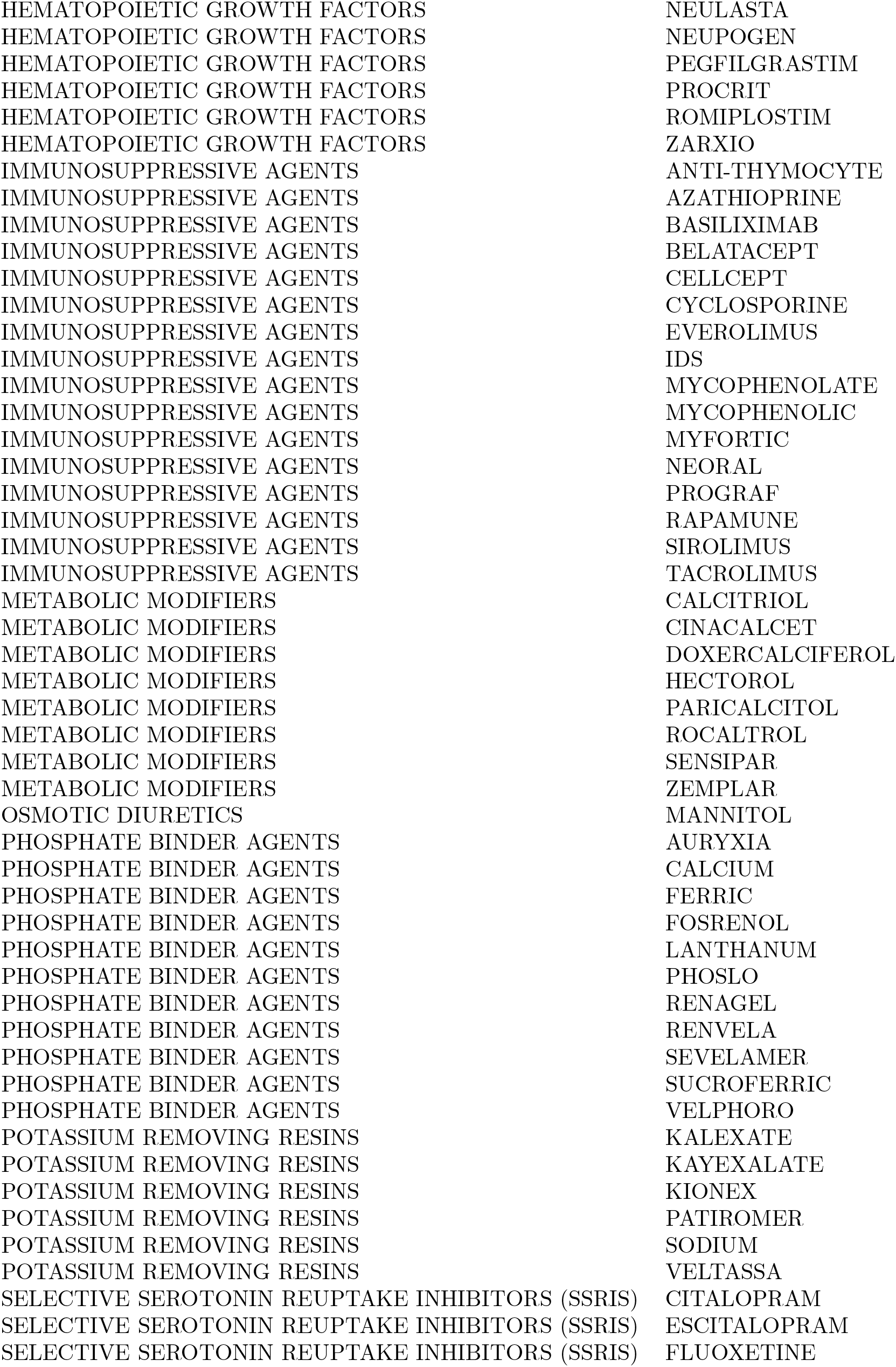
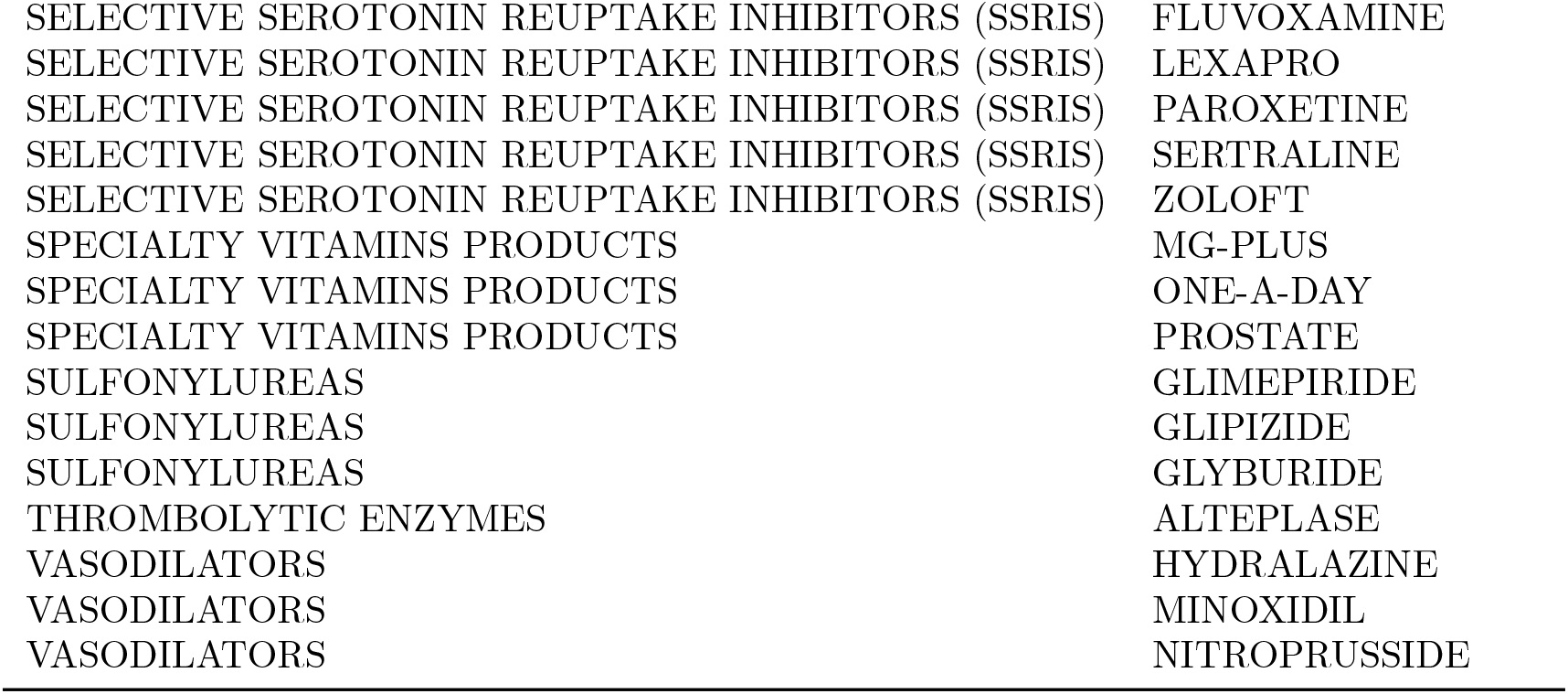
Medications used in each pharmaceutical subclass

**Figure S6.**
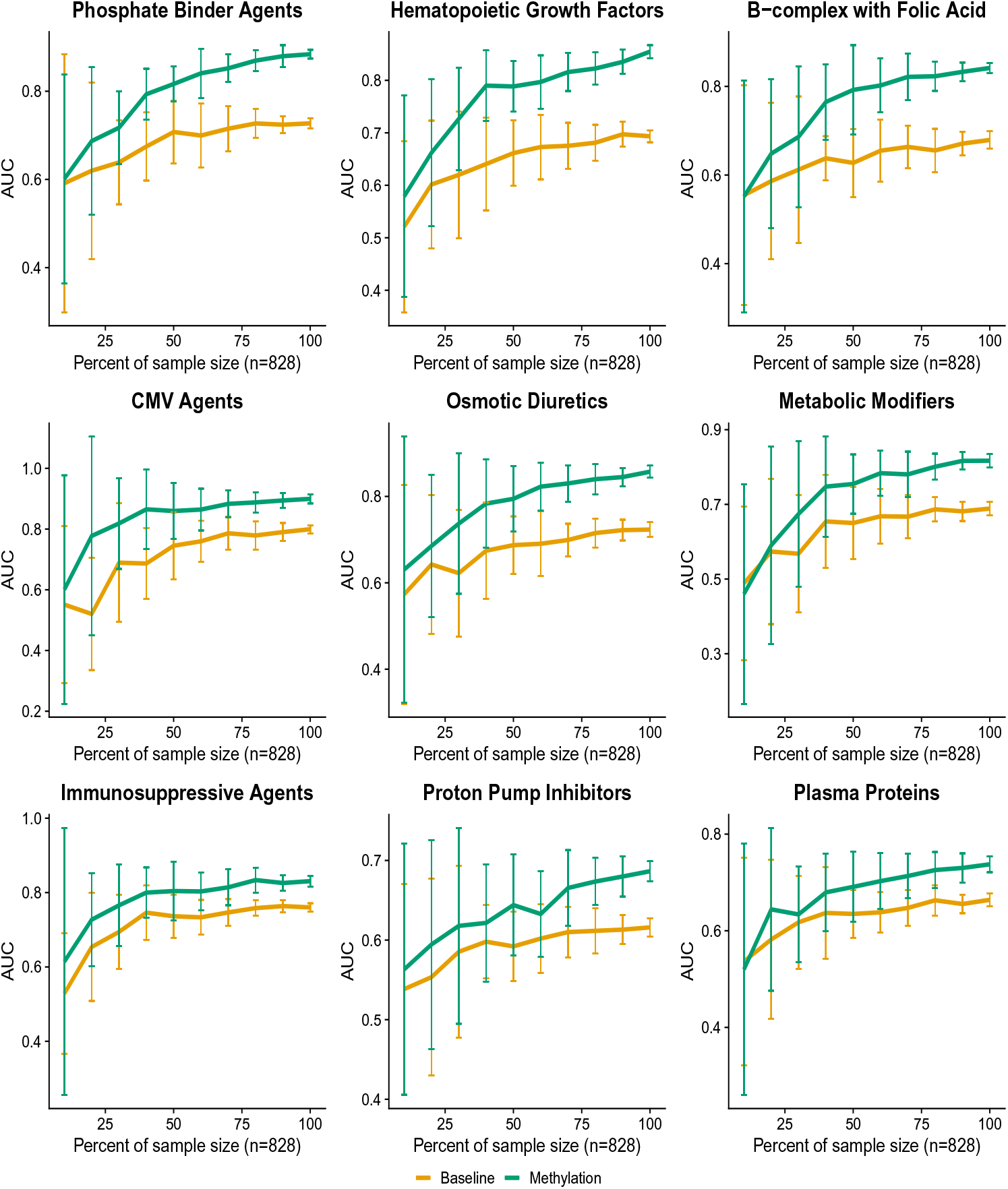
Downsampled performance on additional medications. We extended the downsampling experiments to include the top 10 most accurately imputed medications that also offered significant predictive power over the baseline features. We include here the remaining 9 medications.

**Figure S7.**
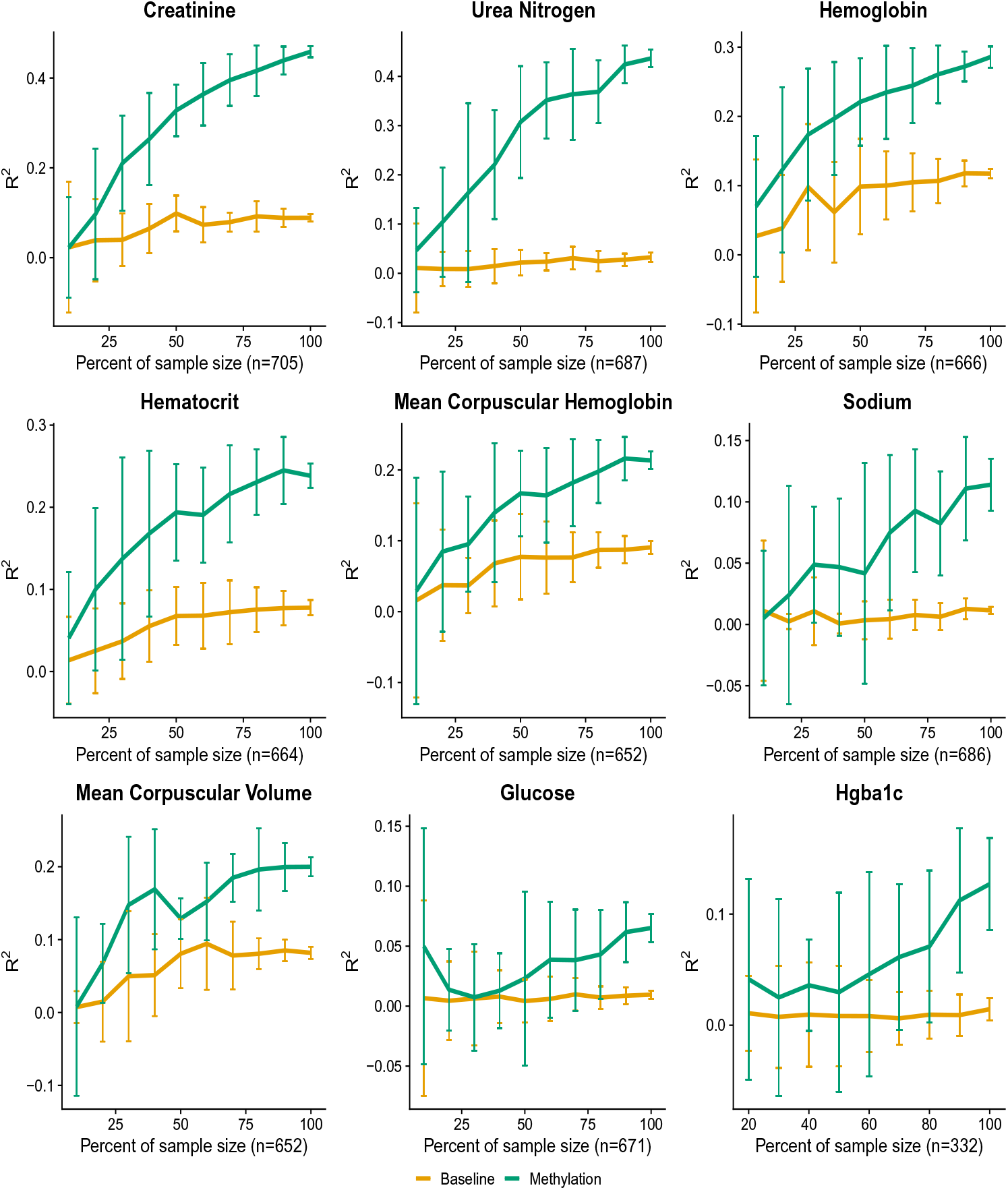
Downsampled performance on additional labs. We extended the downsampling experiments to include the top 10 most accurately imputed labs that also offered significant predictive power over the baseline features. We include here the remaining 9 labs.

**Figure S8.**
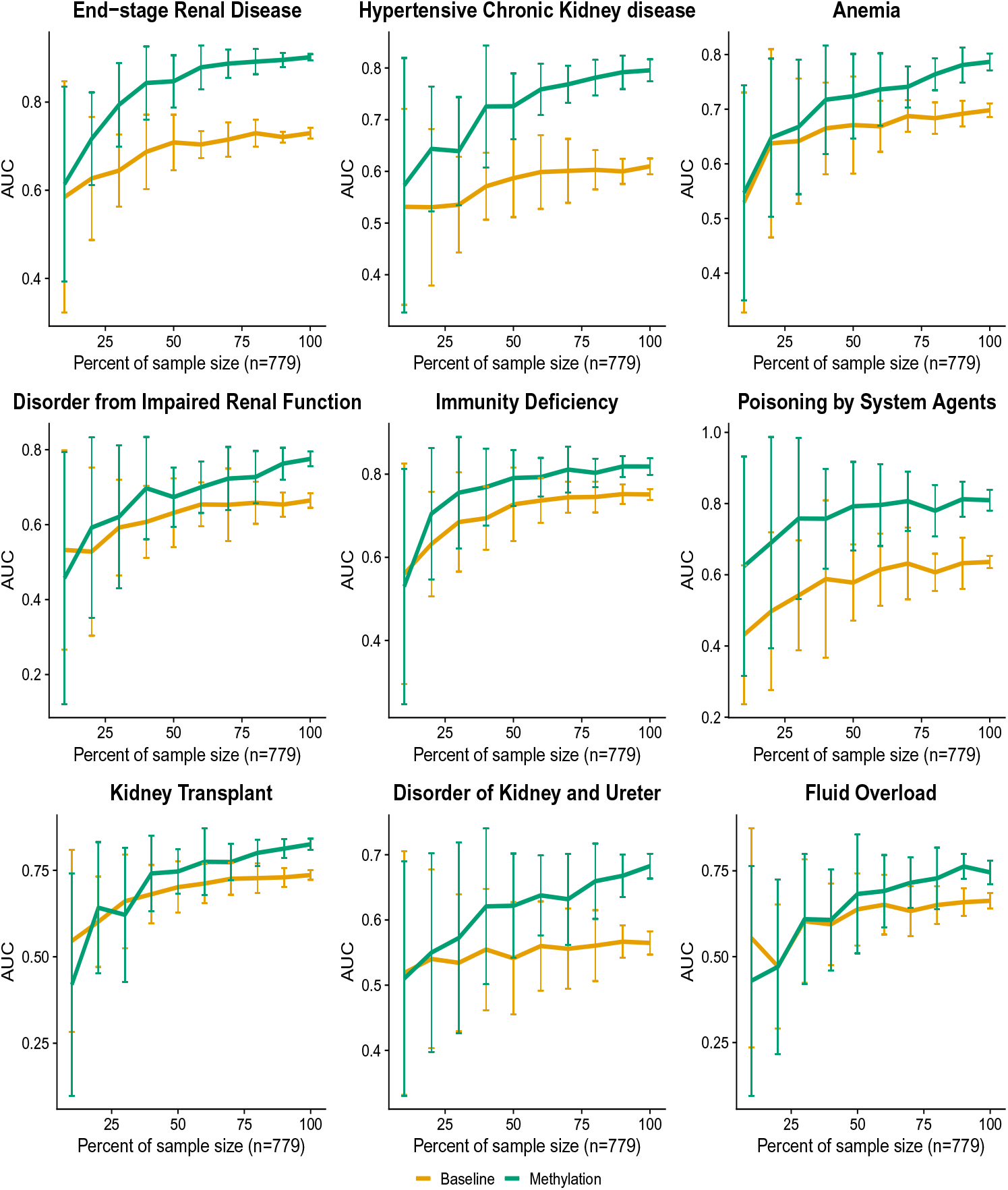
Downsampled performance on additional Phecodes. We extended the downsampling experiments to include the top 10 most accurately imputed Phecodes that also offered significant predictive power over the baseline features. We include here the remaining 9 Phecodes.

**Table S10.**
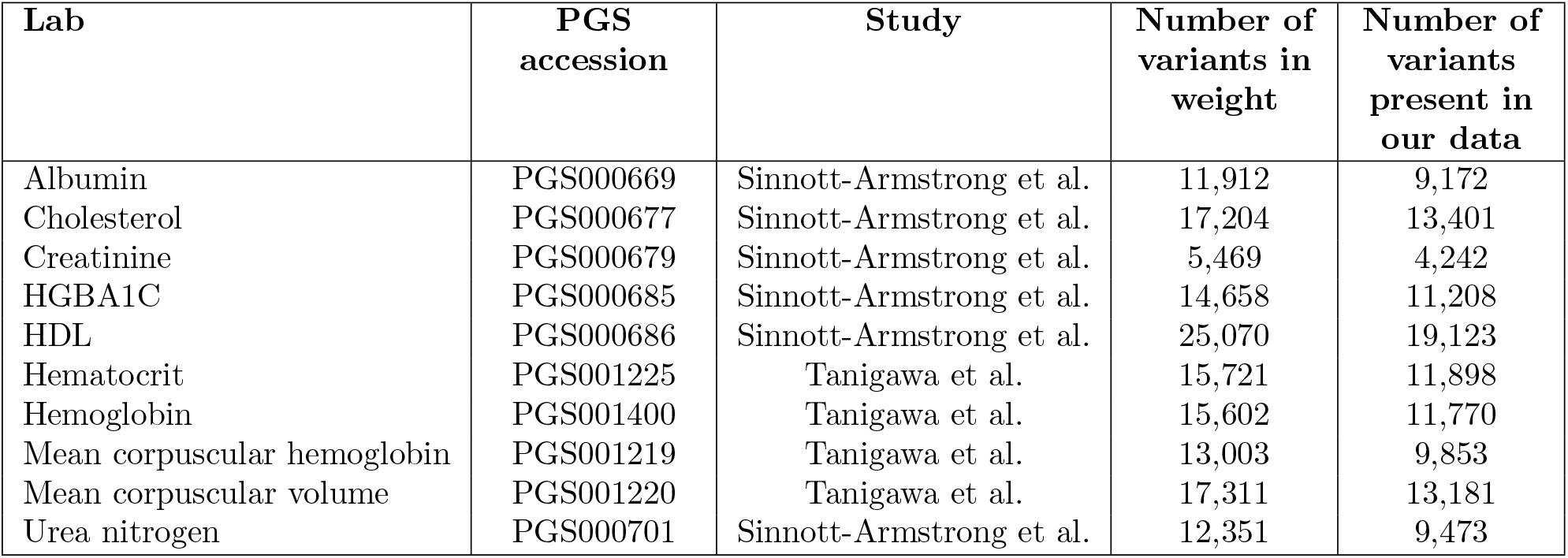
Polygenic scores used for the imputed genotypes. We list below the weights used for computing the polygenic risk scores. We downloaded the weights from the Polygenic Score Catalogue (PGS)[55] from two studies of the UKBiobank [51, 53].

**Figure S9.**
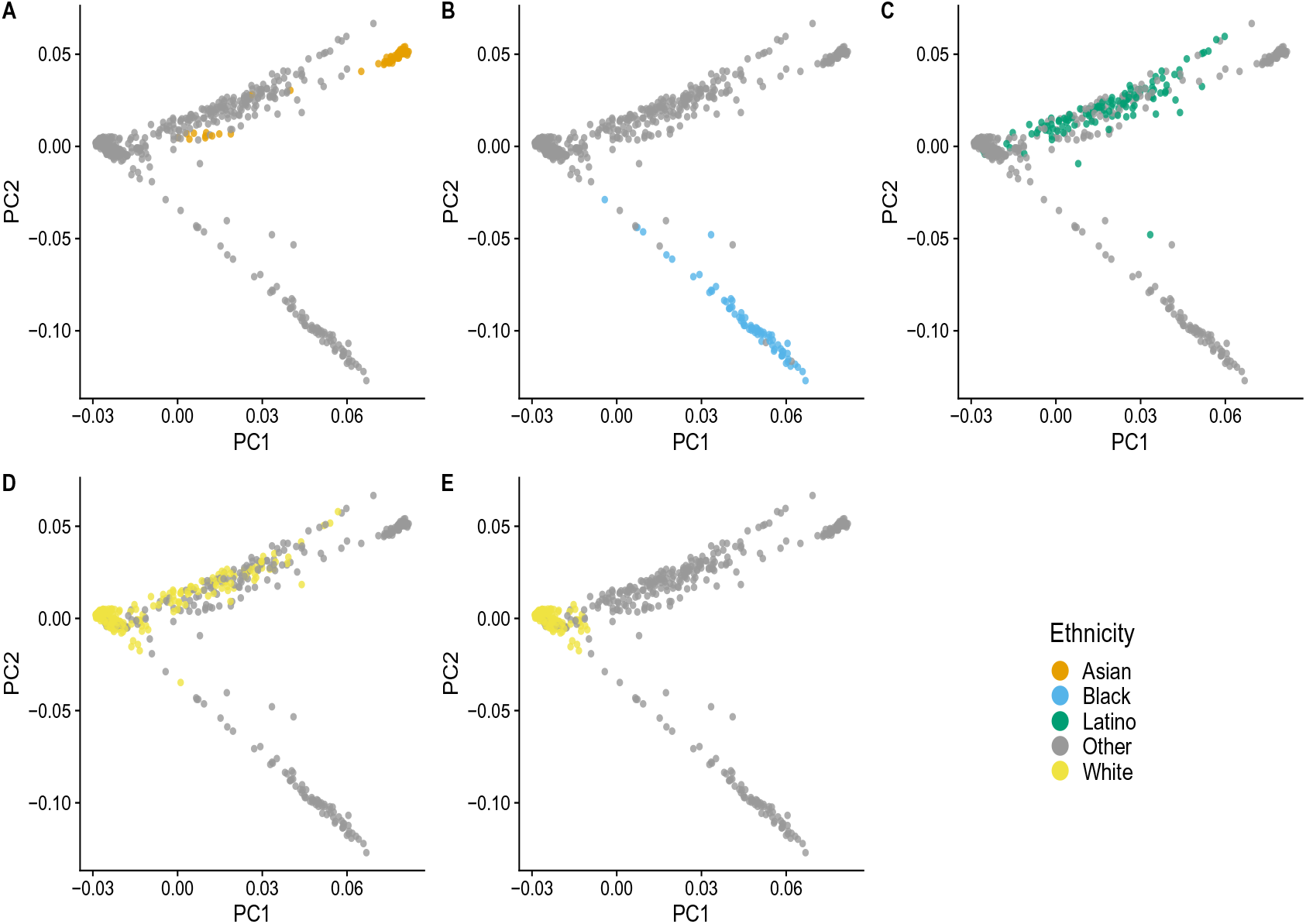
Self-reported ancestry along genetic PCs. We show the primary self-identified ethnicity in each plot individually. For the analysis using external PRS we limited the set of white-identifying individuals to those who additionally had a PC1 score of *− < .*01. We show the individuals used in our analysis in plot E.

**Figure S10.**
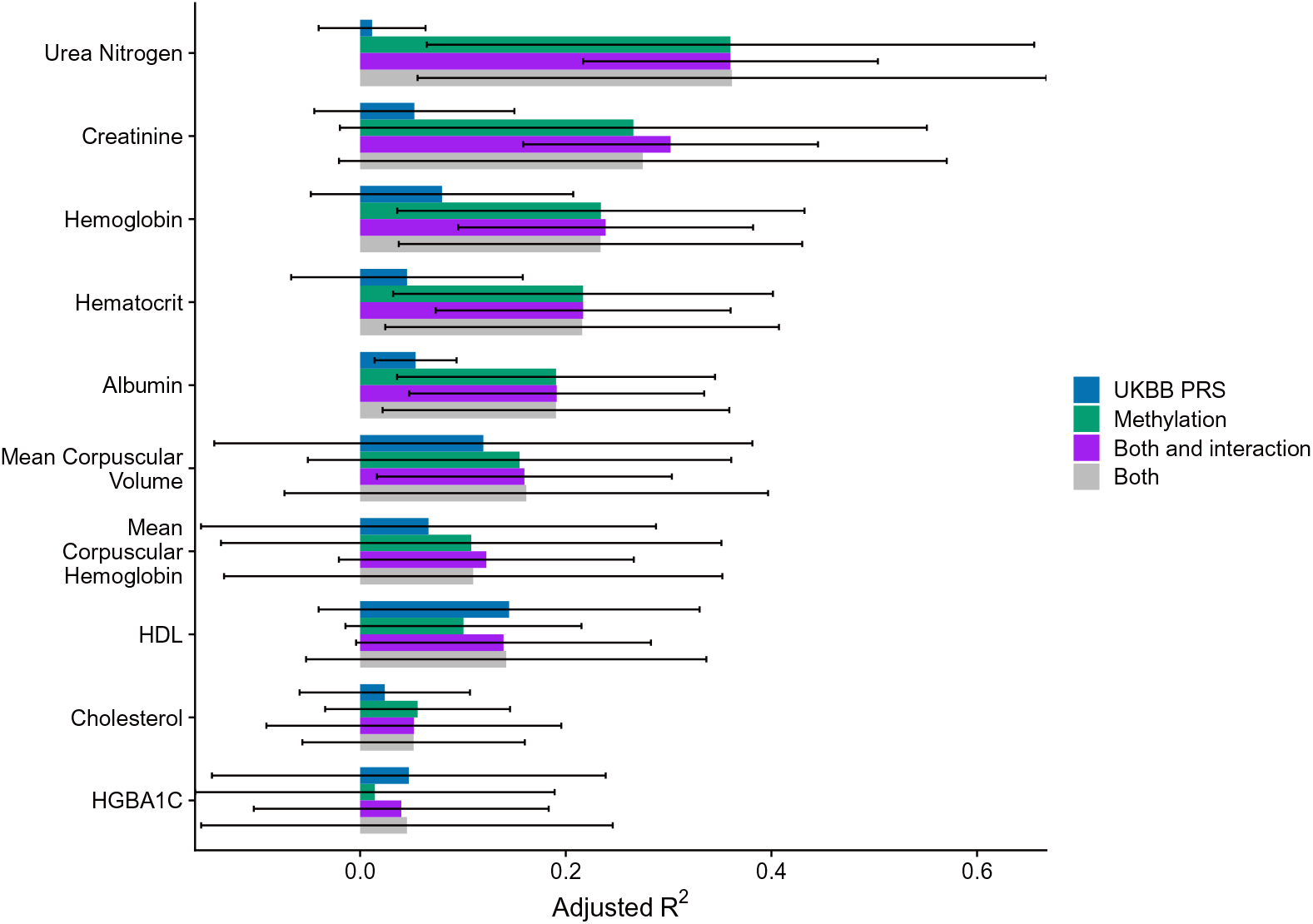
Labs as predicted by methylation, an externally-trained polygenic risk score, both, and a model that includes both as well as their interaction. The cross-validated adjusted *R*^2^ between the true and imputed lab value on 541 unrelated patients of non-Hispanic-Latino white-identifying individuals using predictors that leveraged baseline features with either an MRS, a PRS externally-trained from the UKBiobank, both the MRS and the PRS, or a model that used both as well as the interaction between the MRS and PRS. Creatinine was the only outcome for which the interaction between both risk scores was statistically significant (p=9.16e-05), however the interaction for mean corpuscular hemoglobin was nominally significant (p=1.44e-02). HDL corresponds to high-density lipoprotein cholesterol and HGBA1C to glycated hemoglobin.

**Figure S11.**
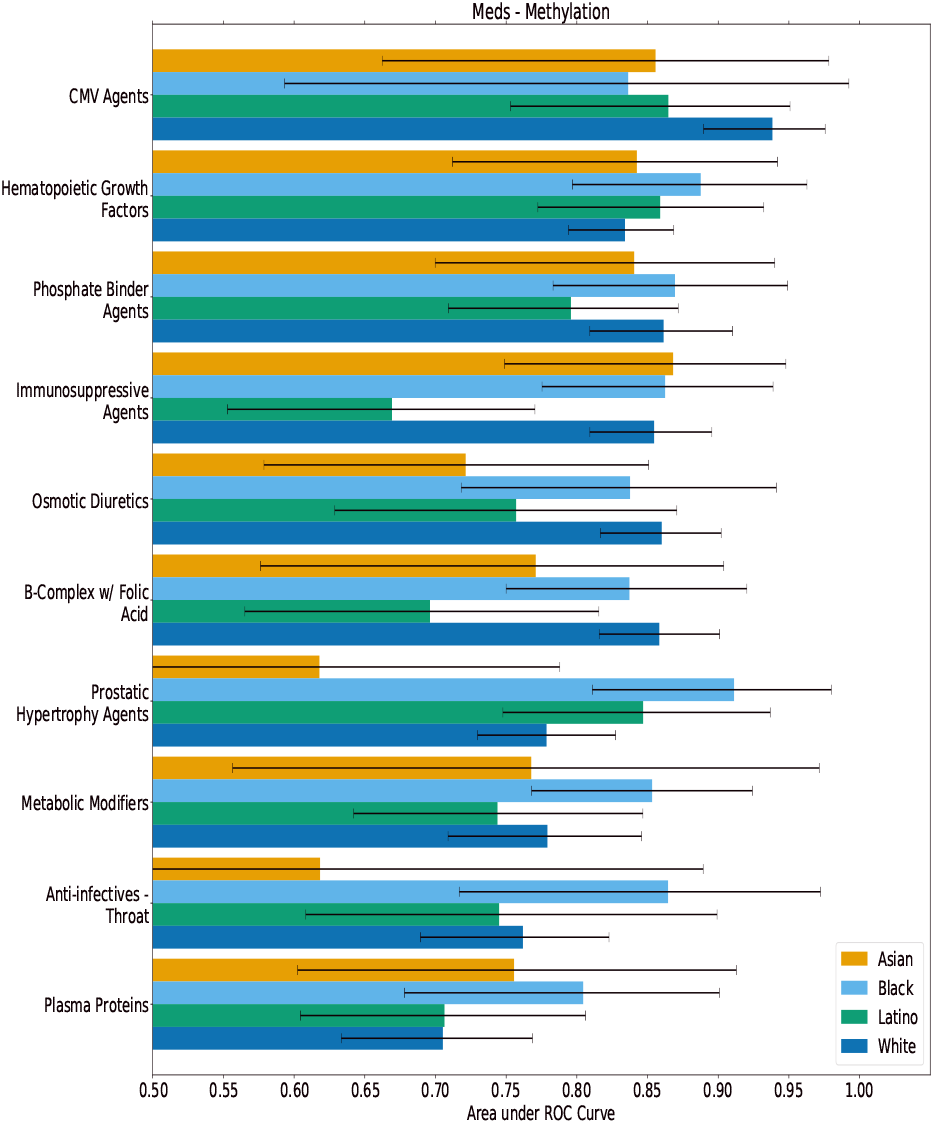
Best methylation-predicted medications within ancestral populations. After training a model on the entire heterogeneous set of individuals, we evaluated the predictive performance within each population separately. We observed no significant differences within self-reported ancestral groupings.

**Figure S12.**
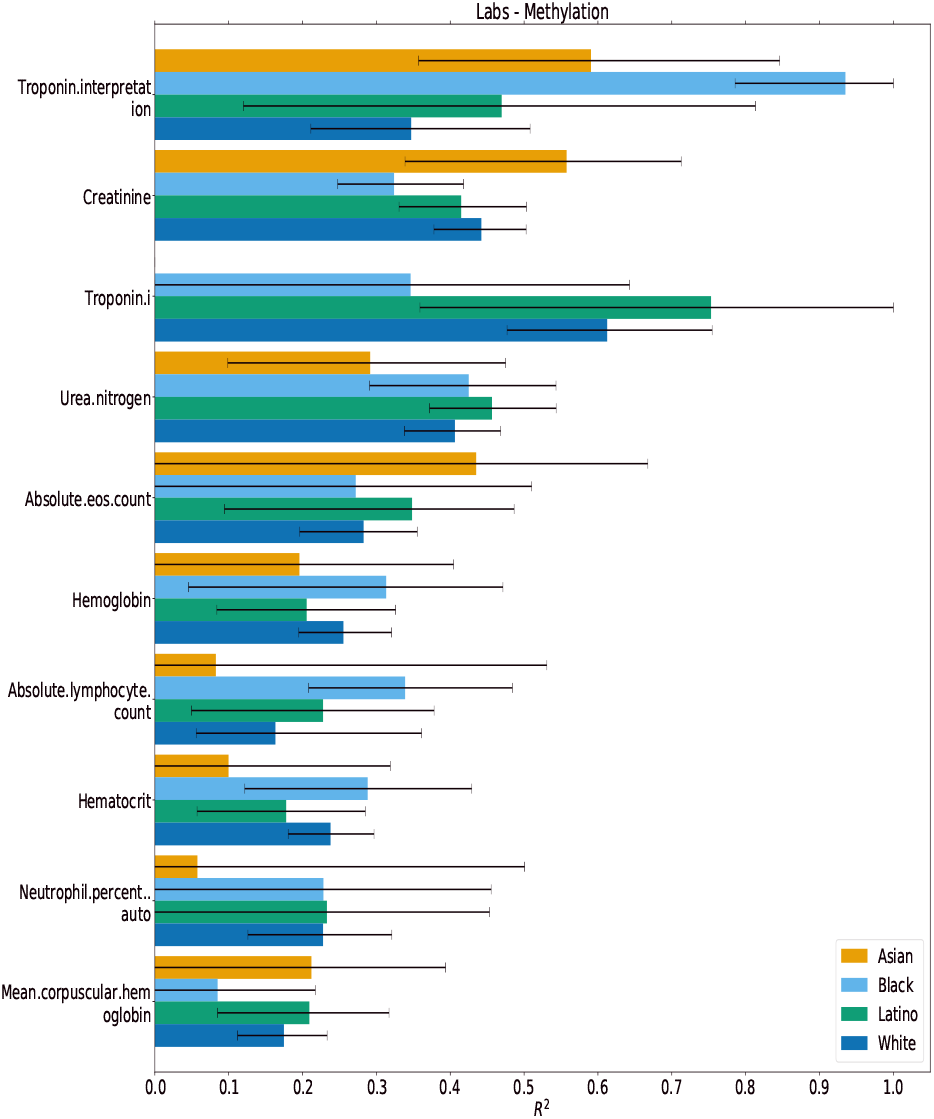
Best methylation-predicted lab panels within ancestral populations. After training a model on the entire heterogeneous set of individuals, we evaluated the predictive performance within each population separately. We observed no significant differences within self-reported ancestral groupings.

**Figure S13.**
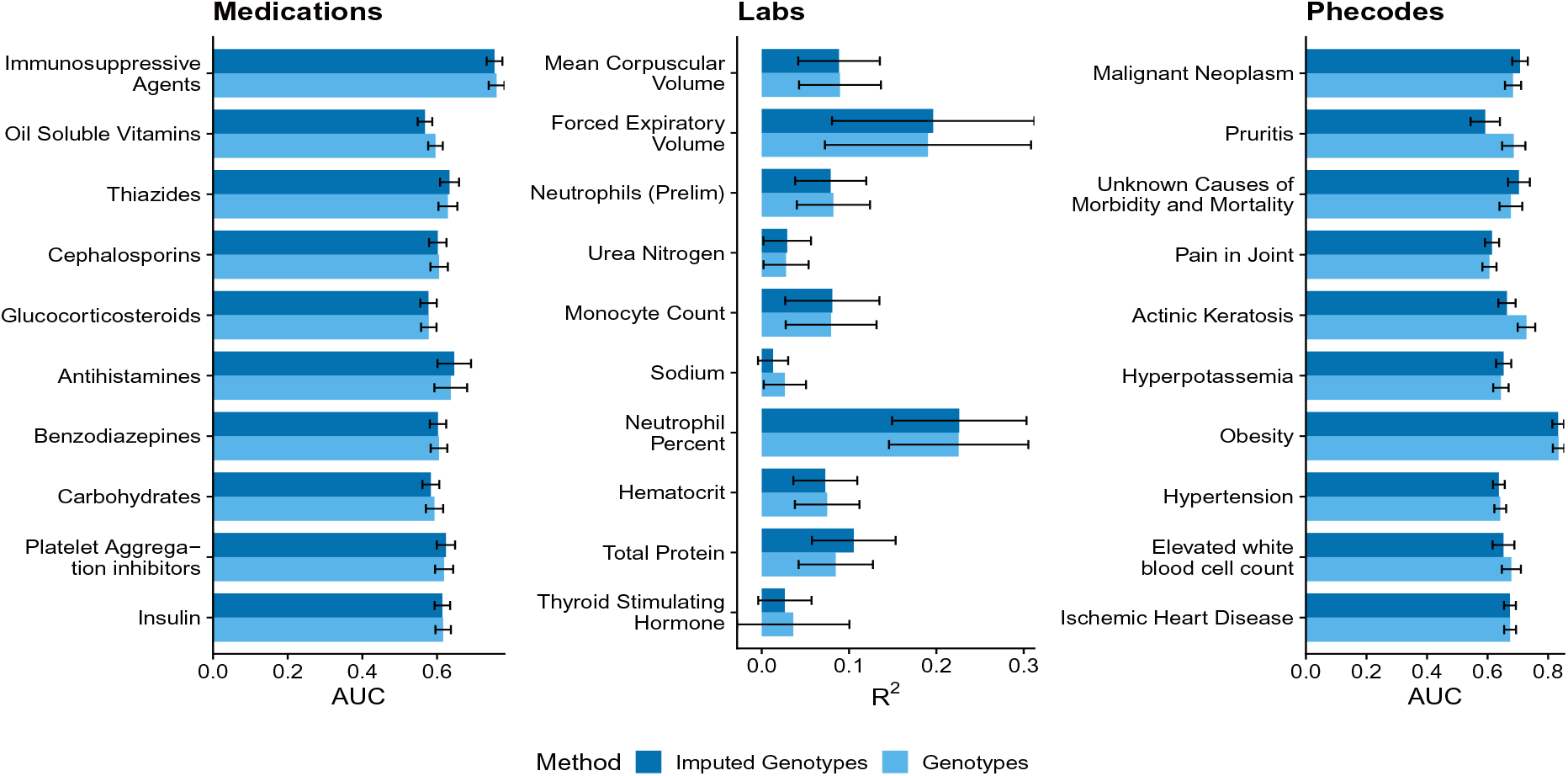
Imputation accuracy when constructing PRS using chipped genotypes compared to using imputed genotypes. We fit models using the imputed genotypes on the outcomes that were best imputed by the chipped genotypes and that significantly improved over the baseline model. Using the imputed genotypes did not result in significant differences in imputation accuracy when compared to the chipped genotypes.

**Figure S14.**
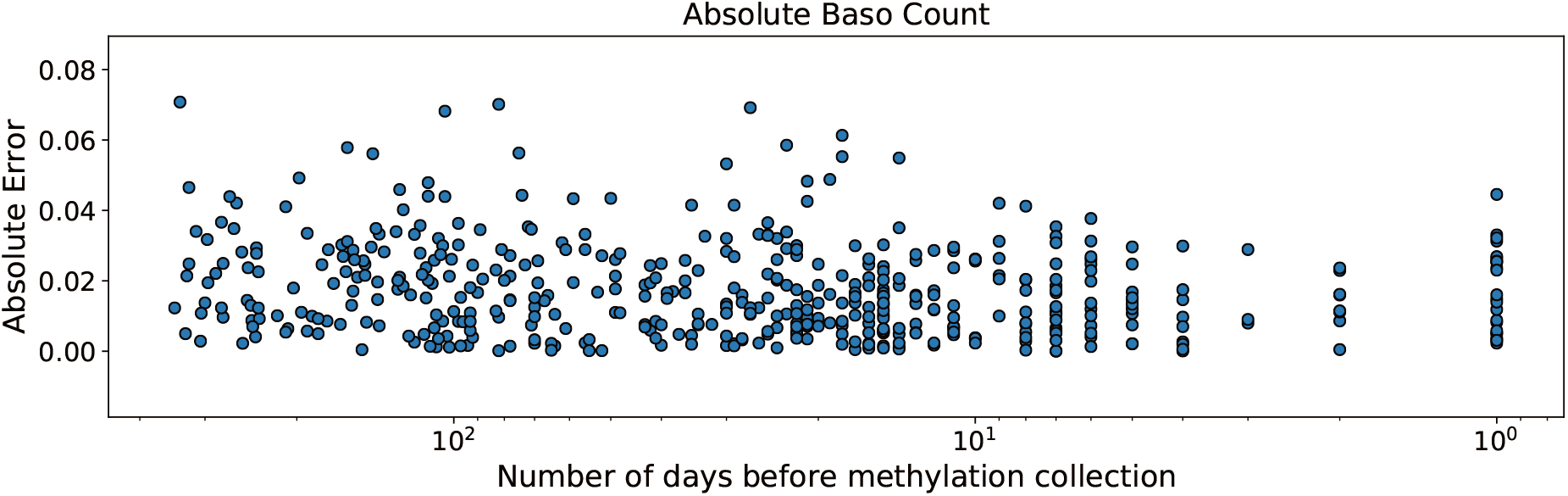
Imputation error as a function of time since methylation sample collection date. We analyzed the lab value imputation residuals to see if the errors were associated with the number of days between the lab result and the methylation collection date. After correcting for multiple hypotheses, only one lab showed a significant association between the imputed value residuals and time between collection dates (absolute basophil count, Pearson *R*=0.178, Bonferroni corrected p-value=0.0045). Here we show the absolute residual error as a function of the number of days the lab resulted before methylation collection for the only significantly associated lab, absolute basophil count (note the log scaling on the x-axis)

## Notes

### Competing Interest Statement

There is a conflict of interest
I.H. is the president of Clarity Healthcare Analytics Inc, a company that assists hospitals with extracting and using data from their electronic medical records. The company currently owns the rights to the PDW software that was used to extract data from the electronic health record. I.H. receives research funding from Merck Pharmaceuticals. M.C. is a consultant for Edwards Lifesciences (Irvine, CA) and Masimo Corp (Irvine, CA), and has funded research from Edwards Lifesciences and Masimo Corp. He is also the founder of Sironis and he owns patents and receives royalties for closed loop hemodynamic management technologies that have been licensed to Edwards Lifesciences. E.H. is senior vice president of AI/ML at OptumLabs (Minnetonka, MN). The other authors declare no competing interests concerning this article.

### Funding Statement

See manuscript for funding statements

### Author Declarations

Retrospective data collection and analysis was approved by the UCLA IRB. We include a reference to [67] which includes the following: Patient Recruitment and Sample Collection for Precision Health Activities at UCLA is an approved study by the UCLA Institutional Review Board (UCLA IRB). IRB#17-001013. All necessary patient/participant consent has been obtained and the appropriate institutional forms have been archived.

